# Challenges and opportunities for monitoring diet and physical activity in younger adults as part of a future brain health study: A UK and US Survey

**DOI:** 10.64898/2025.12.16.25342370

**Authors:** Oliver M Shannon, Alex Griffiths, Jamie Matu, Kirstie Cronin, Katie Bridgeman, Laura Booi, Francesca R. Farina, Sarah Gregory

## Abstract

**Aim:** Young adulthood is a formative life stage during which modifiable behaviours including diet and physical activity (PA) can have lasting impacts on brain health. However, this age group remains understudied in dementia research. This study aimed to explore how younger adults track their diet and PA, and evaluated attitudes, barriers, and enablers to different assessment tools.

**Subject and methods:** An online questionnaire assessed diet and PA tracking behaviours, attitudes, and barriers/enablers in younger adults (18-39 years) across the UK and US. Responses were compared between countries, ages, sexes, and ethnicities.

**Results:** 1006 younger adults (UK n=500, US n=506) participated, with 90.3% reporting they would be likely/very likely to participate in a study exploring lifestyle and brain health. Remote technology-based data collection methods, particularly apps and smartwatches, were widely acceptable. Most participants were willing to provide annual dietary and activity data. Key diet-tracking barriers included estimating portion sizes and tracking outside the home. Key PA-tracking barriers included day-to-day variability and forgetting to record activity. Enablers included receiving incentives and using passive tracking methods. Participants from the US, of a minority ethnic group or aged 18-29 years reported greater barriers to tracking.

**Conclusions:** Younger adults are interested in participating in brain health research and find technology-based diet and PA tracking acceptable in this context. Addressing identified barriers will be key to building a diverse, scalable cohort. Pilot testing is now needed to optimise feasibility and engagement. These findings will inform the design of a future brain health-focussed cohort study.

## INTRODUCTION

The maintenance of lifelong brain health is a major research and public health priority (Livingston et al. 2024). To this end, considerable research has focused on the identification of tractable risk and resilience factors for poor brain health and neurodegenerative diseases. Typically, this research has focused on the identification of behaviour, lifestyle or environmental factors in mid-to-later life which are associated with relevant brain health outcomes (e.g., cognition, mood, neuroimaging, biomarkers or presence of neurodegenerative diseases). Young adulthood (ages 18-39 years) is an important, formative life stage characterised by shifts in behaviours such as smoking, alcohol use and risk taking, which can impact longer-term health outcomes (Farina et al. 2024). Exposure to modifiable risk factors, and adoption of health-influencing behaviours, during this period is increasingly recognised as critical for future brain health and risk of dementia. Despite this, younger adults remain an overlooked population in dementia research (Farina et al. 2024). The Next Generation Brain Health Programme (NextGen) aims to address this research gap by identifying risk and resilience factors for brain health that are most important in young adulthood. Such knowledge is vital to develop interventions to promote brain health and reduce risk of future dementia in this group.

Diet represents a tractable risk factor for cognitive decline, mood disorders and neurodegenerative diseases (Siervo et al. 2021; Shannon et al. 2025). For example, data from our group has shown that adherence to healthy dietary patterns such as the Mediterranean diet in mid-to-later life is associated with greater cognitive function (equivalent in magnitude to several fewer years of cognitive ageing) and a ∼20% lower risk of incident all cause dementia (Shannon et al. 2019, 2023). Similarly, intervention with a Mediterranean diet has been shown to improve cognitive outcomes in older adults, especially those with poor cardiovascular health (which is prognostic for future dementia) (Jennings et al. 2024). Although some research has demonstrated benefits of healthy dietary choices (e.g., higher intake of fruits and vegetables (Mao et al. 2019)) on brain health outcomes in younger adults, there is comparatively less research available in this age demographic compared with older adults. Physical activity (PA) is also recognised as an important modifiable risk factor for brain health, with higher PA associated with better cognition (Erickson et al. 2019) and reduced dementia risk (Xu et al. 2017; Iso-Markku et al. 2022). Nevertheless, most research on PA on brain health outcomes has been conducted in children and older adults, with a lack of high quality data in other age groups (including younger adults) (Erickson et al. 2019).

Assessing diet and PA in younger adults in the context of a brain health promotion study is likely to present unique challenges and opportunities compared with measuring these behaviours in middle-aged and older adults. For example, younger adults tend to have different dietary and activity habits compared with older adults. Equally, younger adults show greater acceptance of technology-based solutions for tracking behaviours such as diet and PA compared with older adults (Baer et al. 2022). This could create opportunities for assessing these behaviours using novel digital or remote data collection approaches. Choosing the appropriate methods for studying diet and PA in younger adults could also have important implications for recruitment and retention of participants in future studies (Adams et al. 2015)

This study aims to evaluate current methods used by younger adults (aged 18-39 years) for monitoring their diet and PA. It also aims to explore attitudes towards, and barriers and enablers to, measuring diet and PA using different approaches in the context of a brain health focussed cohort study. We aim to gather data from both the UK and USA, to provide insight into ways in which data collection methods may require tailoring to different geographical settings which may be included in a future brain health study. The findings will inform the design of a future prospective cohort study to identify risk and resilience factors for poor brain health in later life as part of the wider NextGen Brain Health Programme.

## METHODS

This study received ethical approval prior to questionnaire administration from the Newcastle University Faculty of Medical Sciences Research Ethics Committee (REF: 57735/2023). Participants received a written description of the study requirements and provided informed consent prior to completing the questionnaire.

### Questionnaire design

A questionnaire was developed to evaluate current diet and PA tracking behaviour, alongside attitudes towards, and barriers and enablers to, measuring diet and PA in the context of a brain health study. An initial set of questions was developed by the research team, drawing on existing literature, subject matter and population expertise within the team. Questions on barriers and enablers were developed with reference to the COM-B model and Theoretical Domains Framework (TDF) (Michie et al. 2011, 2015; Cane et al. 2012) to ensure coverage of behavioural determinants relevant to diet and PA tracking. The questionnaire was pilot tested with n=4 younger adults to ensure it was clear and understandable and subsequently refined following participant feedback. Pilot testing also allowed us to estimate completion time.

A final version of the questionnaire was created using an online survey administration platform (Online Surveys, Bristol) and was split into five sections. Section one evaluated participant characteristics (e.g., age, sex, ethnicity, household income and living situation), which were used to describe the cohort and for comparing responses between key population sub-groups (see Statistical Analysis). Separate versions of the questionnaire were administered to UK and US participants, allowing us to capture participant country. Minor adaptations were made to the demographic questions to ensure suitability to each audience, such as using different units for expressing household income (e.g., pounds sterling vs. US dollars) and different terminology for stages of education (e.g., secondary school vs. high school). Section two evaluated current diet and PA tracking behaviour. Section three evaluated general study design preferences (e.g., preference for remote, in person or blended data collection approaches). Section four focused on preferences for specific diet and PA tracking methods, whilst section five evaluated barriers and enablers for diet and PA tracking.

### Questionnaire administration

The questionnaire was administered to younger adults (aged 18-39 years) in the UK and US via Prolific, an online crowd-sourcing platform of verified research participants (Peer et al. 2017). Both US and UK participants were surveyed on the same day (4^th^ June, 2025) to minimise temporal influence on response. Participants were paid a modest fee for completing the questionnaire (∼£9/hour pro rata in both the UK and the US). To be eligible to participate, participants were required to be of an appropriate age, residing in the UK or US, and be registered with the Prolific platform. Data was only collected for participants who finished the entire questionnaire, and a response was not registered if participants stopped before completion.

### Statistical analysis

All statistical analyses were conducted in R (version 4.4.0). Descriptive statistics were calculated to summarise the overall distribution of responses. Between-group differences based on country (UK vs. US), age (younger [18-29 years] vs. older [30-39 years]), sex (male vs. female), and ethnicity (white vs. minority ethnic groups) were assessed using Mann-Whitney U test for ordinal variables and Chi-squared test for non-ordered (i.e., nominal) categorical variables. *P*<0.05 was accepted for statistical significance.

## RESULTS

### Participant characteristics

This study recruited 1006 participants (500 UK and 506 US based) with an average age of 30.0 (5.7) years. There were marginally more female than male participants, with most participants reporting white ethnicity. Most participants were in full time employment and the most common highest qualification was an undergraduate university or college degree. Participant characteristics stratified by country can be seen in Table 1.

**Table 1.**
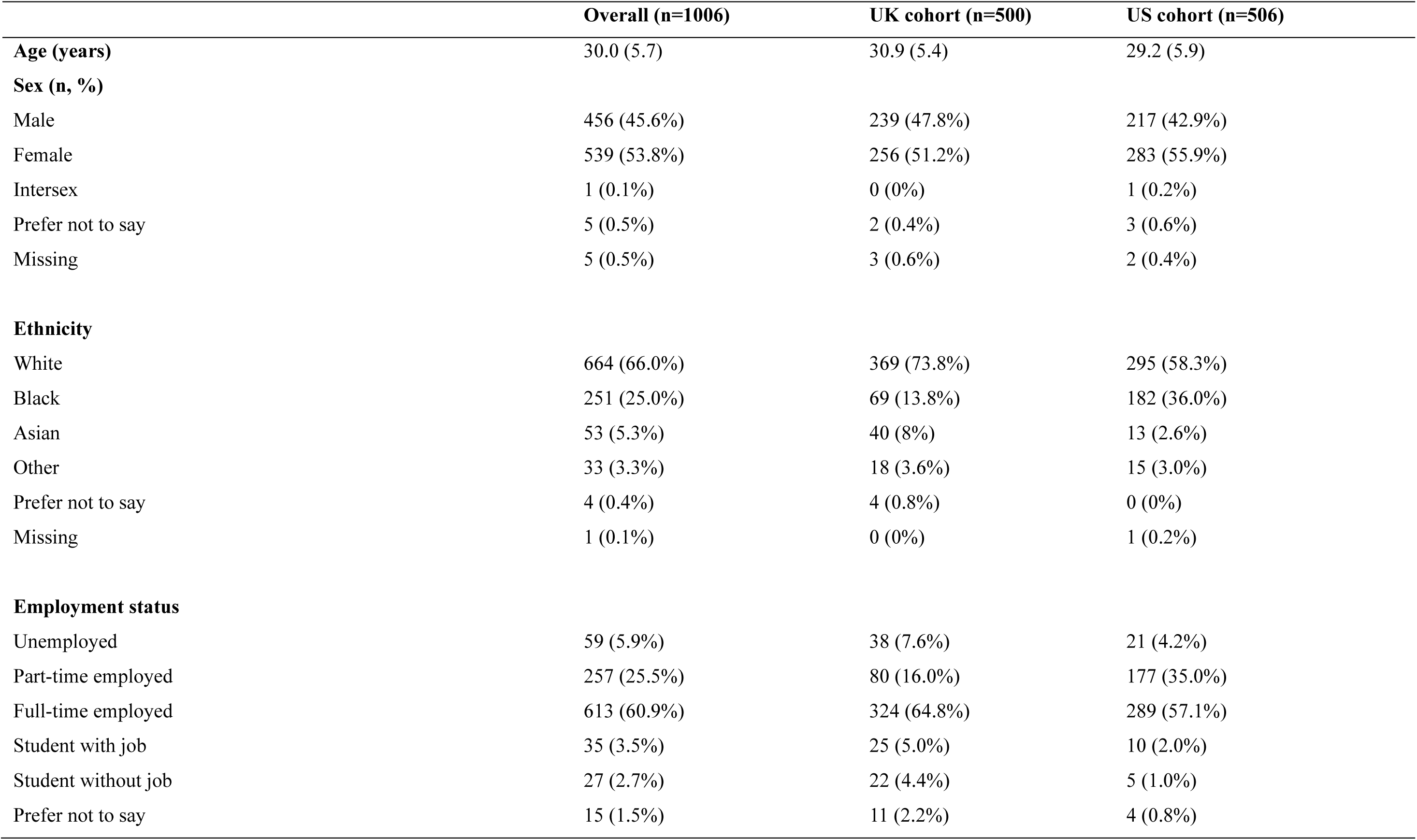
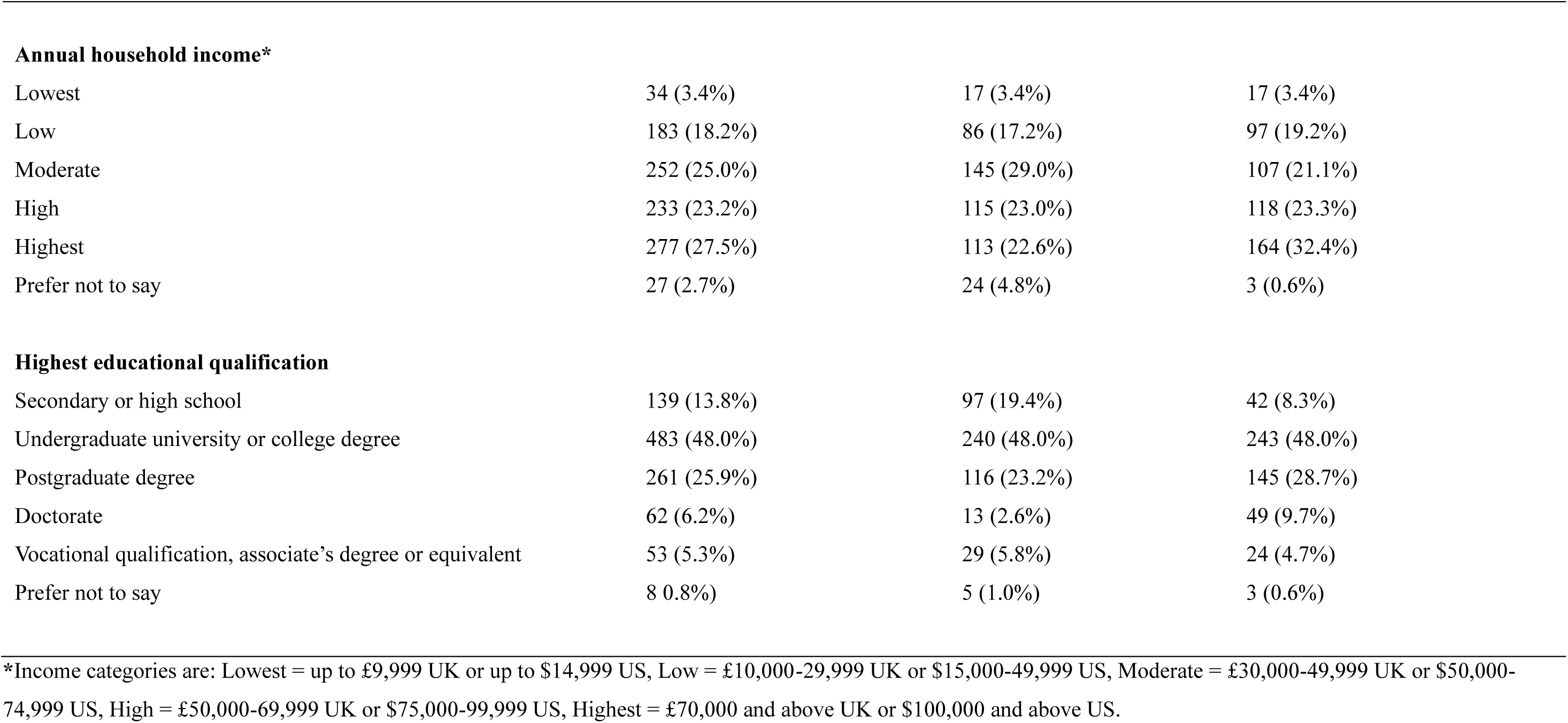
Participant characteristics

|  | Overall (n=1006) | UK cohort (n=500) | US cohort (n=506) |
| --- | --- | --- | --- |
| **Age (years)** | 30.0 (5.7) | 30.9 (5.4) | 29.2 (5.9) |
| **Sex (n, %)** |  |  |  |
| Male | 456 (45.6%) | 239 (47.8%) | 217 (42.9%) |
| Female | 539 (53.8%) | 256 (51.2%) | 283 (55.9%) |
| Intersex | 1 (0.1%) | 0 (0%) | 1 (0.2%) |
| Prefer not to say | 5 (0.5%) | 2 (0.4%) | 3 (0.6%) |
| Missing | 5 (0.5%) | 3 (0.6%) | 2 (0.4%) |
| **Ethnicity** |  |  |  |
| White | 664 (66.0%) | 369 (73.8%) | 295 (58.3%) |
| Black | 251 (25.0%) | 69 (13.8%) | 182 (36.0%) |
| Asian | 53 (5.3%) | 40 (8%) | 13 (2.6%) |
| Other | 33 (3.3%) | 18 (3.6%) | 15 (3.0%) |
| Prefer not to say | 4 (0.4%) | 4 (0.8%) | 0 (0%) |
| Missing | 1 (0.1%) | 0 (0%) | 1 (0.2%) |
| **Employment status** |  |  |  |
| Unemployed | 59 (5.9%) | 38 (7.6%) | 21 (4.2%) |
| Part-time employed | 257 (25.5%) | 80 (16.0%) | 177 (35.0%) |
| Full-time employed | 613 (60.9%) | 324 (64.8%) | 289 (57.1%) |
| Student with job | 35 (3.5%) | 25 (5.0%) | 10 (2.0%) |
| Student without job | 27 (2.7%) | 22 (4.4%) | 5 (1.0%) |
| Prefer not to say | 15 (1.5%) | 11 (2.2%) | 4 (0.8%) |
| **Annual household income\*** |  |  |  |
| Lowest | 34 (3.4%) | 17 (3.4%) | 17 (3.4%) |
| Low | 183 (18.2%) | 86 (17.2%) | 97 (19.2%) |
| Moderate | 252 (25.0%) | 145 (29.0%) | 107 (21.1%) |
| High | 233 (23.2%) | 115 (23.0%) | 118 (23.3%) |
| Highest | 277 (27.5%) | 113 (22.6%) | 164 (32.4%) |
| Prefer not to say | 27 (2.7%) | 24 (4.8%) | 3 (0.6%) |
| **Highest educational qualification** |  |  |  |
| Secondary or high school | 139 (13.8%) | 97 (19.4%) | 42 (8.3%) |
| Undergraduate university or college degree | 483 (48.0%) | 240 (48.0%) | 243 (48.0%) |
| Postgraduate degree | 261 (25.9%) | 116 (23.2%) | 145 (28.7%) |
| Doctorate | 62 (6.2%) | 13 (2.6%) | 49 (9.7%) |
| Vocational qualification, associate's degree or equivalent | 53 (5.3%) | 29 (5.8%) | 24 (4.7%) |
| Prefer not to say | 8 (0.8%) | 5 (1.0%) | 3 (0.6%) |

\*Income categories are: Lowest = up to £9,999 UK or up to \$14,999 US, Low = £10,000-29,999 UK or \$15,000-49,999 US, Moderate = £30,000-49,999 UK or \$50,000-74,999 US, High = £50,000-69,999 UK or \$75,000-99,999 US, Highest = £70,000 and above UK or \$100,000 and above US.

### Current behaviour

Most participants reported consuming a somewhat healthy diet (Table 2). Participants most frequently stated that they were moderately confident in their ability to track their diet and very motivated to do so. Participants most frequently reported tracking their diet occasionally, primarily using a mobile app. The main reason reported for tracking diet was to help manage weight.

**Table 2.**
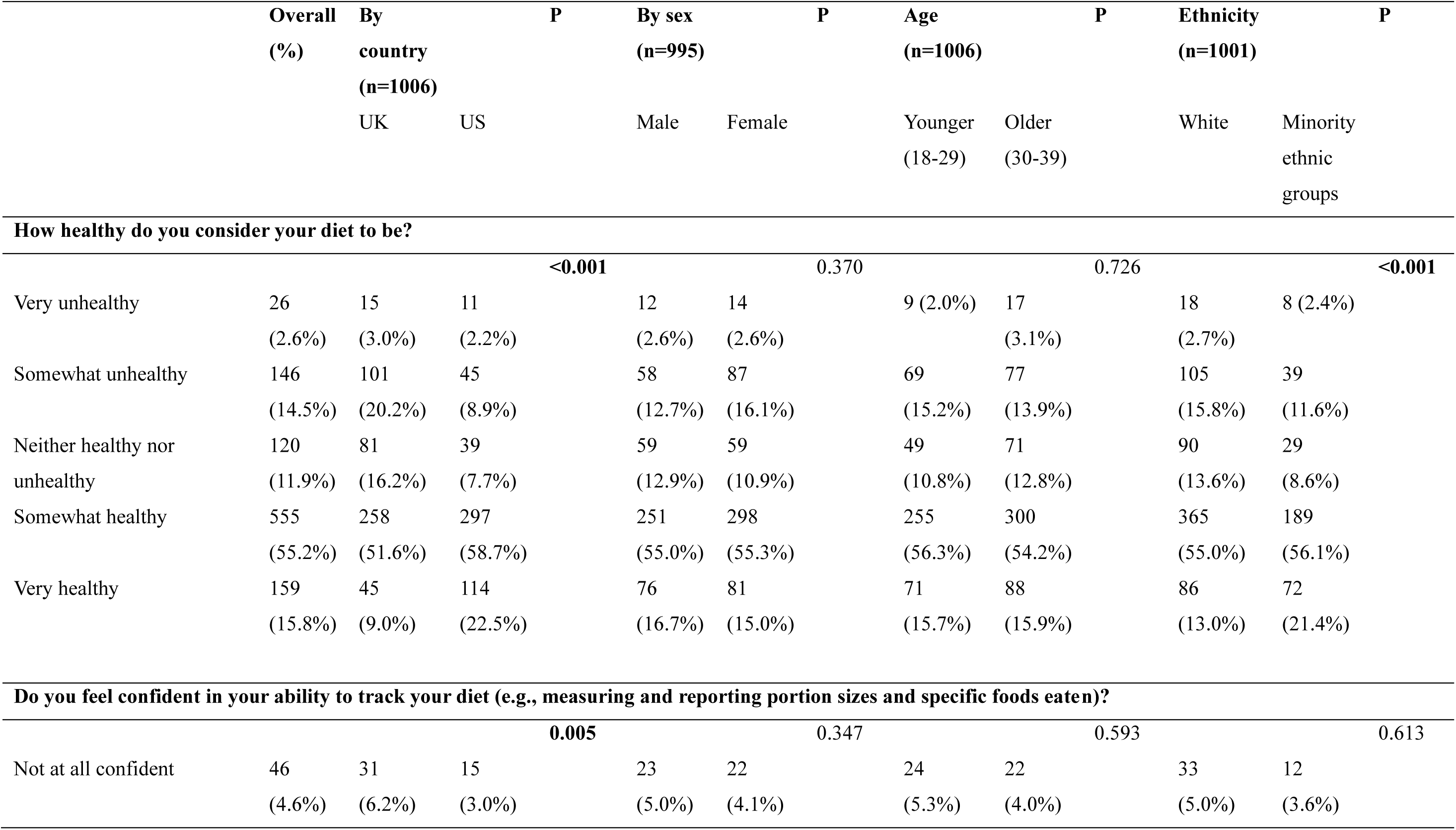
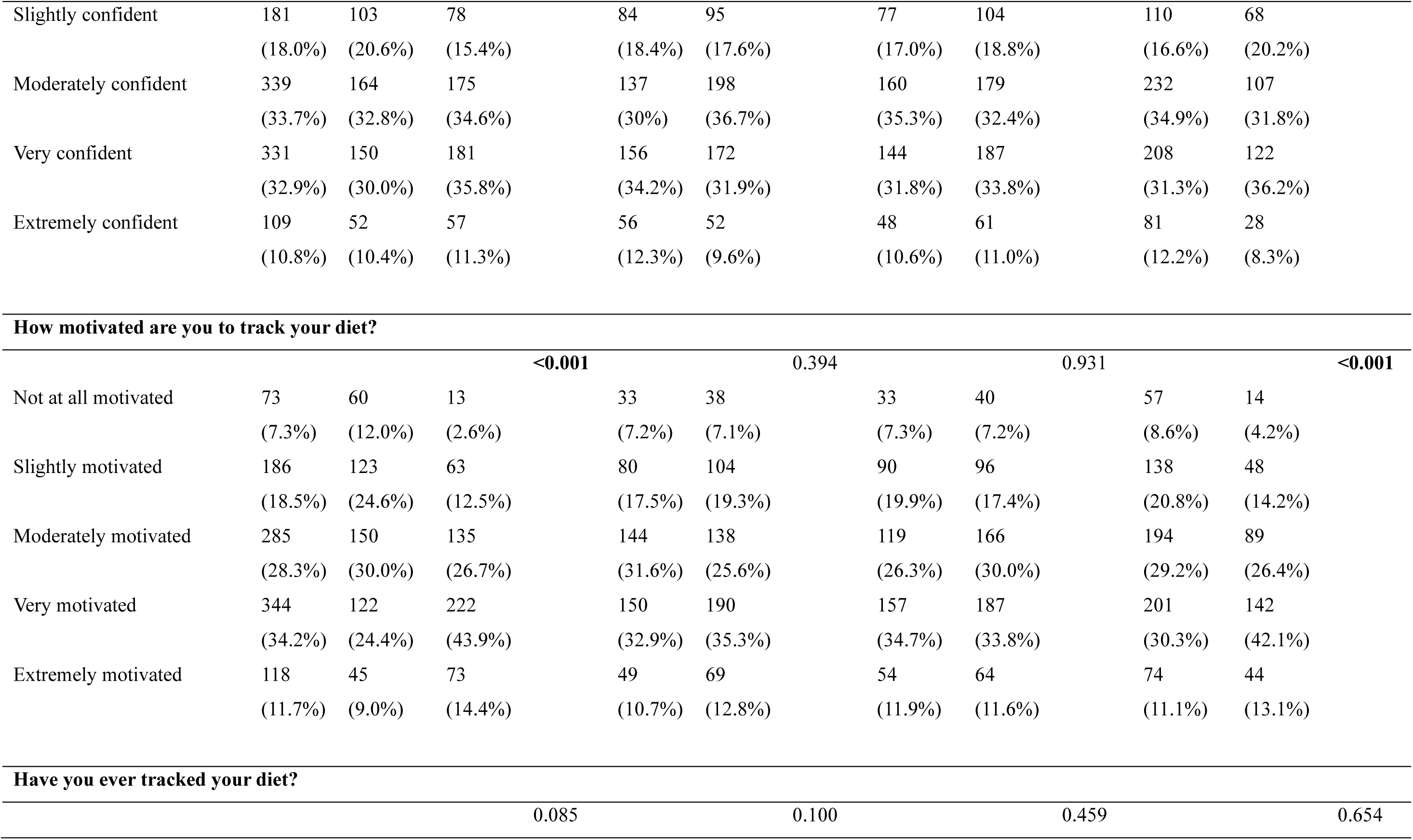
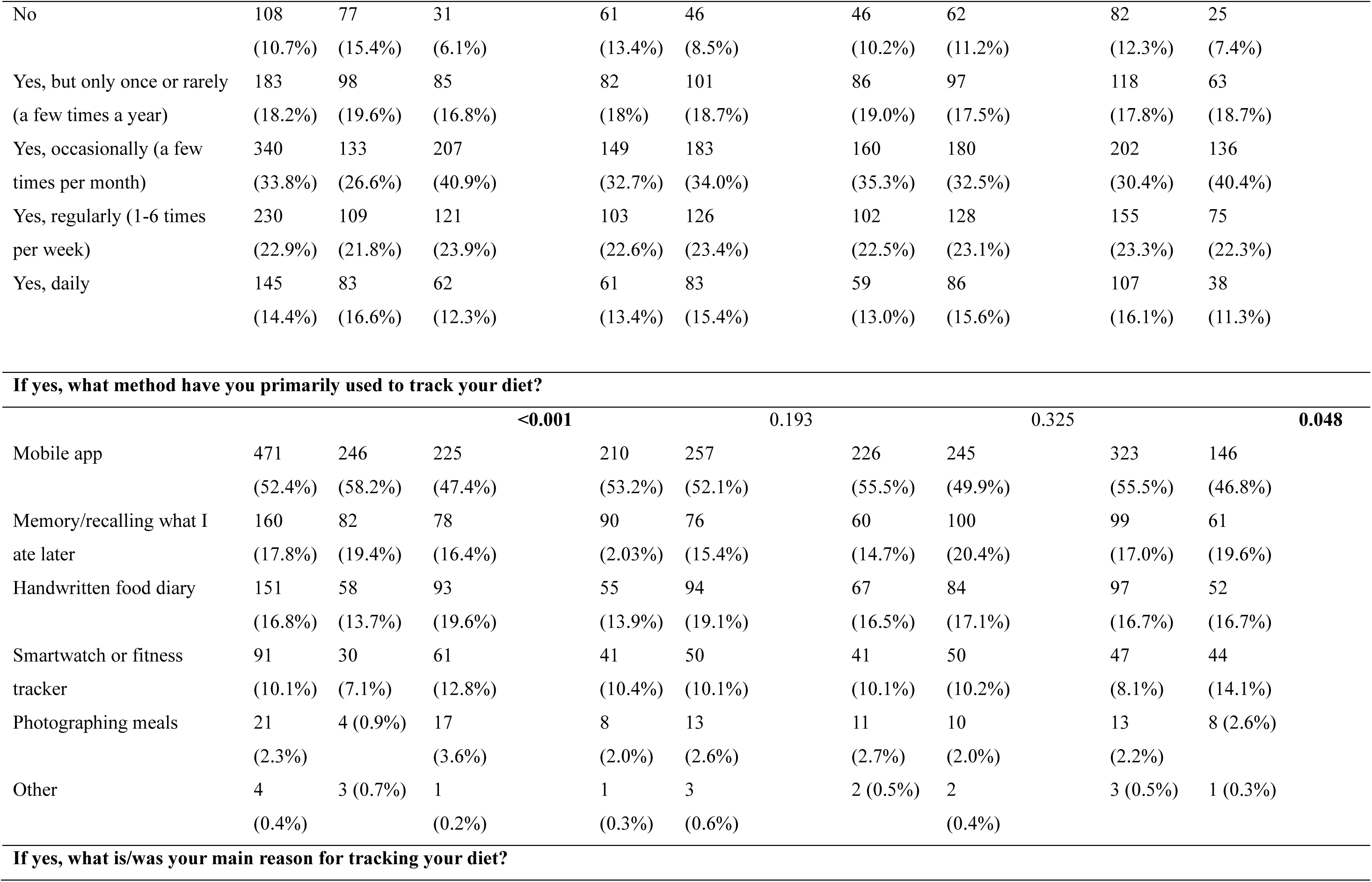
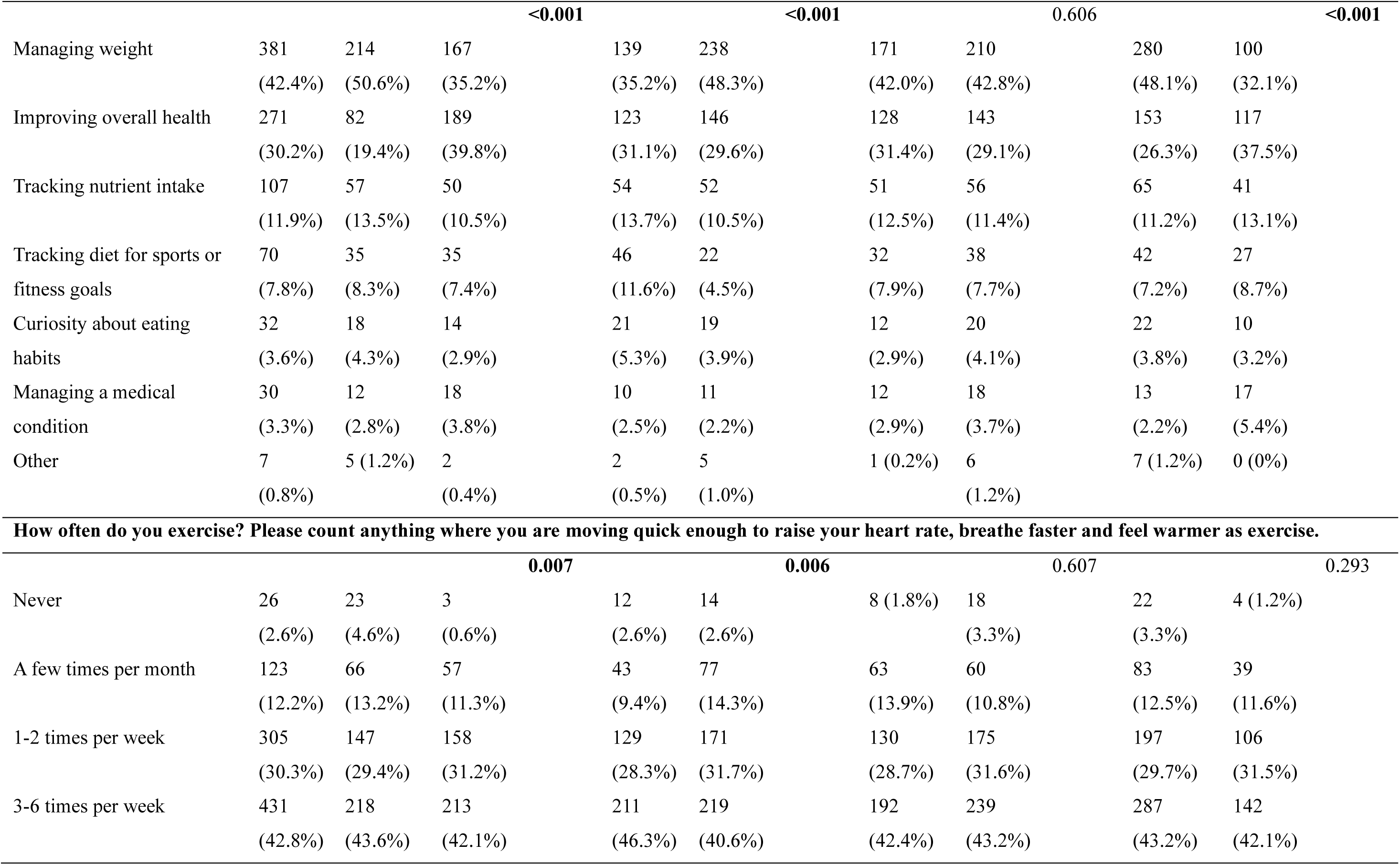
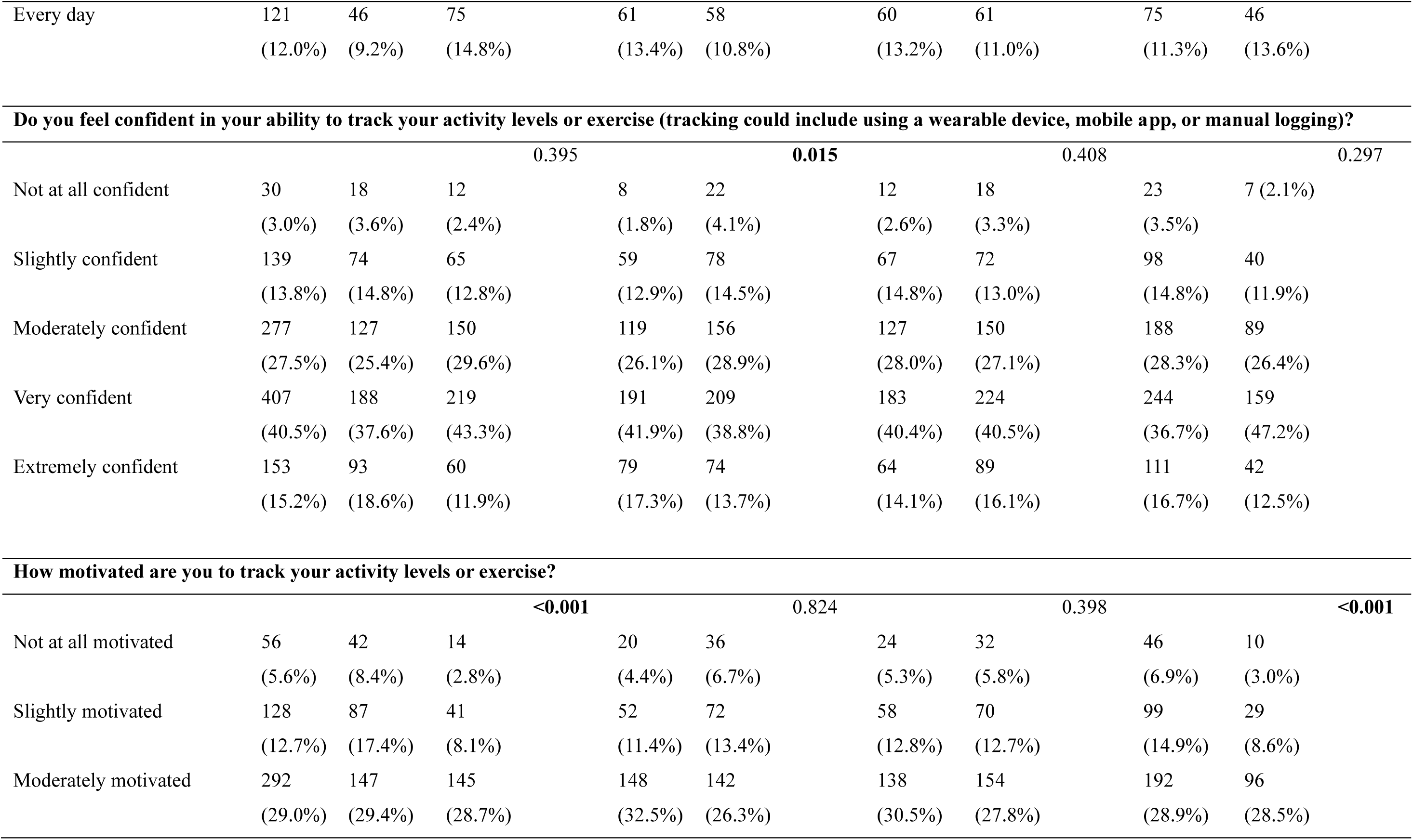
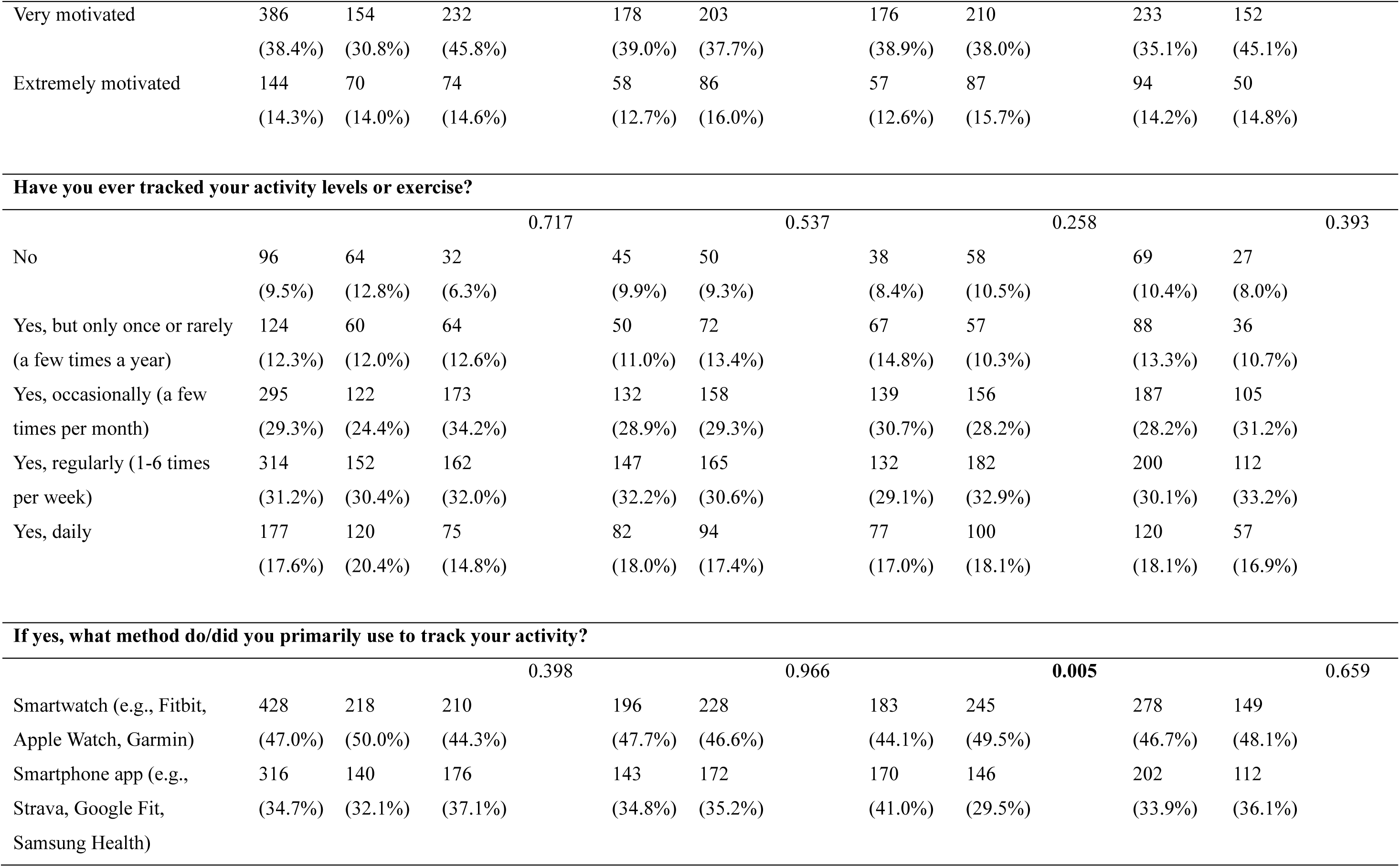
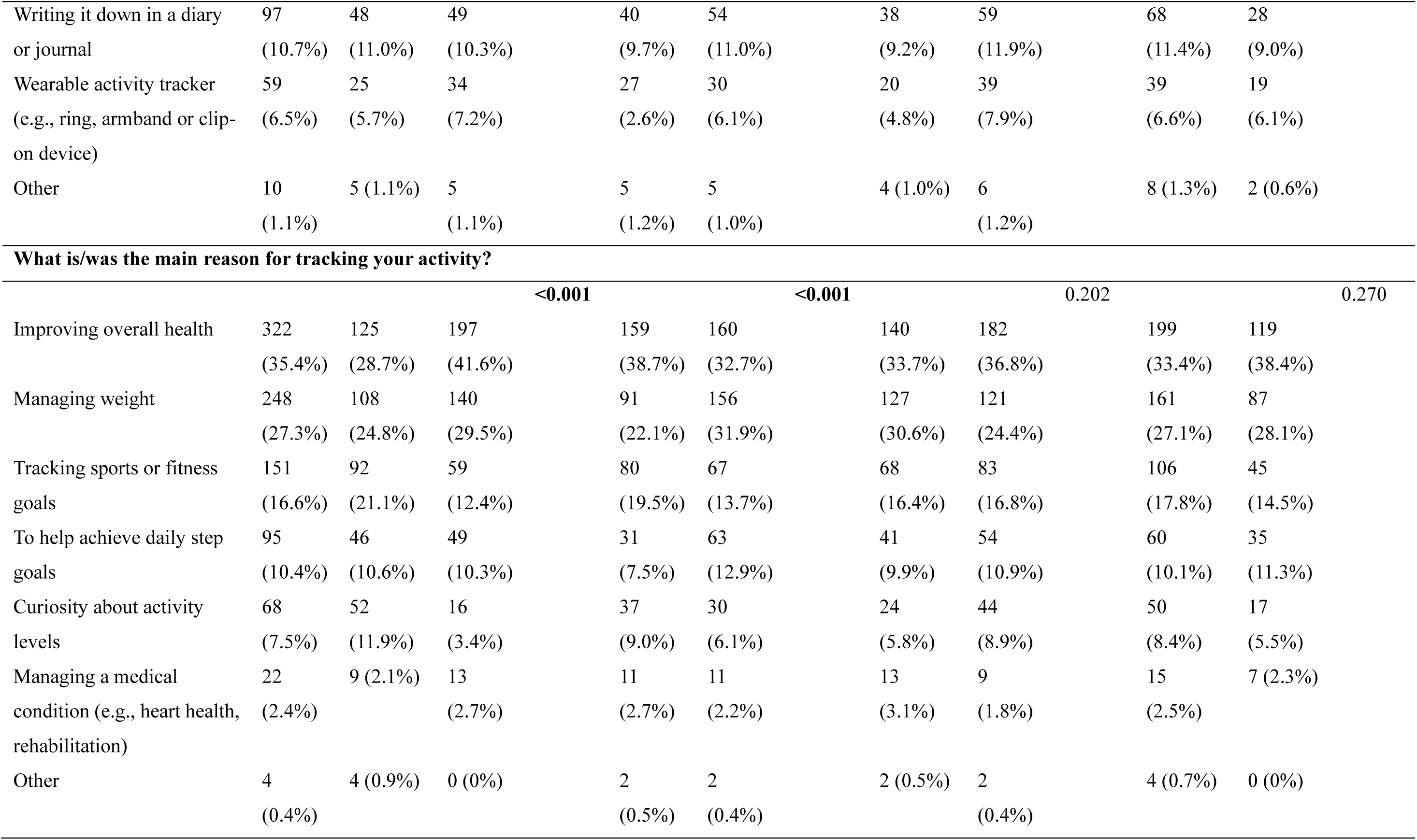
Current diet and PA behaviour and tracking

|  | Overall<br>(%) | By<br>country<br>(n=1006) |  | P | By sex<br>(n=995) |  | P | Age<br>(n=1006) |  | P | Ethnicity<br>(n=1001) |  | P |
| --- | --- | --- | --- | --- | --- | --- | --- | --- | --- | --- | --- | --- | --- |
|  |  | UK | US |  | Male | Female |  | Younger<br>(18-29) | Older<br>(30-39) |  | White | Minority<br>ethnic<br>groups |  |
| **How healthy do you consider your diet to be?** |  |  |  |  |  |  |  |  |  |  |  |  |  |
|  |  |  |  | **<0.001** |  |  | 0.370 |  |  | 0.726 |  |  | **<0.001** |
| Very unhealthy | 26<br>(2.6%) | 15<br>(3.0%) | 11<br>(2.2%) |  | 12<br>(2.6%) | 14<br>(2.6%) |  | 9 (2.0%) | 17<br>(3.1%) |  | 18<br>(2.7%) | 8 (2.4%) |  |
| Somewhat unhealthy | 146<br>(14.5%) | 101<br>(20.2%) | 45<br>(8.9%) |  | 58<br>(12.7%) | 87<br>(16.1%) |  | 69<br>(15.2%) | 77<br>(13.9%) |  | 105<br>(15.8%) | 39<br>(11.6%) |  |
| Neither healthy nor<br>unhealthy | 120<br>(11.9%) | 81<br>(16.2%) | 39<br>(7.7%) |  | 59<br>(12.9%) | 59<br>(10.9%) |  | 49<br>(10.8%) | 71<br>(12.8%) |  | 90<br>(13.6%) | 29<br>(8.6%) |  |
| Somewhat healthy | 555<br>(55.2%) | 258<br>(51.6%) | 297<br>(58.7%) |  | 251<br>(55.0%) | 298<br>(55.3%) |  | 255<br>(56.3%) | 300<br>(54.2%) |  | 365<br>(55.0%) | 189<br>(56.1%) |  |
| Very healthy | 159<br>(15.8%) | 45<br>(9.0%) | 114<br>(22.5%) |  | 76<br>(16.7%) | 81<br>(15.0%) |  | 71<br>(15.7%) | 88<br>(15.9%) |  | 86<br>(13.0%) | 72<br>(21.4%) |  |
| **Do you feel confident in your ability to track your diet (e.g., measuring and reporting portion sizes and specific foods eaten)?** |  |  |  |  |  |  |  |  |  |  |  |  |  |
|  |  |  |  | **0.005** |  |  | 0.347 |  |  | 0.593 |  |  | 0.613 |
| Not at all confident | 46<br>(4.6%) | 31<br>(6.2%) | 15<br>(3.0%) |  | 23<br>(5.0%) | 22<br>(4.1%) |  | 24<br>(5.3%) | 22<br>(4.0%) |  | 33<br>(5.0%) | 12<br>(3.6%) |  |

|  |  |  |  |  |  |  |  |  |  |
| --- | --- | --- | --- | --- | --- | --- | --- | --- | --- |
| Slightly confident | 181<br>(18.0%) | 103<br>(20.6%) | 78<br>(15.4%) | 84<br>(18.4%) | 95<br>(17.6%) | 77<br>(17.0%) | 104<br>(18.8%) | 110<br>(16.6%) | 68<br>(20.2%) |
| Moderately confident | 339<br>(33.7%) | 164<br>(32.8%) | 175<br>(34.6%) | 137<br>(30%) | 198<br>(36.7%) | 160<br>(35.3%) | 179<br>(32.4%) | 232<br>(34.9%) | 107<br>(31.8%) |
| Very confident | 331<br>(32.9%) | 150<br>(30.0%) | 181<br>(35.8%) | 156<br>(34.2%) | 172<br>(31.9%) | 144<br>(31.8%) | 187<br>(33.8%) | 208<br>(31.3%) | 122<br>(36.2%) |
| Extremely confident | 109<br>(10.8%) | 52<br>(10.4%) | 57<br>(11.3%) | 56<br>(12.3%) | 52<br>(9.6%) | 48<br>(10.6%) | 61<br>(11.0%) | 81<br>(12.2%) | 28<br>(8.3%) |

| How motivated are you to track your diet? |  |  |  |  |  |  |  |  |  |
| --- | --- | --- | --- | --- | --- | --- | --- | --- | --- |
|  | <0.001 |  |  | 0.394 |  | 0.931 |  | <0.001 |  |
| Not at all motivated | 73<br>(7.3%) | 60<br>(12.0%) | 13<br>(2.6%) | 33<br>(7.2%) | 38<br>(7.1%) | 33<br>(7.3%) | 40<br>(7.2%) | 57<br>(8.6%) | 14<br>(4.2%) |
| Slightly motivated | 186<br>(18.5%) | 123<br>(24.6%) | 63<br>(12.5%) | 80<br>(17.5%) | 104<br>(19.3%) | 90<br>(19.9%) | 96<br>(17.4%) | 138<br>(20.8%) | 48<br>(14.2%) |
| Moderately motivated | 285<br>(28.3%) | 150<br>(30.0%) | 135<br>(26.7%) | 144<br>(31.6%) | 138<br>(25.6%) | 119<br>(26.3%) | 166<br>(30.0%) | 194<br>(29.2%) | 89<br>(26.4%) |
| Very motivated | 344<br>(34.2%) | 122<br>(24.4%) | 222<br>(43.9%) | 150<br>(32.9%) | 190<br>(35.3%) | 157<br>(34.7%) | 187<br>(33.8%) | 201<br>(30.3%) | 142<br>(42.1%) |
| Extremely motivated | 118<br>(11.7%) | 45<br>(9.0%) | 73<br>(14.4%) | 49<br>(10.7%) | 69<br>(12.8%) | 54<br>(11.9%) | 64<br>(11.6%) | 74<br>(11.1%) | 44<br>(13.1%) |
| Have you ever tracked your diet? |  |  |  |  |  |  |  |  |  |
|  | 0.085 |  |  | 0.100 |  | 0.459 |  | 0.654 |  |
| No | 108<br>(10.7%) | 77<br>(15.4%) | 31<br>(6.1%) | 61<br>(13.4%) | 46<br>(8.5%) | 46<br>(10.2%) | 62<br>(11.2%) | 82<br>(12.3%) | 25<br>(7.4%) |
| Yes, but only once or rarely<br>(a few times a year) | 183<br>(18.2%) | 98<br>(19.6%) | 85<br>(16.8%) | 82<br>(18%) | 101<br>(18.7%) | 86<br>(19.0%) | 97<br>(17.5%) | 118<br>(17.8%) | 63<br>(18.7%) |
| Yes, occasionally (a few<br>times per month) | 340<br>(33.8%) | 133<br>(26.6%) | 207<br>(40.9%) | 149<br>(32.7%) | 183<br>(34.0%) | 160<br>(35.3%) | 180<br>(32.5%) | 202<br>(30.4%) | 136<br>(40.4%) |
| Yes, regularly (1-6 times<br>per week) | 230<br>(22.9%) | 109<br>(21.8%) | 121<br>(23.9%) | 103<br>(22.6%) | 126<br>(23.4%) | 102<br>(22.5%) | 128<br>(23.1%) | 155<br>(23.3%) | 75<br>(22.3%) |
| Yes, daily | 145<br>(14.4%) | 83<br>(16.6%) | 62<br>(12.3%) | 61<br>(13.4%) | 83<br>(15.4%) | 59<br>(13.0%) | 86<br>(15.6%) | 107<br>(16.1%) | 38<br>(11.3%) |

**If yes, what method have you primarily used to track your diet?**
|  | <0.001 |  |  | 0.193 |  | 0.325 |  | 0.048 |  |
| --- | --- | --- | --- | --- | --- | --- | --- | --- | --- |
| Mobile app | 471<br>(52.4%) | 246<br>(58.2%) | 225<br>(47.4%) | 210<br>(53.2%) | 257<br>(52.1%) | 226<br>(55.5%) | 245<br>(49.9%) | 323<br>(55.5%) | 146<br>(46.8%) |
| Memory/recalling what I<br>ate later | 160<br>(17.8%) | 82<br>(19.4%) | 78<br>(16.4%) | 90<br>(2.03%) | 76<br>(15.4%) | 60<br>(14.7%) | 100<br>(20.4%) | 99<br>(17.0%) | 61<br>(19.6%) |
| Handwritten food diary | 151<br>(16.8%) | 58<br>(13.7%) | 93<br>(19.6%) | 55<br>(13.9%) | 94<br>(19.1%) | 67<br>(16.5%) | 84<br>(17.1%) | 97<br>(16.7%) | 52<br>(16.7%) |
| Smartwatch or fitness<br>tracker | 91<br>(10.1%) | 30<br>(7.1%) | 61<br>(12.8%) | 41<br>(10.4%) | 50<br>(10.1%) | 41<br>(10.1%) | 50<br>(10.2%) | 47<br>(8.1%) | 44<br>(14.1%) |
| Photographing meals | 21<br>(2.3%) | 4 (0.9%) | 17<br>(3.6%) | 8<br>(2.0%) | 13<br>(2.6%) | 11<br>(2.7%) | 10<br>(2.0%) | 13<br>(2.2%) | 8 (2.6%) |
| Other | 4<br>(0.4%) | 3 (0.7%) | 1<br>(0.2%) | 1<br>(0.3%) | 3<br>(0.6%) | 2 (0.5%) | 2<br>(0.4%) | 3 (0.5%) | 1 (0.3%) |

**If yes, what is/was your main reason for tracking your diet?**
|  | <0.001 |  |  | <0.001 |  |  | 0.606 |  | <0.001 |
| --- | --- | --- | --- | --- | --- | --- | --- | --- | --- |
| Managing weight | 381 | 214 | 167 | 139 | 238 | 171 | 210 | 280 | 100 |
|  | (42.4%) | (50.6%) | (35.2%) | (35.2%) | (48.3%) | (42.0%) | (42.8%) | (48.1%) | (32.1%) |
| Improving overall health | 271 | 82 | 189 | 123 | 146 | 128 | 143 | 153 | 117 |
|  | (30.2%) | (19.4%) | (39.8%) | (31.1%) | (29.6%) | (31.4%) | (29.1%) | (26.3%) | (37.5%) |
| Tracking nutrient intake | 107 | 57 | 50 | 54 | 52 | 51 | 56 | 65 | 41 |
|  | (11.9%) | (13.5%) | (10.5%) | (13.7%) | (10.5%) | (12.5%) | (11.4%) | (11.2%) | (13.1%) |
| Tracking diet for sports or fitness goals | 70 | 35 | 35 | 46 | 22 | 32 | 38 | 42 | 27 |
|  | (7.8%) | (8.3%) | (7.4%) | (11.6%) | (4.5%) | (7.9%) | (7.7%) | (7.2%) | (8.7%) |
| Curiosity about eating habits | 32 | 18 | 14 | 21 | 19 | 12 | 20 | 22 | 10 |
|  | (3.6%) | (4.3%) | (2.9%) | (5.3%) | (3.9%) | (2.9%) | (4.1%) | (3.8%) | (3.2%) |
| Managing a medical condition | 30 | 12 | 18 | 10 | 11 | 12 | 18 | 13 | 17 |
|  | (3.3%) | (2.8%) | (3.8%) | (2.5%) | (2.2%) | (2.9%) | (3.7%) | (2.2%) | (5.4%) |
| Other | 7 | 5 (1.2%) | 2 | 2 | 5 | 1 (0.2%) | 6 | 7 (1.2%) | 0 (0%) |
|  | (0.8%) |  | (0.4%) | (0.5%) | (1.0%) |  | (1.2%) |  |  |
| How often do you exercise? Please count anything where you are moving quick enough to raise your heart rate, breathe faster and feel warmer as exercise. |  |  |  |  |  |  |  |  |  |
|  | 0.007 |  |  | 0.006 |  |  | 0.607 |  | 0.293 |
| Never | 26 | 23 | 3 | 12 | 14 | 8 (1.8%) | 18 | 22 | 4 (1.2%) |
|  | (2.6%) | (4.6%) | (0.6%) | (2.6%) | (2.6%) |  | (3.3%) | (3.3%) |  |
| A few times per month | 123 | 66 | 57 | 43 | 77 | 63 | 60 | 83 | 39 |
|  | (12.2%) | (13.2%) | (11.3%) | (9.4%) | (14.3%) | (13.9%) | (10.8%) | (12.5%) | (11.6%) |
| 1-2 times per week | 305 | 147 | 158 | 129 | 171 | 130 | 175 | 197 | 106 |
|  | (30.3%) | (29.4%) | (31.2%) | (28.3%) | (31.7%) | (28.7%) | (31.6%) | (29.7%) | (31.5%) |
| 3-6 times per week | 431 | 218 | 213 | 211 | 219 | 192 | 239 | 287 | 142 |
|  | (42.8%) | (43.6%) | (42.1%) | (46.3%) | (40.6%) | (42.4%) | (43.2%) | (43.2%) | (42.1%) |
| Every day | 121 | 46 | 75 | 61 | 58 | 60 | 61 | 75 | 46 |
|  | (12.0%) | (9.2%) | (14.8%) | (13.4%) | (10.8%) | (13.2%) | (11.0%) | (11.3%) | (13.6%) |

| Do you feel confident in your ability to track your activity levels or exercise (tracking could include using a wearable device, mobile app, or manual logging)? |  |  |  |  |  |  |  |  |  |
| --- | --- | --- | --- | --- | --- | --- | --- | --- | --- |
|  | 0.395 |  |  | 0.015 |  |  | 0.408 |  | 0.297 |
| Not at all confident | 30 | 18 | 12 | 8 | 22 | 12 | 18 | 23 | 7 (2.1%) |
|  | (3.0%) | (3.6%) | (2.4%) | (1.8%) | (4.1%) | (2.6%) | (3.3%) | (3.5%) |  |
| Slightly confident | 139 | 74 | 65 | 59 | 78 | 67 | 72 | 98 | 40 |
|  | (13.8%) | (14.8%) | (12.8%) | (12.9%) | (14.5%) | (14.8%) | (13.0%) | (14.8%) | (11.9%) |
| Moderately confident | 277 | 127 | 150 | 119 | 156 | 127 | 150 | 188 | 89 |
|  | (27.5%) | (25.4%) | (29.6%) | (26.1%) | (28.9%) | (28.0%) | (27.1%) | (28.3%) | (26.4%) |
| Very confident | 407 | 188 | 219 | 191 | 209 | 183 | 224 | 244 | 159 |
|  | (40.5%) | (37.6%) | (43.3%) | (41.9%) | (38.8%) | (40.4%) | (40.5%) | (36.7%) | (47.2%) |
| Extremely confident | 153 | 93 | 60 | 79 | 74 | 64 | 89 | 111 | 42 |
|  | (15.2%) | (18.6%) | (11.9%) | (17.3%) | (13.7%) | (14.1%) | (16.1%) | (16.7%) | (12.5%) |

| How motivated are you to track your activity levels or exercise? |  |  |  |  |  |  |  |  |  |
| --- | --- | --- | --- | --- | --- | --- | --- | --- | --- |
|  | <0.001 |  |  | 0.824 |  |  | 0.398 |  | <0.001 |
| Not at all motivated | 56 | 42 | 14 | 20 | 36 | 24 | 32 | 46 | 10 |
|  | (5.6%) | (8.4%) | (2.8%) | (4.4%) | (6.7%) | (5.3%) | (5.8%) | (6.9%) | (3.0%) |
| Slightly motivated | 128 | 87 | 41 | 52 | 72 | 58 | 70 | 99 | 29 |
|  | (12.7%) | (17.4%) | (8.1%) | (11.4%) | (13.4%) | (12.8%) | (12.7%) | (14.9%) | (8.6%) |
| Moderately motivated | 292 | 147 | 145 | 148 | 142 | 138 | 154 | 192 | 96 |
|  | (29.0%) | (29.4%) | (28.7%) | (32.5%) | (26.3%) | (30.5%) | (27.8%) | (28.9%) | (28.5%) |
| Very motivated | 386<br>(38.4%) | 154<br>(30.8%) | 232<br>(45.8%) | 178<br>(39.0%) | 203<br>(37.7%) | 176<br>(38.9%) | 210<br>(38.0%) | 233<br>(35.1%) | 152<br>(45.1%) |
| Extremely motivated | 144<br>(14.3%) | 70<br>(14.0%) | 74<br>(14.6%) | 58<br>(12.7%) | 86<br>(16.0%) | 57<br>(12.6%) | 87<br>(15.7%) | 94<br>(14.2%) | 50<br>(14.8%) |

| Have you ever tracked your activity levels or exercise? |  |  |  |  |  |  |  |  |  |
| --- | --- | --- | --- | --- | --- | --- | --- | --- | --- |
|  | 0.717 |  |  | 0.537 |  |  | 0.258 |  | 0.393 |
| No | 96<br>(9.5%) | 64<br>(12.8%) | 32<br>(6.3%) | 45<br>(9.9%) | 50<br>(9.3%) | 38<br>(8.4%) | 58<br>(10.5%) | 69<br>(10.4%) | 27<br>(8.0%) |
| Yes, but only once or rarely<br>(a few times a year) | 124<br>(12.3%) | 60<br>(12.0%) | 64<br>(12.6%) | 50<br>(11.0%) | 72<br>(13.4%) | 67<br>(14.8%) | 57<br>(10.3%) | 88<br>(13.3%) | 36<br>(10.7%) |
| Yes, occasionally (a few<br>times per month) | 295<br>(29.3%) | 122<br>(24.4%) | 173<br>(34.2%) | 132<br>(28.9%) | 158<br>(29.3%) | 139<br>(30.7%) | 156<br>(28.2%) | 187<br>(28.2%) | 105<br>(31.2%) |
| Yes, regularly (1-6 times<br>per week) | 314<br>(31.2%) | 152<br>(30.4%) | 162<br>(32.0%) | 147<br>(32.2%) | 165<br>(30.6%) | 132<br>(29.1%) | 182<br>(32.9%) | 200<br>(30.1%) | 112<br>(33.2%) |
| Yes, daily | 177<br>(17.6%) | 120<br>(20.4%) | 75<br>(14.8%) | 82<br>(18.0%) | 94<br>(17.4%) | 77<br>(17.0%) | 100<br>(18.1%) | 120<br>(18.1%) | 57<br>(16.9%) |

| If yes, what method do/did you primarily use to track your activity? |  |  |  |  |  |  |  |  |  |
| --- | --- | --- | --- | --- | --- | --- | --- | --- | --- |
|  | 0.398 |  |  | 0.966 |  |  | 0.005 |  | 0.659 |
| Smartwatch (e.g., Fitbit,<br>Apple Watch, Garmin) | 428<br>(47.0%) | 218<br>(50.0%) | 210<br>(44.3%) | 196<br>(47.7%) | 228<br>(46.6%) | 183<br>(44.1%) | 245<br>(49.5%) | 278<br>(46.7%) | 149<br>(48.1%) |
| Smartphone app (e.g.,<br>Strava, Google Fit,<br>Samsung Health) | 316<br>(34.7%) | 140<br>(32.1%) | 176<br>(37.1%) | 143<br>(34.8%) | 172<br>(35.2%) | 170<br>(41.0%) | 146<br>(29.5%) | 202<br>(33.9%) | 112<br>(36.1%) |
| Writing it down in a diary or journal | 97<br>(10.7%) | 48<br>(11.0%) | 49<br>(10.3%) | 40<br>(9.7%) | 54<br>(11.0%) | 38<br>(9.2%) | 59<br>(11.9%) | 68<br>(11.4%) | 28<br>(9.0%) |
| Wearable activity tracker (e.g., ring, armband or clip-on device) | 59<br>(6.5%) | 25<br>(5.7%) | 34<br>(7.2%) | 27<br>(2.6%) | 30<br>(6.1%) | 20<br>(4.8%) | 39<br>(7.9%) | 39<br>(6.6%) | 19<br>(6.1%) |
| Other | 10<br>(1.1%) | 5 (1.1%) | 5<br>(1.1%) | 5<br>(1.2%) | 5<br>(1.0%) | 4 (1.0%) | 6<br>(1.2%) | 8 (1.3%) | 2 (0.6%) |
| **What is/was the main reason for tracking your activity?** |  |  |  |  |  |  |  |  |  |
|  |  |  | <0.001 |  | <0.001 |  |  | 0.202 | 0.270 |
| Improving overall health | 322<br>(35.4%) | 125<br>(28.7%) | 197<br>(41.6%) | 159<br>(38.7%) | 160<br>(32.7%) | 140<br>(33.7%) | 182<br>(36.8%) | 199<br>(33.4%) | 119<br>(38.4%) |
| Managing weight | 248<br>(27.3%) | 108<br>(24.8%) | 140<br>(29.5%) | 91<br>(22.1%) | 156<br>(31.9%) | 127<br>(30.6%) | 121<br>(24.4%) | 161<br>(27.1%) | 87<br>(28.1%) |
| Tracking sports or fitness goals | 151<br>(16.6%) | 92<br>(21.1%) | 59<br>(12.4%) | 80<br>(19.5%) | 67<br>(13.7%) | 68<br>(16.4%) | 83<br>(16.8%) | 106<br>(17.8%) | 45<br>(14.5%) |
| To help achieve daily step goals | 95<br>(10.4%) | 46<br>(10.6%) | 49<br>(10.3%) | 31<br>(7.5%) | 63<br>(12.9%) | 41<br>(9.9%) | 54<br>(10.9%) | 60<br>(10.1%) | 35<br>(11.3%) |
| Curiosity about activity levels | 68<br>(7.5%) | 52<br>(11.9%) | 16<br>(3.4%) | 37<br>(9.0%) | 30<br>(6.1%) | 24<br>(5.8%) | 44<br>(8.9%) | 50<br>(8.4%) | 17<br>(5.5%) |
| Managing a medical condition (e.g., heart health, rehabilitation) | 22<br>(2.4%) | 9 (2.1%) | 13<br>(2.7%) | 11<br>(2.7%) | 11<br>(2.2%) | 13<br>(3.1%) | 9<br>(1.8%) | 15<br>(2.5%) | 7 (2.3%) |
| Other | 4<br>(0.4%) | 4 (0.9%) | 0 (0%) | 2<br>(0.5%) | 2<br>(0.4%) | 2 (0.5%) | 2<br>(0.4%) | 4 (0.7%) | 0 (0%) |

For PA, most participants reported exercising 3-6 times per week. Participants were typically very confident in their ability to track their activity levels and very motivated to do so. Participants typically reported tracking their activity levels 1-6 times per week, usually via a smartwatch. The main reason for tracking activity levels was to improve overall health.

### General study design features

Over 90% participants stated that they were very likely or likely to participate in a study exploring how lifestyle impacts brain health/dementia risk (Table 3). For most participants, none of their social circle participated in research. Remote data collection methods were most acceptable to participants.

**Table 3.**
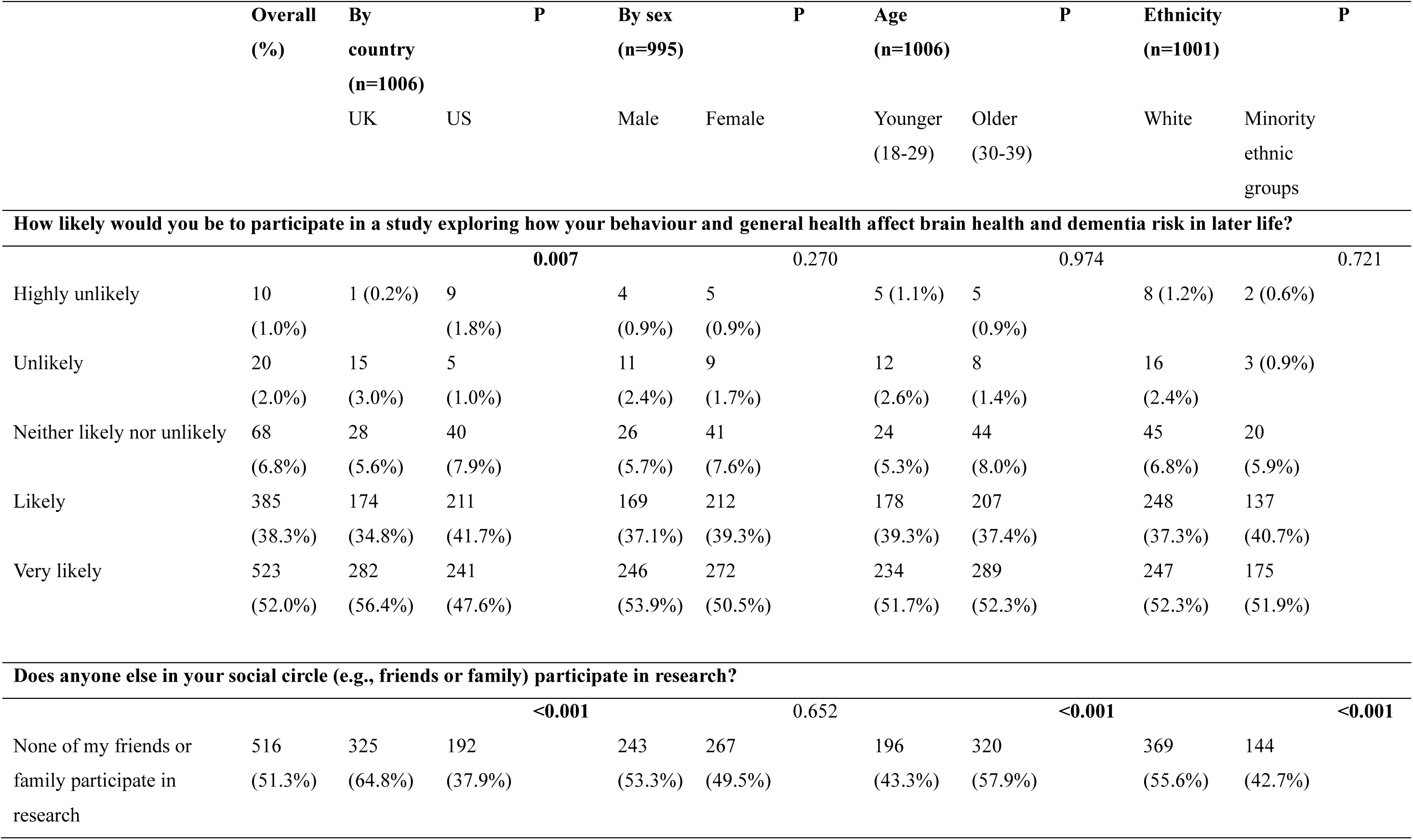
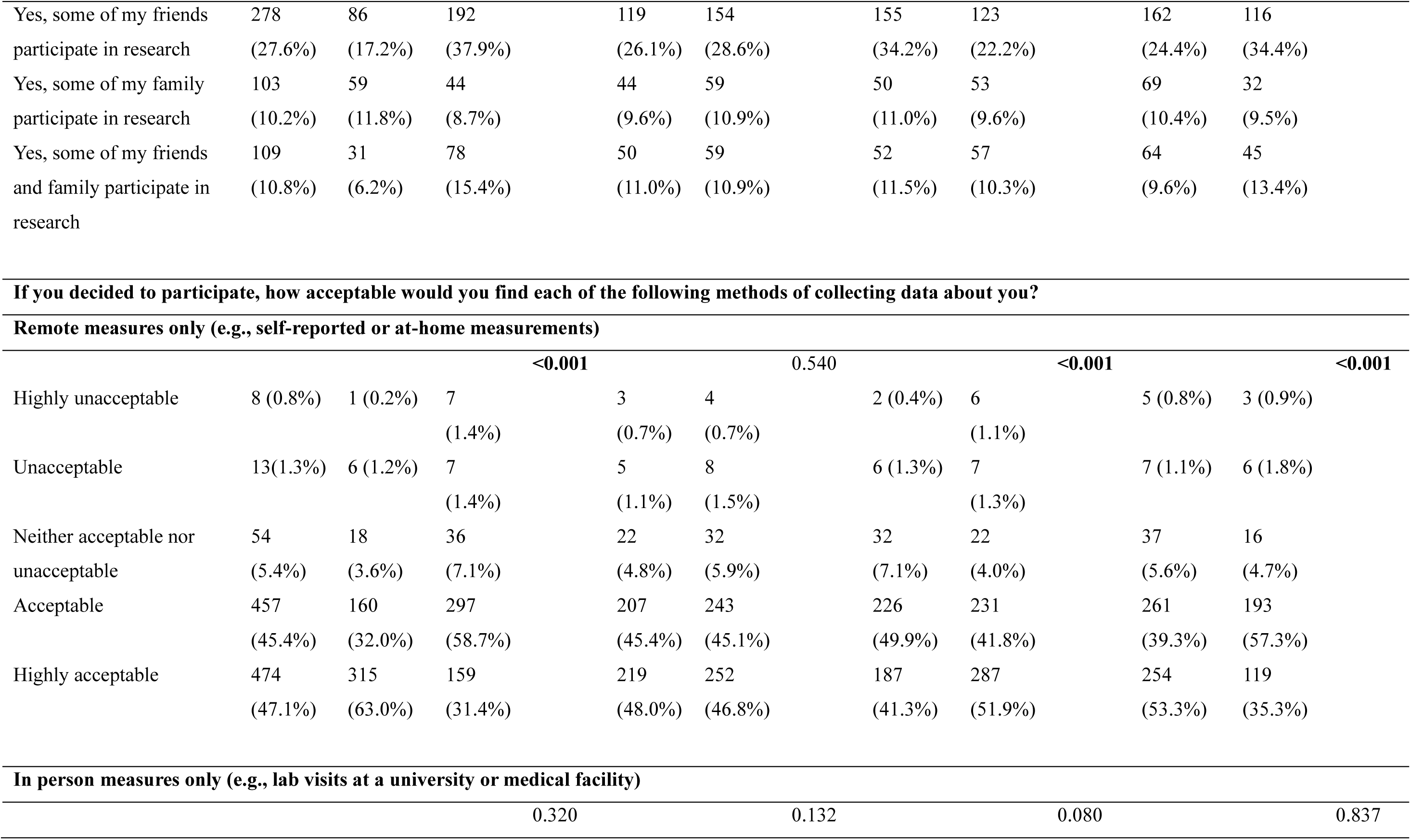
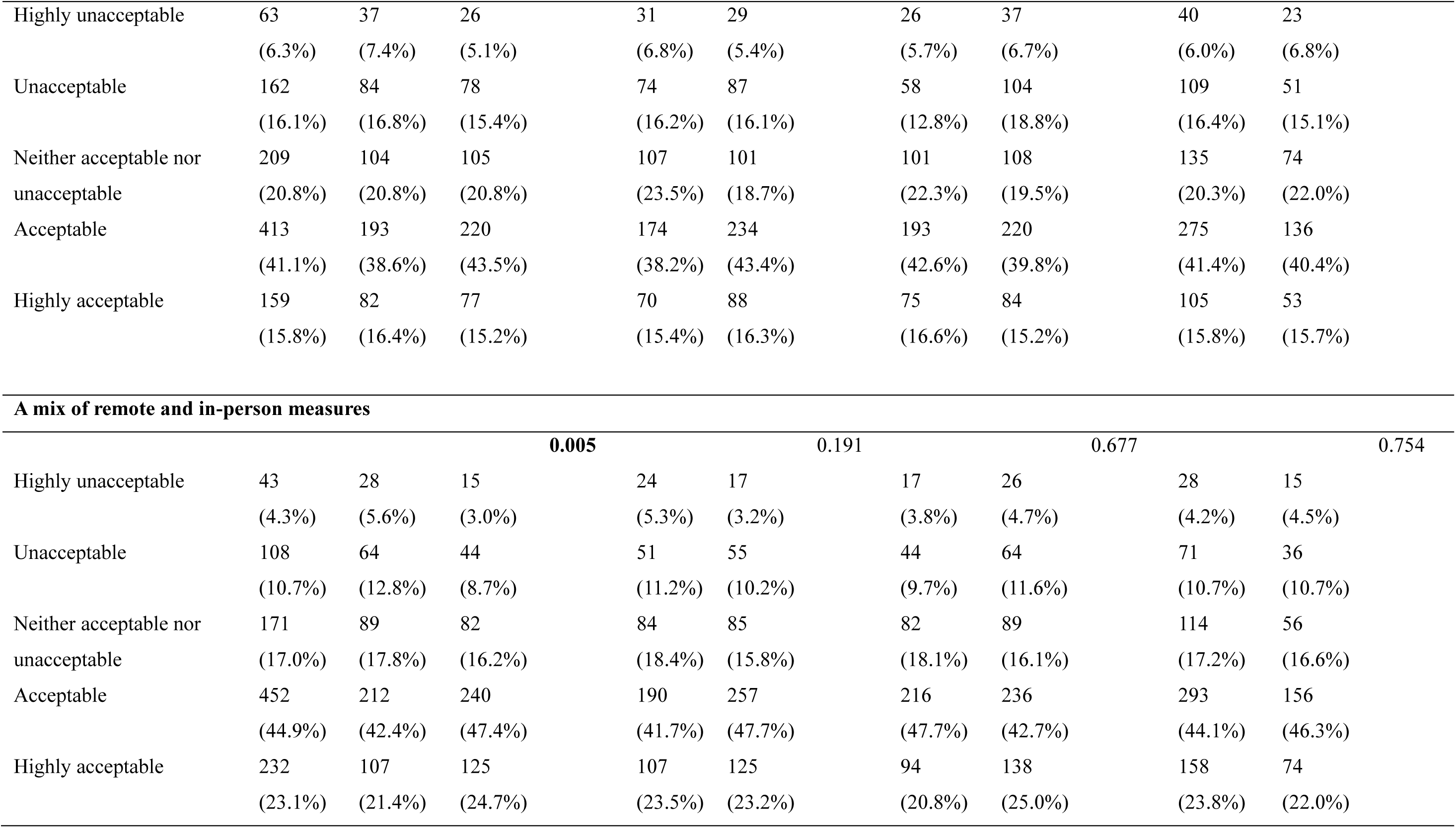
General preferences for study design

|  | Overall<br>(%) | By<br>country<br>(n=1006) |  | P |  | By sex<br>(n=995) |  | P |  | Age<br>(n=1006) |  | P |  | Ethnicity<br>(n=1001) |  | P |
| --- | --- | --- | --- | --- | --- | --- | --- | --- | --- | --- | --- | --- | --- | --- | --- | --- |
|  |  | UK | US |  |  | Male | Female |  |  | Younger<br>(18-29) | Older<br>(30-39) |  |  | White | Minority<br>ethnic<br>groups |  |
| **How likely would you be to participate in a study exploring how your behaviour and general health affect brain health and dementia risk in later life?** |  |  |  |  |  |  |  |  |  |  |  |  |  |  |  |  |
|  |  |  |  | **0.007** |  |  |  | 0.270 |  |  |  | 0.974 |  |  |  | 0.721 |
| Highly unlikely | 10<br>(1.0%) | 1 (0.2%) | 9<br>(1.8%) |  |  | 4<br>(0.9%) | 5<br>(0.9%) |  |  | 5 (1.1%) | 5<br>(0.9%) |  |  | 8 (1.2%) | 2 (0.6%) |  |
| Unlikely | 20<br>(2.0%) | 15<br>(3.0%) | 5<br>(1.0%) |  |  | 11<br>(2.4%) | 9<br>(1.7%) |  |  | 12<br>(2.6%) | 8<br>(1.4%) |  |  | 16<br>(2.4%) | 3 (0.9%) |  |
| Neither likely nor unlikely | 68<br>(6.8%) | 28<br>(5.6%) | 40<br>(7.9%) |  |  | 26<br>(5.7%) | 41<br>(7.6%) |  |  | 24<br>(5.3%) | 44<br>(8.0%) |  |  | 45<br>(6.8%) | 20<br>(5.9%) |  |
| Likely | 385<br>(38.3%) | 174<br>(34.8%) | 211<br>(41.7%) |  |  | 169<br>(37.1%) | 212<br>(39.3%) |  |  | 178<br>(39.3%) | 207<br>(37.4%) |  |  | 248<br>(37.3%) | 137<br>(40.7%) |  |
| Very likely | 523<br>(52.0%) | 282<br>(56.4%) | 241<br>(47.6%) |  |  | 246<br>(53.9%) | 272<br>(50.5%) |  |  | 234<br>(51.7%) | 289<br>(52.3%) |  |  | 247<br>(52.3%) | 175<br>(51.9%) |  |
| **Does anyone else in your social circle (e.g., friends or family) participate in research?** |  |  |  |  |  |  |  |  |  |  |  |  |  |  |  |  |
|  |  |  |  | **<0.001** |  |  |  | 0.652 |  |  |  | **<0.001** |  |  |  | **<0.001** |
| None of my friends or<br>family participate in<br>research | 516<br>(51.3%) | 325<br>(64.8%) | 192<br>(37.9%) |  |  | 243<br>(53.3%) | 267<br>(49.5%) |  |  | 196<br>(43.3%) | 320<br>(57.9%) |  |  | 369<br>(55.6%) | 144<br>(42.7%) |  |

|  |  |  |  |  |  |  |  |  |  |
| --- | --- | --- | --- | --- | --- | --- | --- | --- | --- |
| Yes, some of my friends<br>participate in research | 278<br>(27.6%) | 86<br>(17.2%) | 192<br>(37.9%) | 119<br>(26.1%) | 154<br>(28.6%) | 155<br>(34.2%) | 123<br>(22.2%) | 162<br>(24.4%) | 116<br>(34.4%) |
| Yes, some of my family<br>participate in research | 103<br>(10.2%) | 59<br>(11.8%) | 44<br>(8.7%) | 44<br>(9.6%) | 59<br>(10.9%) | 50<br>(11.0%) | 53<br>(9.6%) | 69<br>(10.4%) | 32<br>(9.5%) |
| Yes, some of my friends<br>and family participate in<br>research | 109<br>(10.8%) | 31<br>(6.2%) | 78<br>(15.4%) | 50<br>(11.0%) | 59<br>(10.9%) | 52<br>(11.5%) | 57<br>(10.3%) | 64<br>(9.6%) | 45<br>(13.4%) |

|  |  |  | <0.001 |  | 0.540 |  | <0.001 |  | <0.001 |
| --- | --- | --- | --- | --- | --- | --- | --- | --- | --- |
| Highly unacceptable | 8 (0.8%) | 1 (0.2%) | 7<br>(1.4%) | 3<br>(0.7%) | 4<br>(0.7%) | 2 (0.4%) | 6<br>(1.1%) | 5 (0.8%) | 3 (0.9%) |
| Unacceptable | 13(1.3%) | 6 (1.2%) | 7<br>(1.4%) | 5<br>(1.1%) | 8<br>(1.5%) | 6 (1.3%) | 7<br>(1.3%) | 7 (1.1%) | 6 (1.8%) |
| Neither acceptable nor<br>unacceptable | 54<br>(5.4%) | 18<br>(3.6%) | 36<br>(7.1%) | 22<br>(4.8%) | 32<br>(5.9%) | 32<br>(7.1%) | 22<br>(4.0%) | 37<br>(5.6%) | 16<br>(4.7%) |
| Acceptable | 457<br>(45.4%) | 160<br>(32.0%) | 297<br>(58.7%) | 207<br>(45.4%) | 243<br>(45.1%) | 226<br>(49.9%) | 231<br>(41.8%) | 261<br>(39.3%) | 193<br>(57.3%) |
| Highly acceptable | 474<br>(47.1%) | 315<br>(63.0%) | 159<br>(31.4%) | 219<br>(48.0%) | 252<br>(46.8%) | 187<br>(41.3%) | 287<br>(51.9%) | 254<br>(53.3%) | 119<br>(35.3%) |

**In person measures only (e.g., lab visits at a university or medical facility)**
|  |  |  |  |  |  |  |  |  |  |  |
| --- | --- | --- | --- | --- | --- | --- | --- | --- | --- | --- |
|  |  |  |  | 0.320 |  | 0.132 |  | 0.080 |  | 0.837 |

|  |  |  |  |  |  |  |  |  |  |
| --- | --- | --- | --- | --- | --- | --- | --- | --- | --- |
| Highly unacceptable | 63 | 37 | 26 | 31 | 29 | 26 | 37 | 40 | 23 |
|  | (6.3%) | (7.4%) | (5.1%) | (6.8%) | (5.4%) | (5.7%) | (6.7%) | (6.0%) | (6.8%) |
| Unacceptable | 162 | 84 | 78 | 74 | 87 | 58 | 104 | 109 | 51 |
|  | (16.1%) | (16.8%) | (15.4%) | (16.2%) | (16.1%) | (12.8%) | (18.8%) | (16.4%) | (15.1%) |
| Neither acceptable nor unacceptable | 209 | 104 | 105 | 107 | 101 | 101 | 108 | 135 | 74 |
|  | (20.8%) | (20.8%) | (20.8%) | (23.5%) | (18.7%) | (22.3%) | (19.5%) | (20.3%) | (22.0%) |
| Acceptable | 413 | 193 | 220 | 174 | 234 | 193 | 220 | 275 | 136 |
|  | (41.1%) | (38.6%) | (43.5%) | (38.2%) | (43.4%) | (42.6%) | (39.8%) | (41.4%) | (40.4%) |
| Highly acceptable | 159 | 82 | 77 | 70 | 88 | 75 | 84 | 105 | 53 |
|  | (15.8%) | (16.4%) | (15.2%) | (15.4%) | (16.3%) | (16.6%) | (15.2%) | (15.8%) | (15.7%) |

| A mix of remote and in-person measures |  |  |  |  |  |  |  |  |  |
| --- | --- | --- | --- | --- | --- | --- | --- | --- | --- |
|  | 0.005 |  |  | 0.191 |  | 0.677 |  | 0.754 |  |
| Highly unacceptable | 43 | 28 | 15 | 24 | 17 | 17 | 26 | 28 | 15 |
|  | (4.3%) | (5.6%) | (3.0%) | (5.3%) | (3.2%) | (3.8%) | (4.7%) | (4.2%) | (4.5%) |
| Unacceptable | 108 | 64 | 44 | 51 | 55 | 44 | 64 | 71 | 36 |
|  | (10.7%) | (12.8%) | (8.7%) | (11.2%) | (10.2%) | (9.7%) | (11.6%) | (10.7%) | (10.7%) |
| Neither acceptable nor unacceptable | 171 | 89 | 82 | 84 | 85 | 82 | 89 | 114 | 56 |
|  | (17.0%) | (17.8%) | (16.2%) | (18.4%) | (15.8%) | (18.1%) | (16.1%) | (17.2%) | (16.6%) |
| Acceptable | 452 | 212 | 240 | 190 | 257 | 216 | 236 | 293 | 156 |
|  | (44.9%) | (42.4%) | (47.4%) | (41.7%) | (47.7%) | (47.7%) | (42.7%) | (44.1%) | (46.3%) |
| Highly acceptable | 232 | 107 | 125 | 107 | 125 | 94 | 138 | 158 | 74 |
|  | (23.1%) | (21.4%) | (24.7%) | (23.5%) | (23.2%) | (20.8%) | (25.0%) | (23.8%) | (22.0%) |

### Dietary assessment: Preferred methods

Food frequency questionnaires, multiple 24-hour dietary recalls, diet tracking apps, and photographing meals were all rated as being either ‘highly acceptable’ or ‘acceptable’ by >80% of participants (Table 4). Diet tracking apps were rated by the highest percentage of participants as highly acceptable. Collection of biological samples for the assessment of dietary intake was generally deemed to be less acceptable. Urine samples showed the highest acceptability among biological methods. Most participants indicated willingness to provide dietary data once or more every year.

**Table 4.**
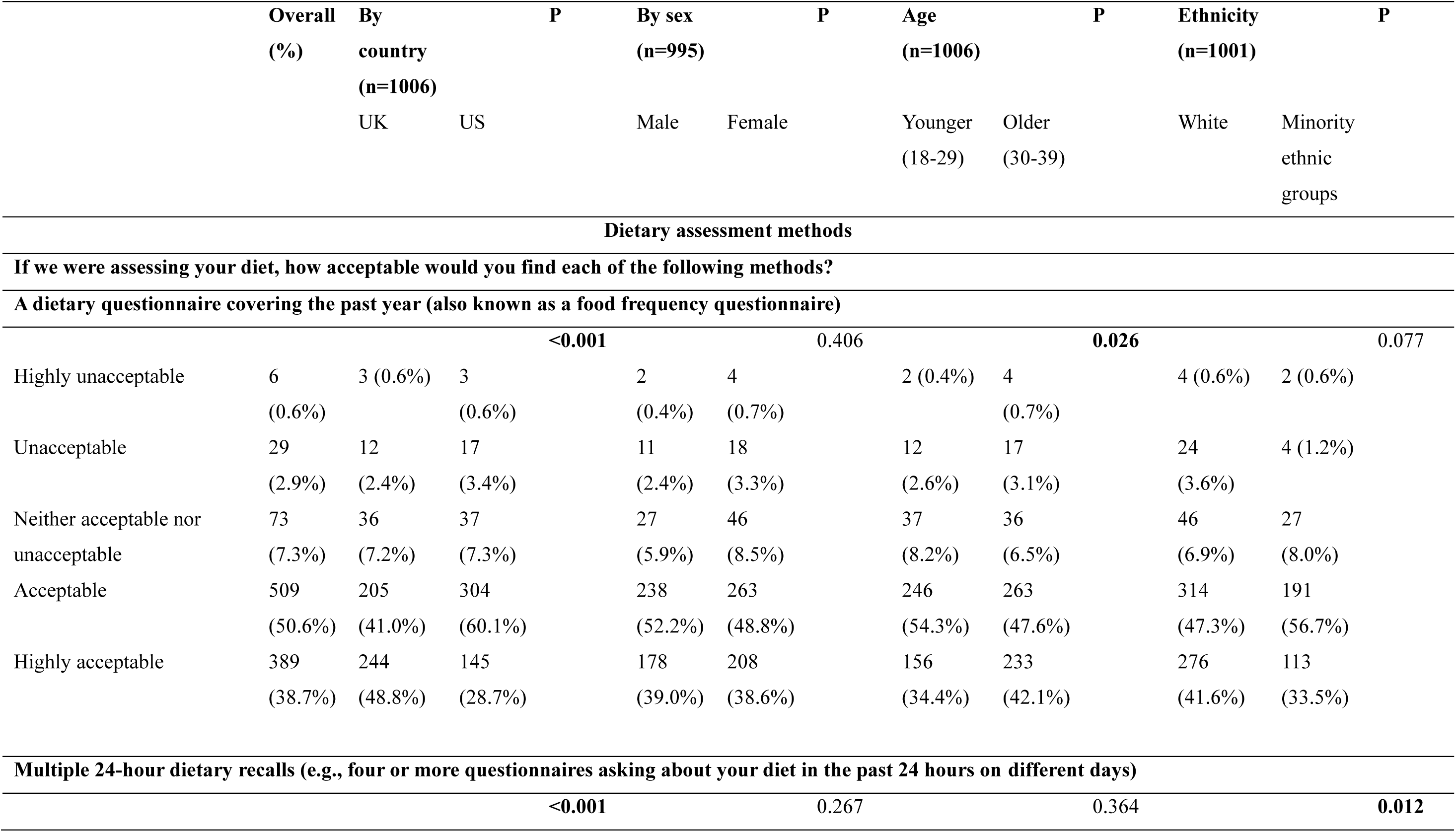
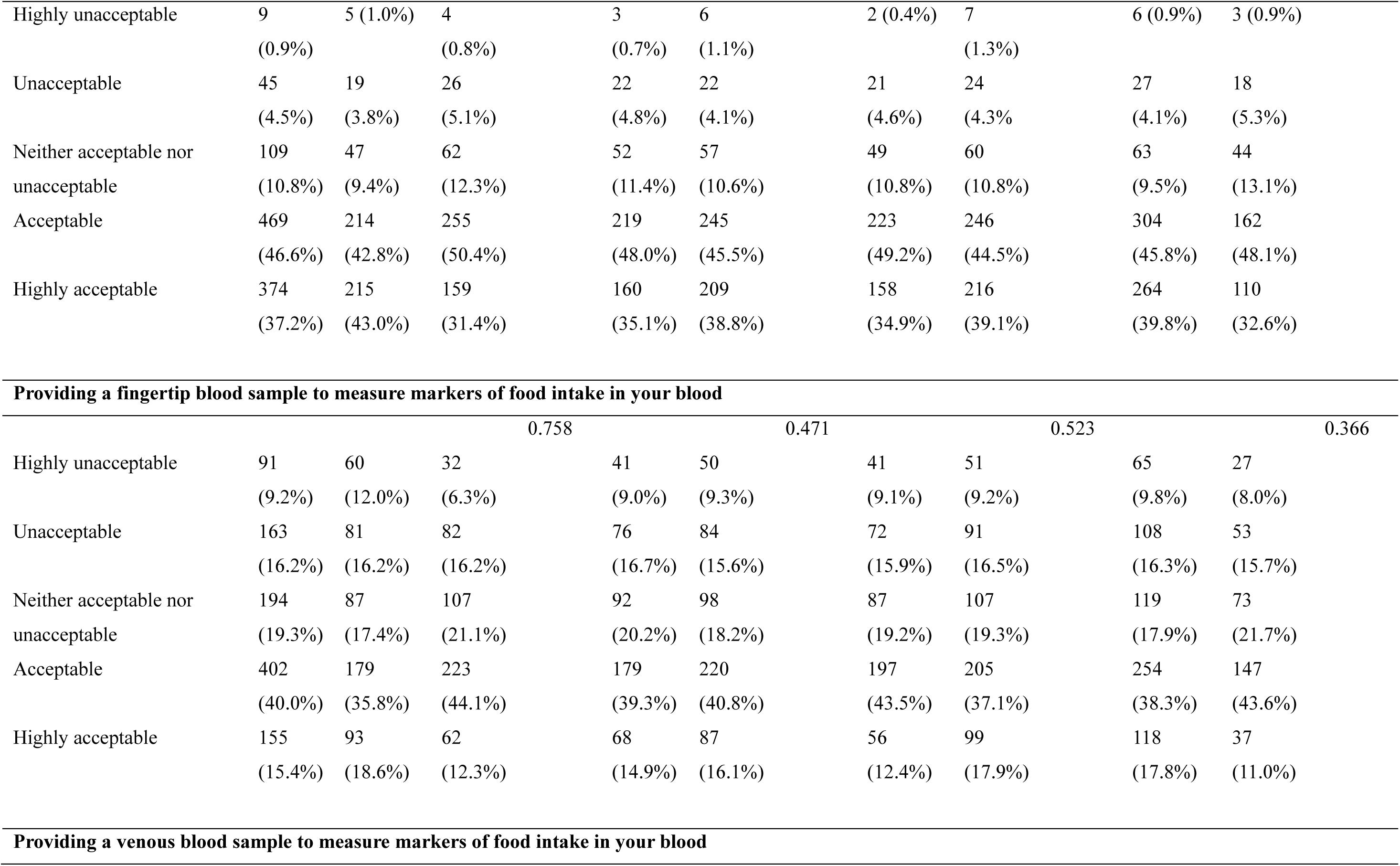
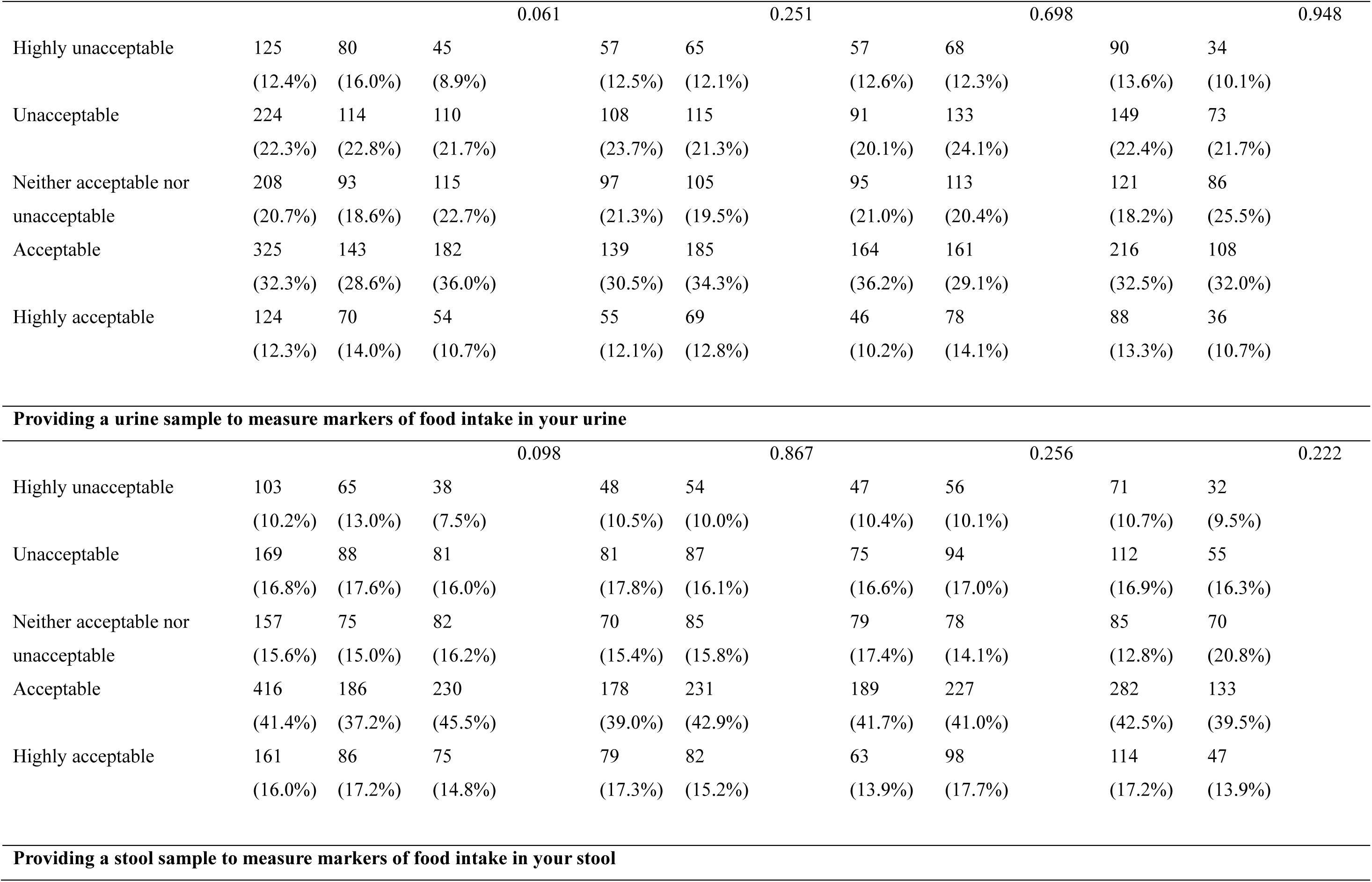
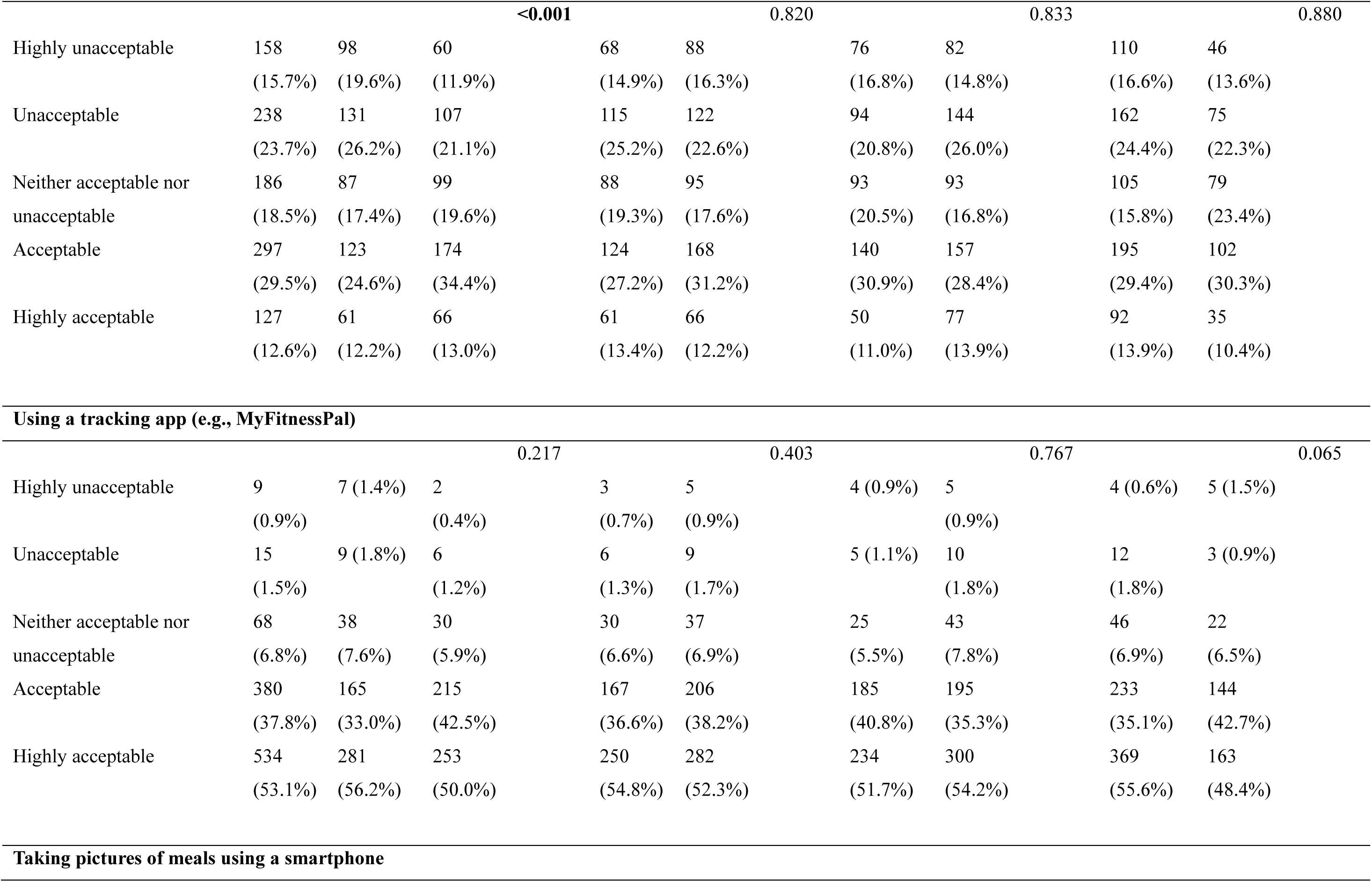
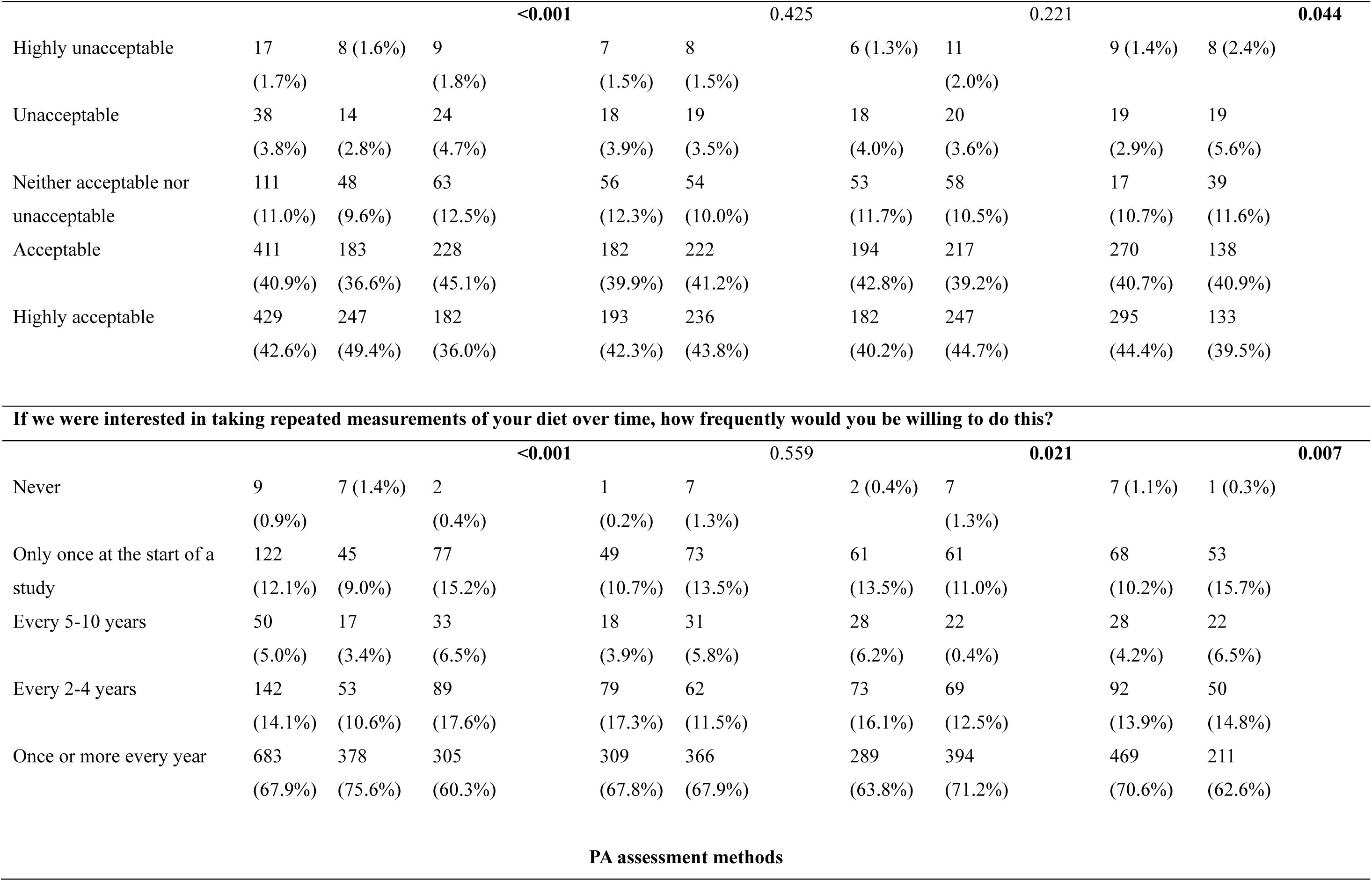
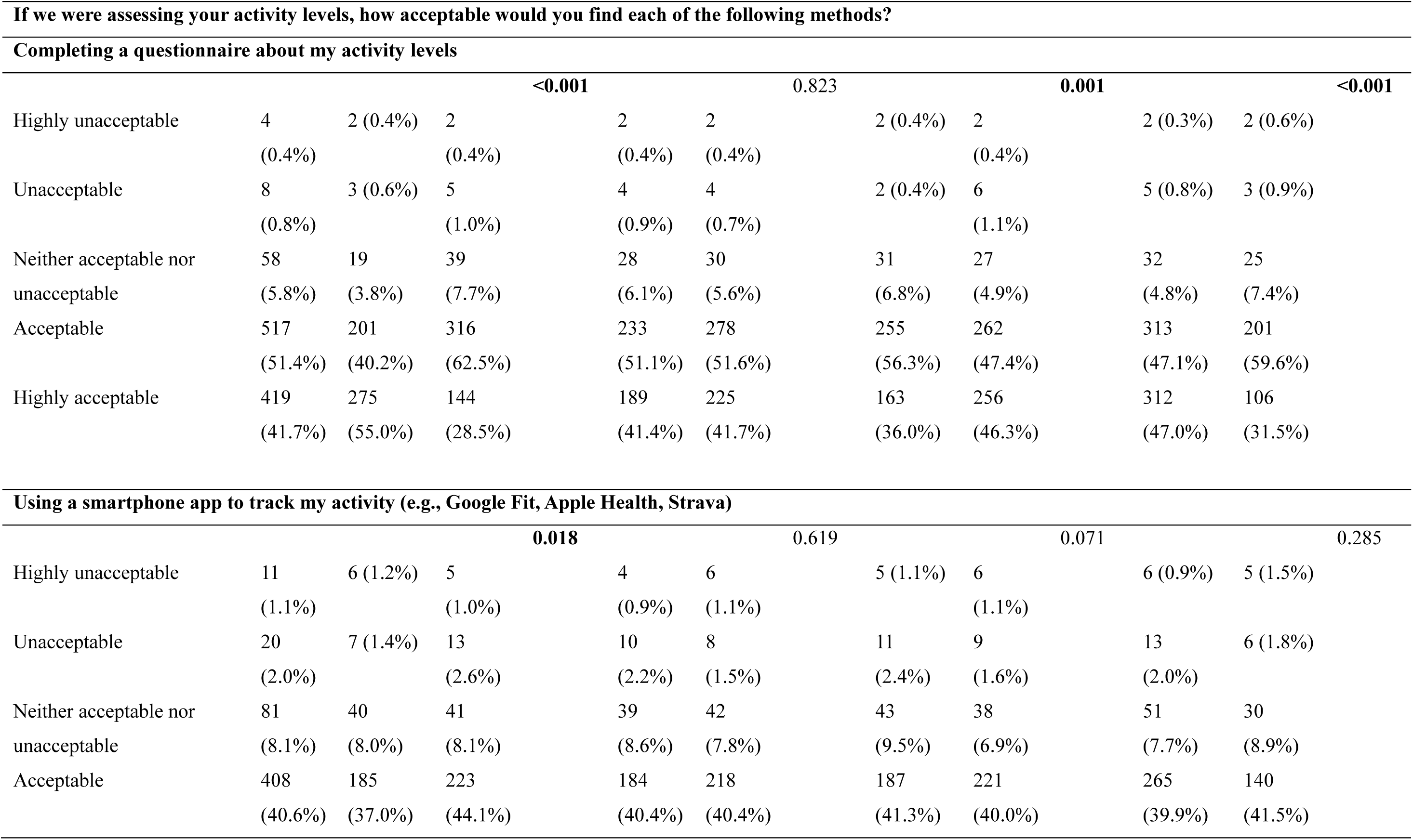
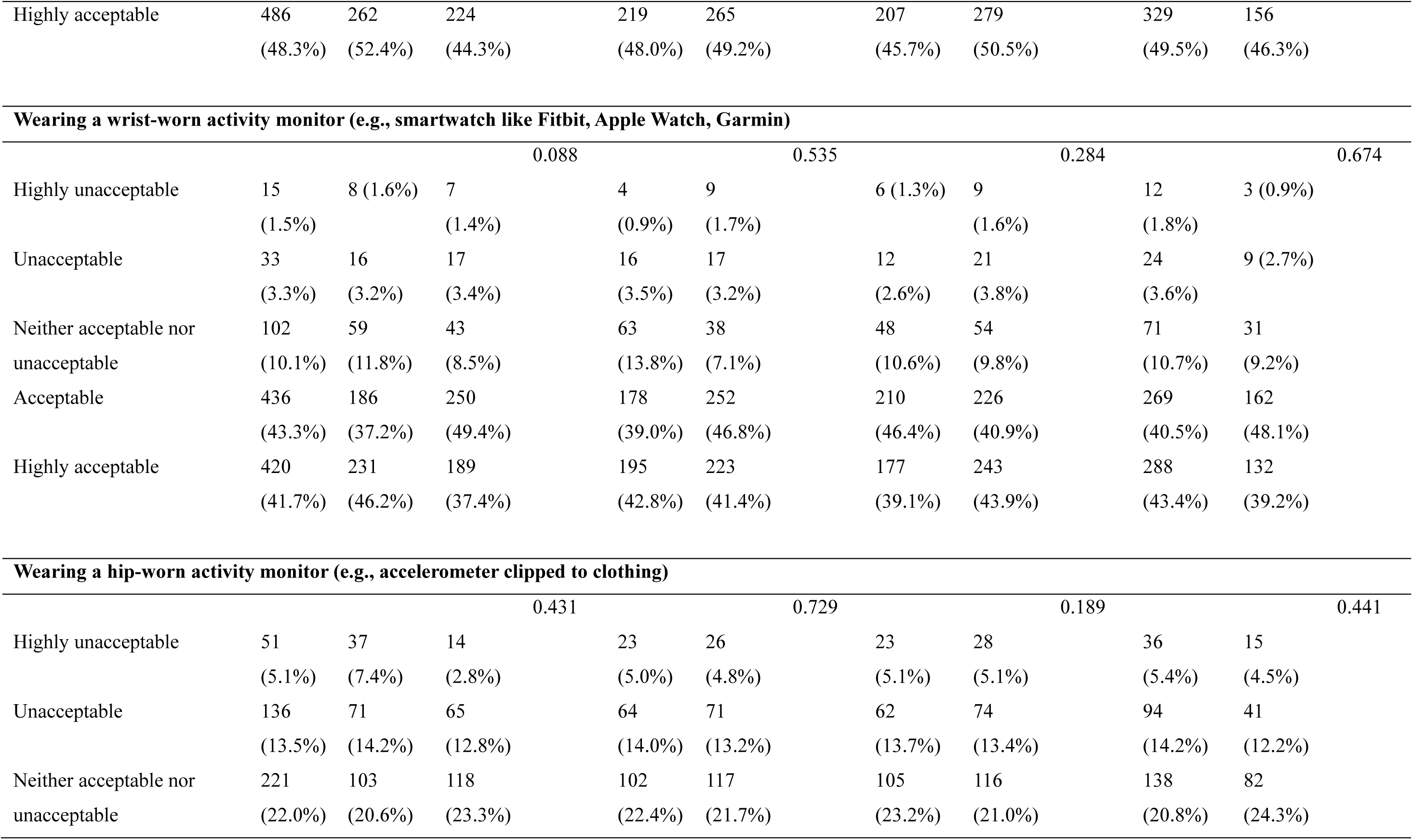
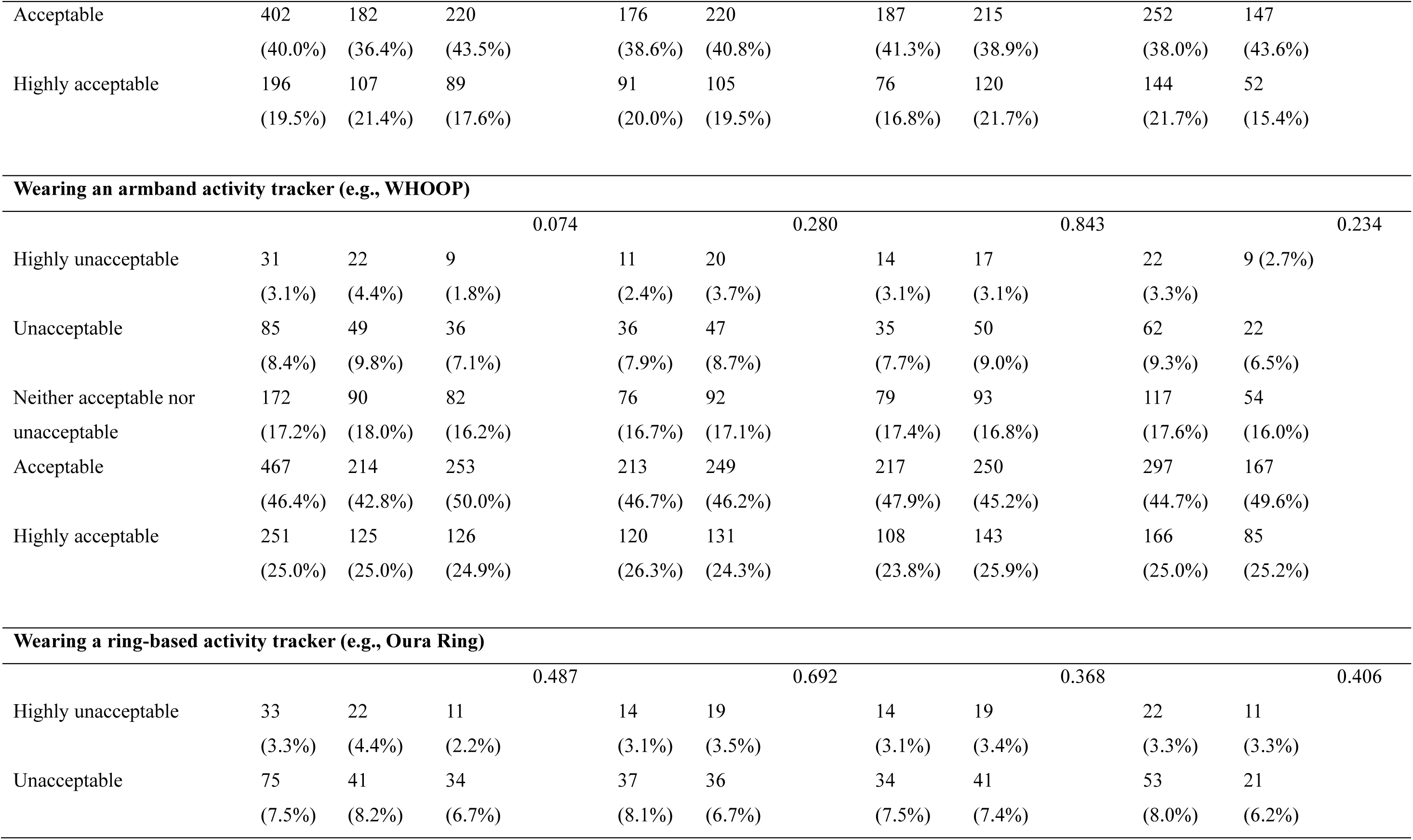
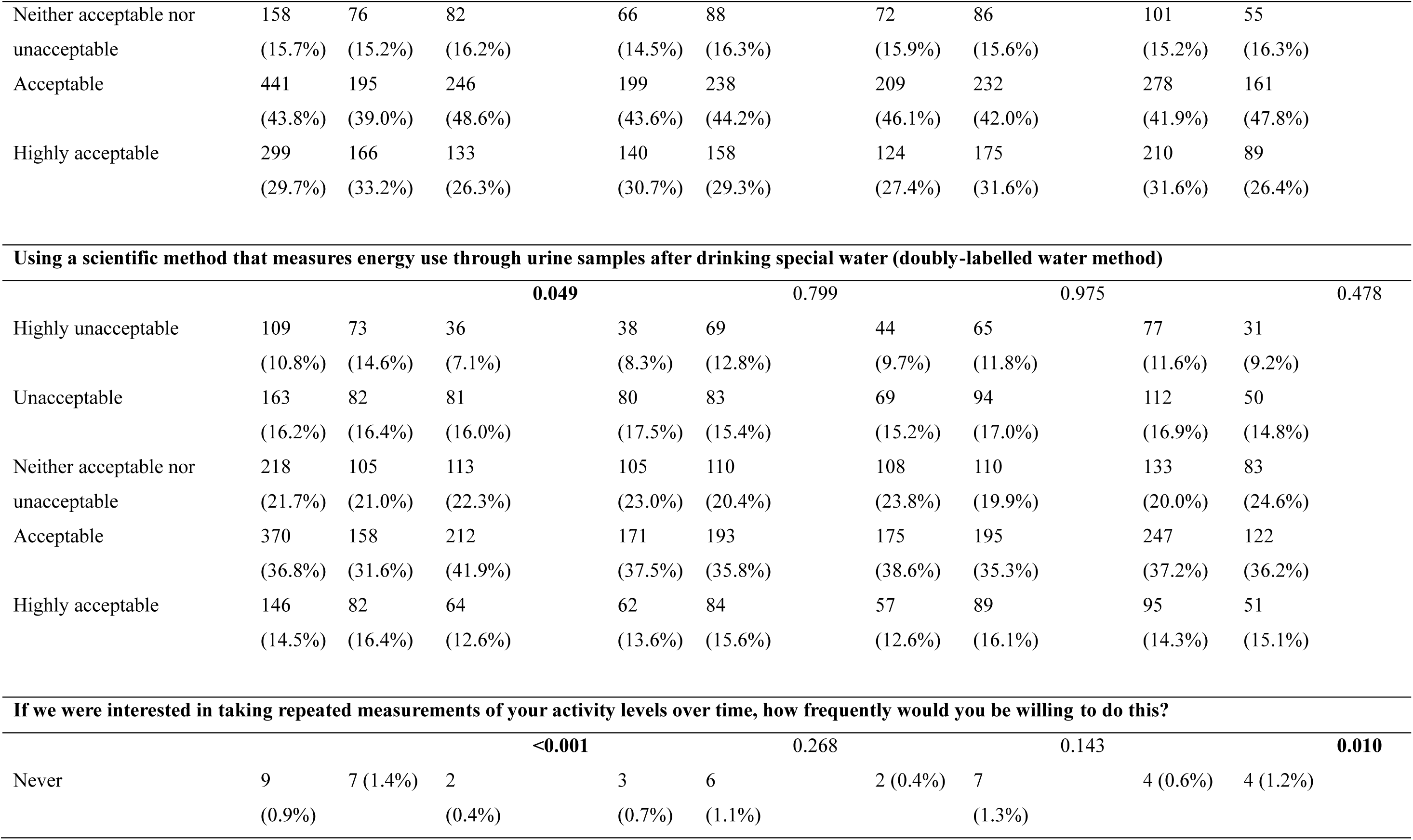
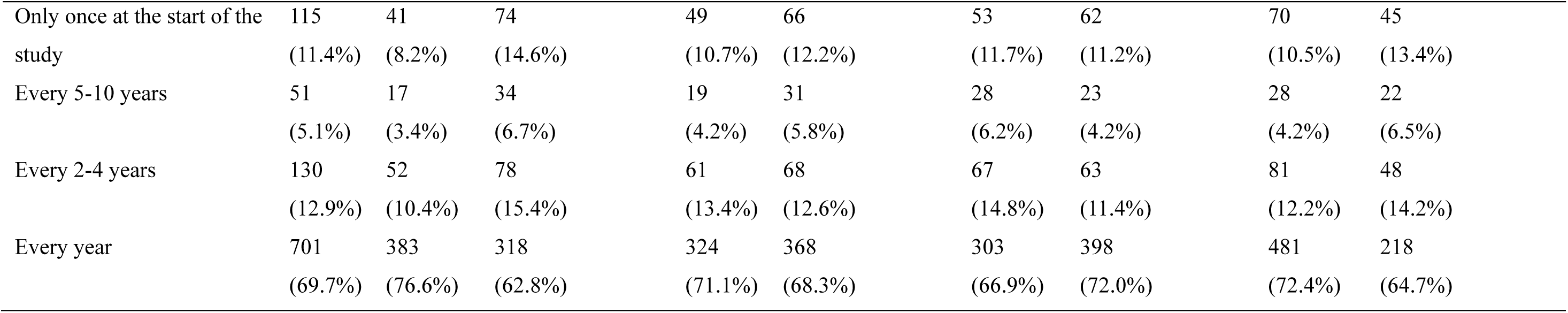
Preferred methods for diet and PA tracking

### Physical activity assessment: Preferred methods

Most PA tracking methods showed high acceptability (Table 4). Completing a questionnaire about activity levels, smartphone apps for activity tracking, and wrist-worn activity monitors were rated as being either ’highly acceptable’ or ’acceptable’ by >80% of participants. Smartphone apps were the most acceptable PA tracking method. The most invasive method, doubly labelled water - a method that measures energy expenditure through the elimination rates of stable isotopes - showed the lowest acceptability. Most participants indicated willingness to provide PA data annually.

### Barriers and enablers to diet tracking

Most barriers were not perceived as major challenges and were typically rated as not at all or slightly challenging (Table 5). The greatest perceived barriers for diet tracking were estimating portion sizes accurately, tracking diet outside of the home, and frequently eating out or getting takeaways. These barriers primarily map onto the TDF domains of ’Knowledge’ and ’Skills’ (e.g., estimating portion sizes) and ’Environmental context and resources’ (e.g., tracking difficulties in non-home settings). The strongest perceived enablers were receiving incentives or rewards, using a smartphone to record everything, and flexibility in when participants can report their diets. These enablers primarily map onto the TDF domains of ’Reinforcement’ (e.g., incentives/rewards) and ’Behavioural regulation’ (e.g., smartphone recording, pictures, flexibility).

**Table 5.**
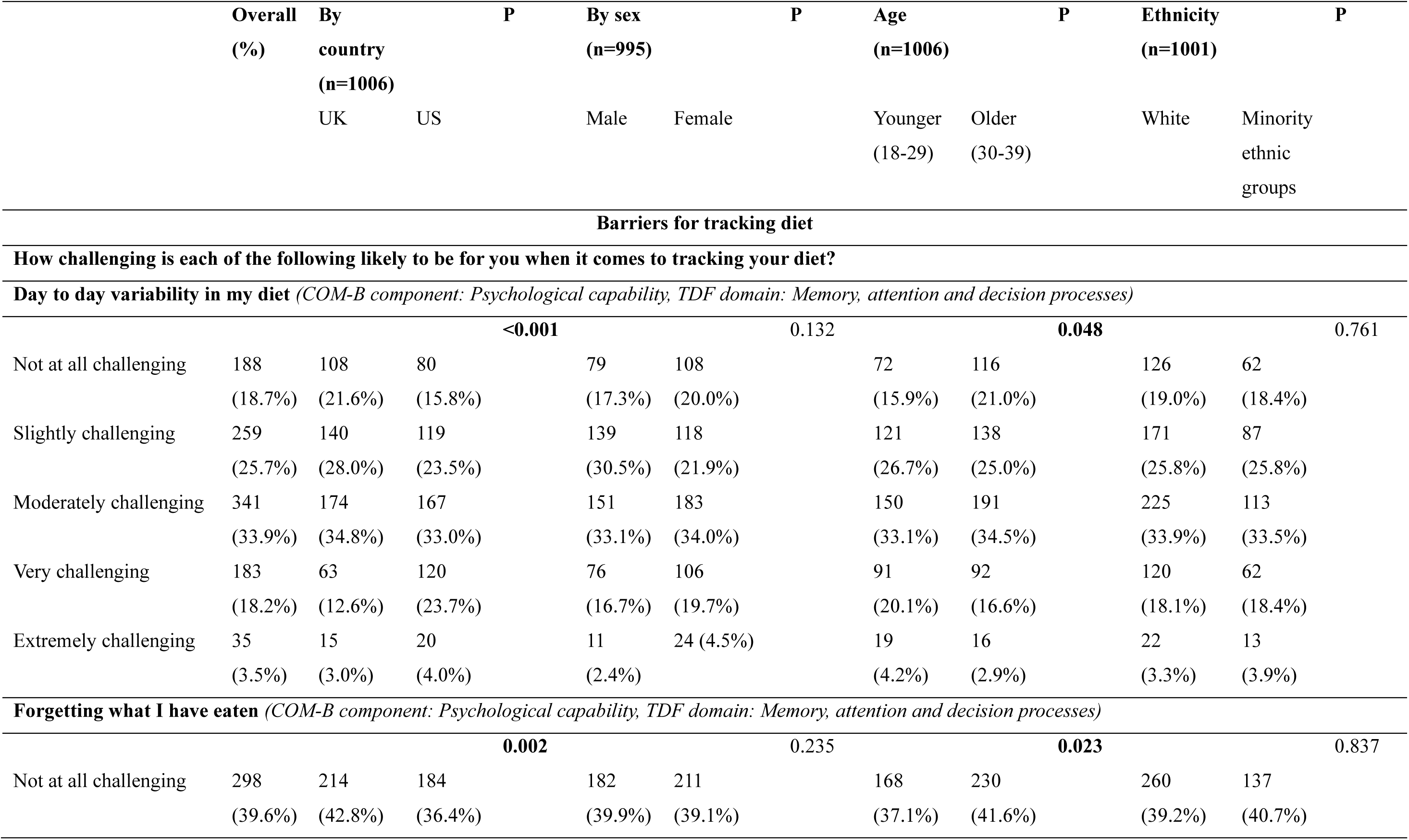
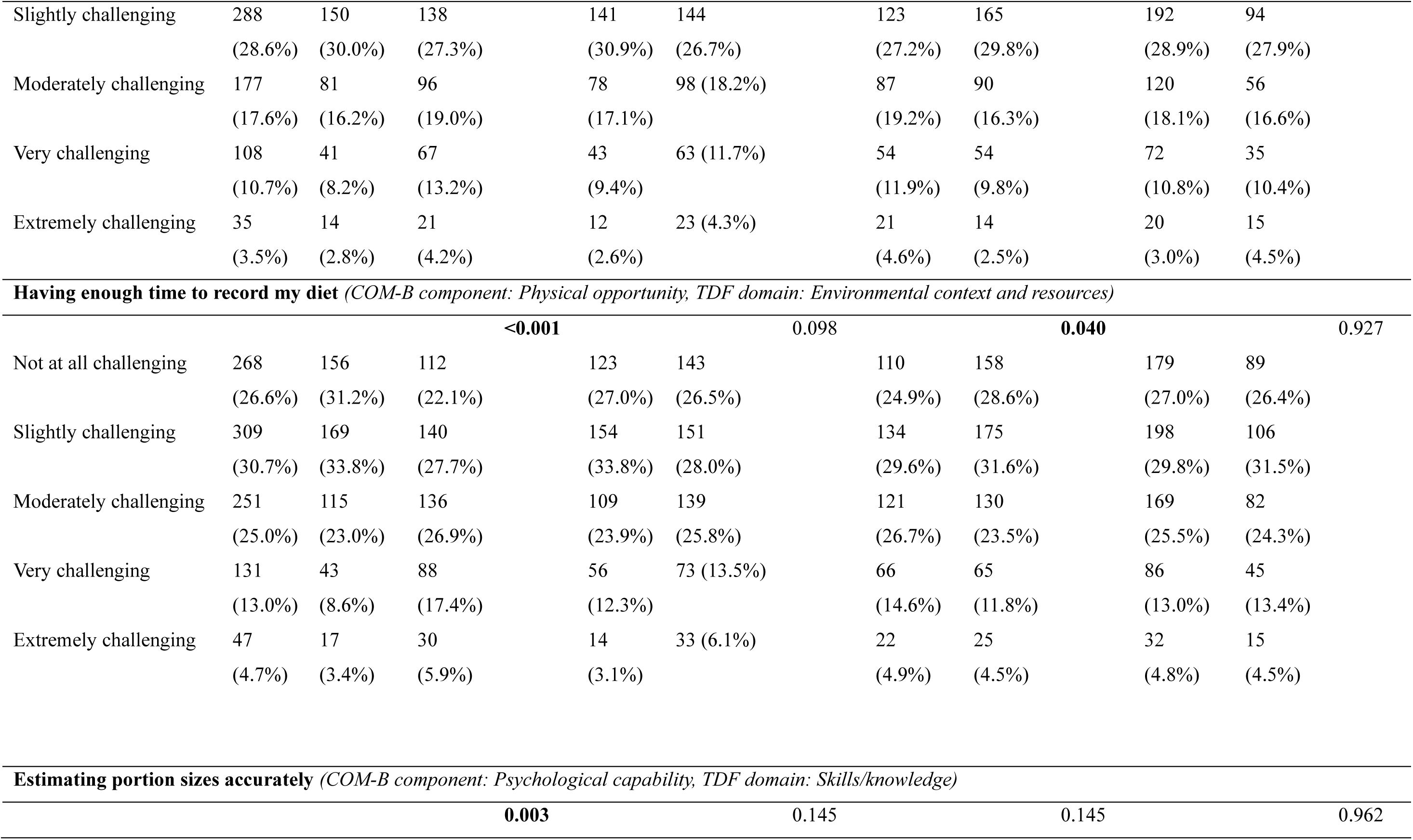
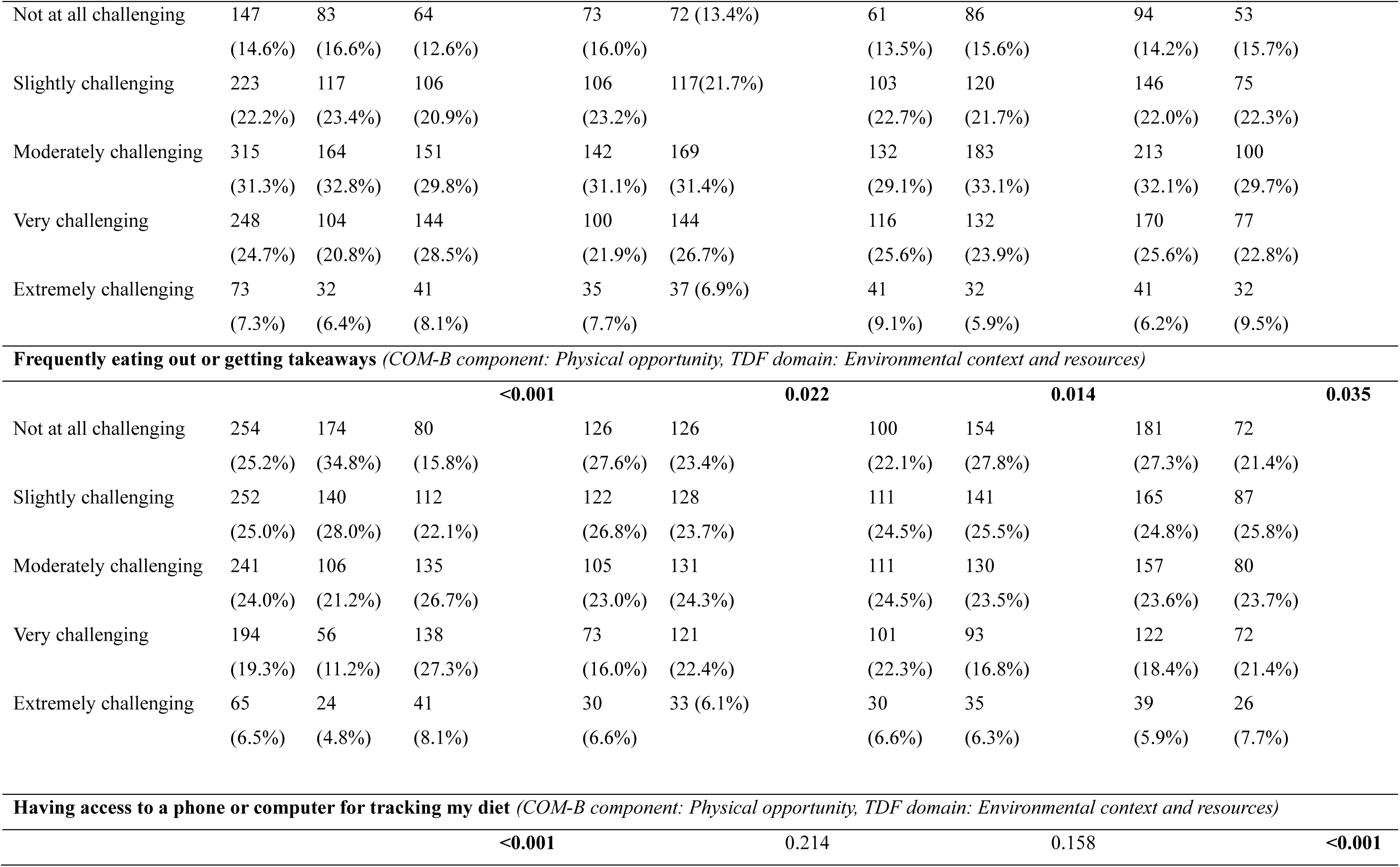
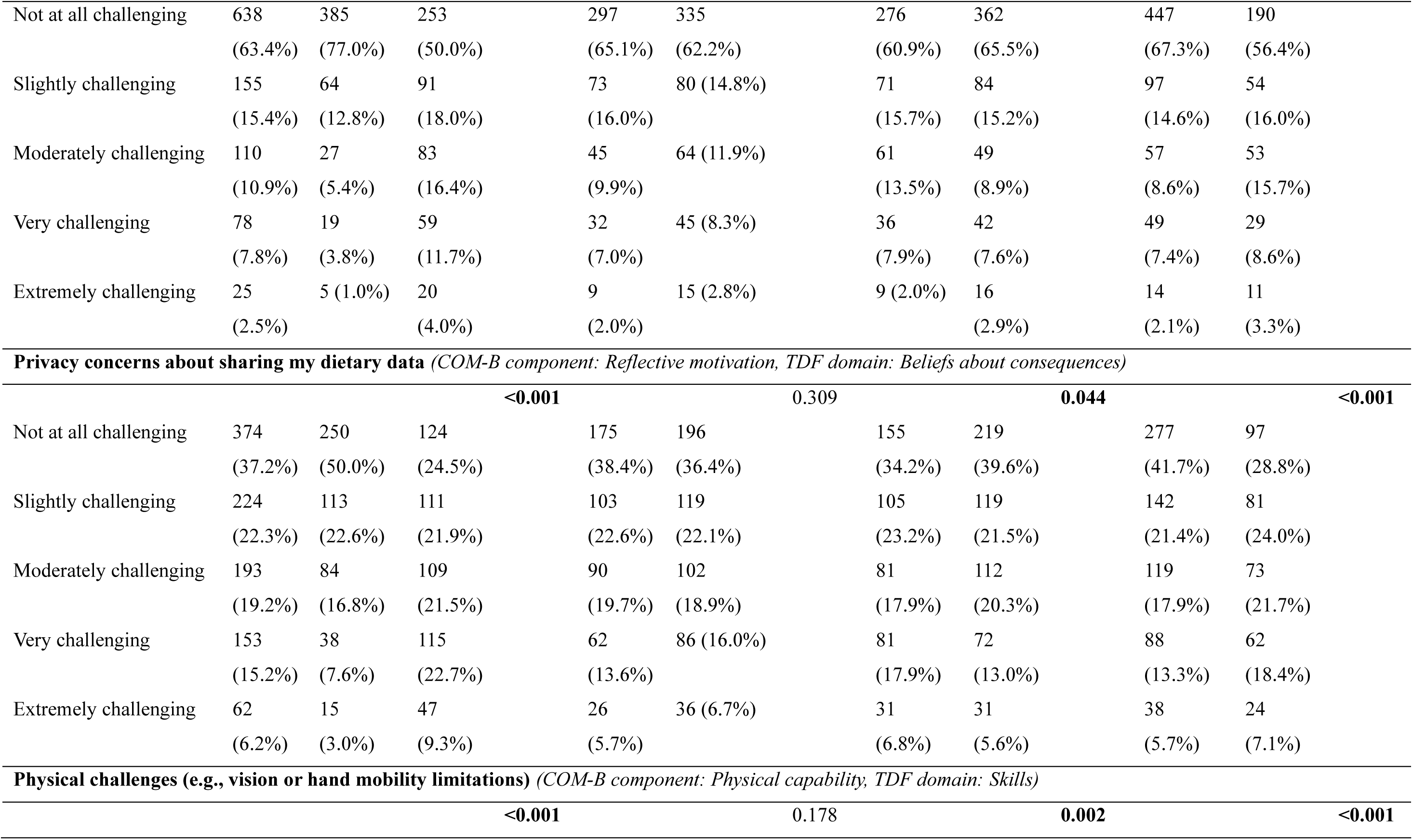
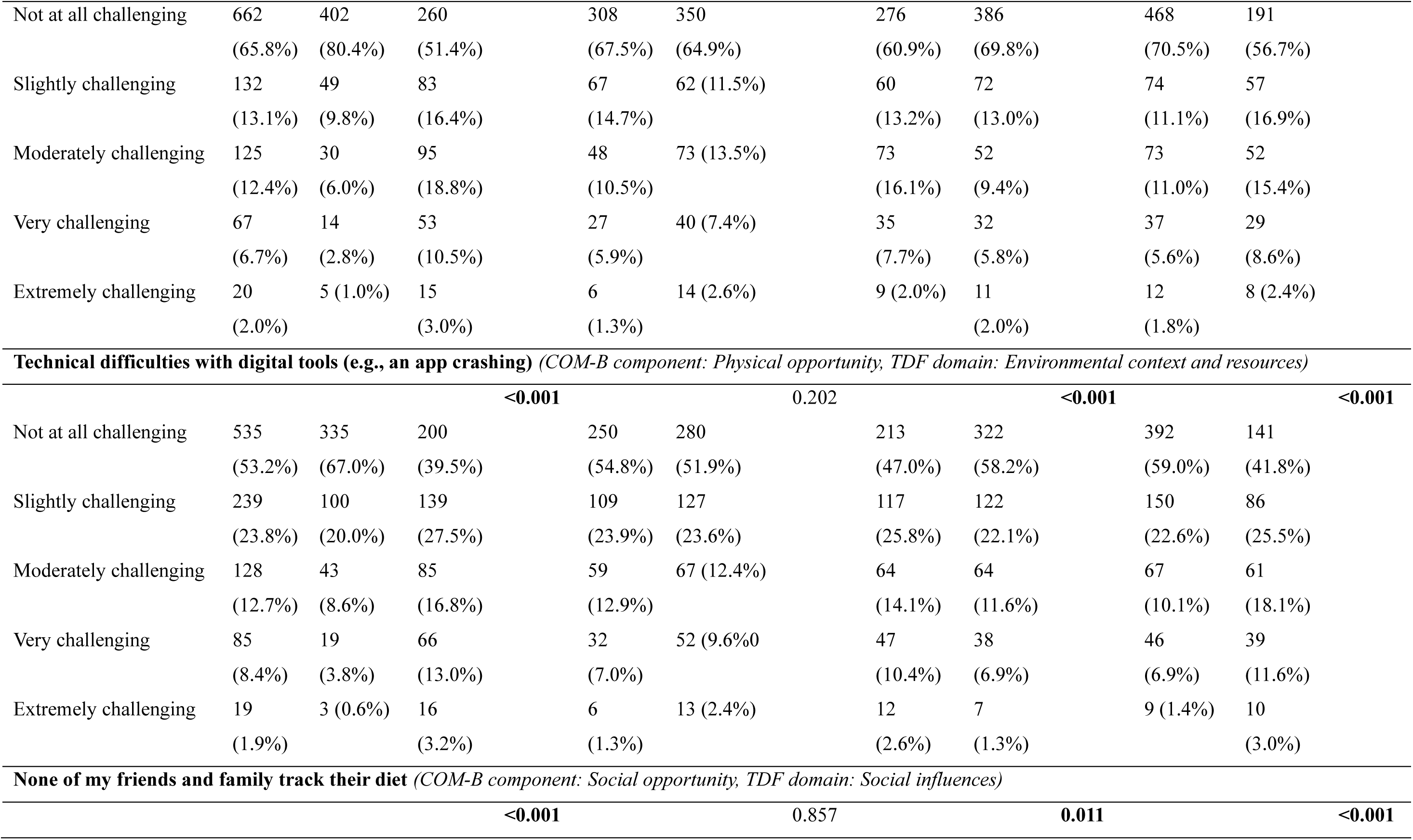
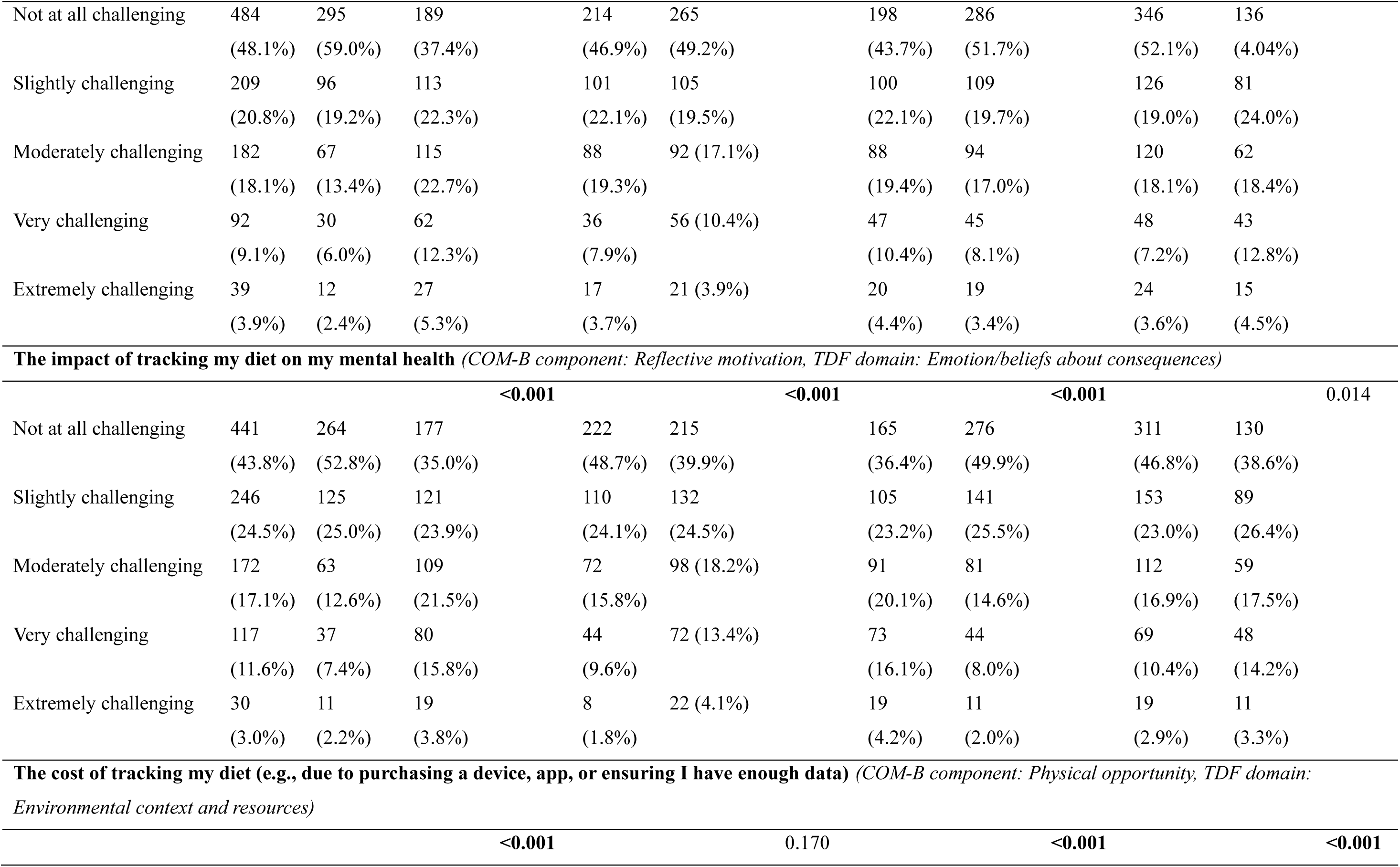
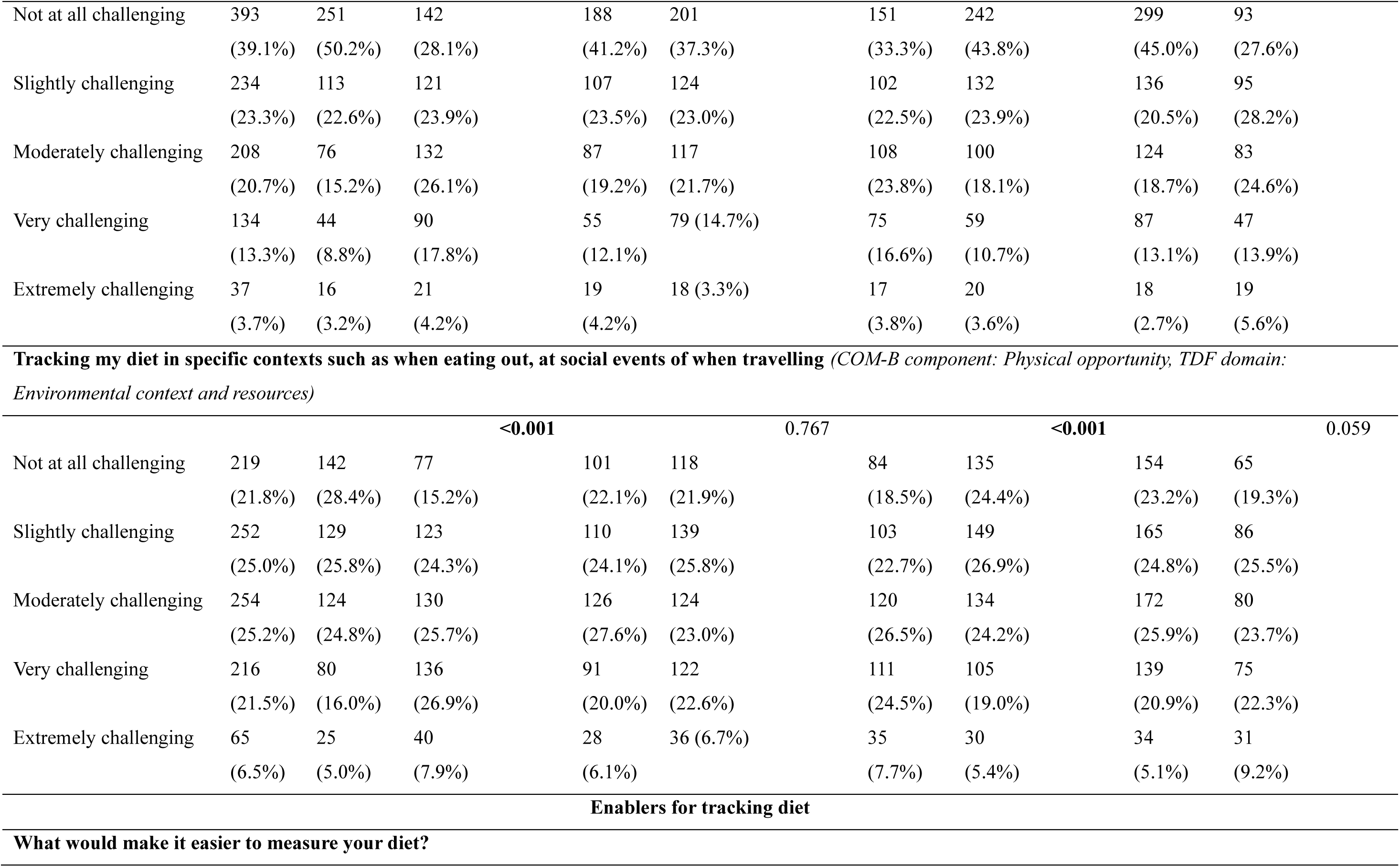
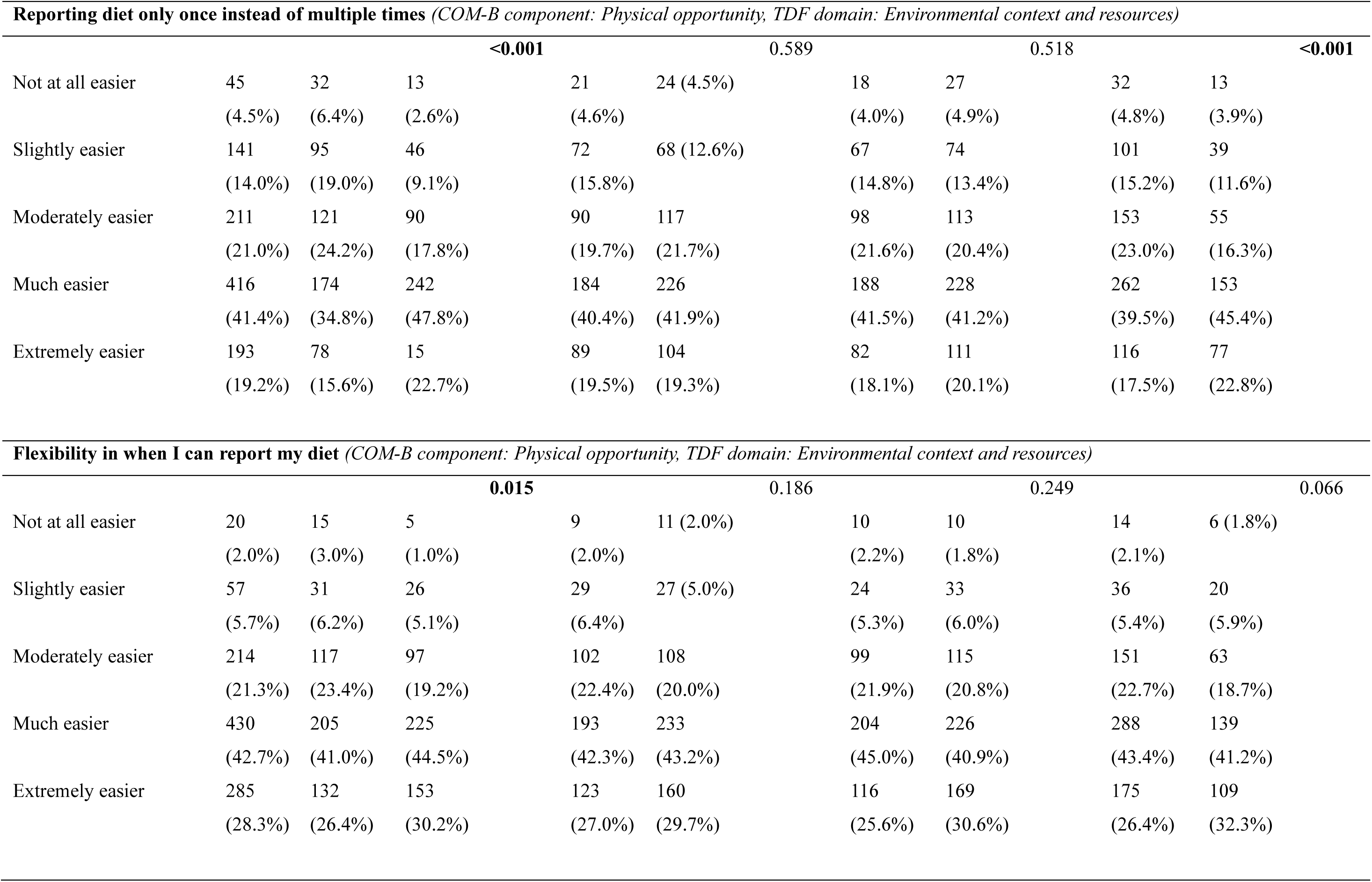
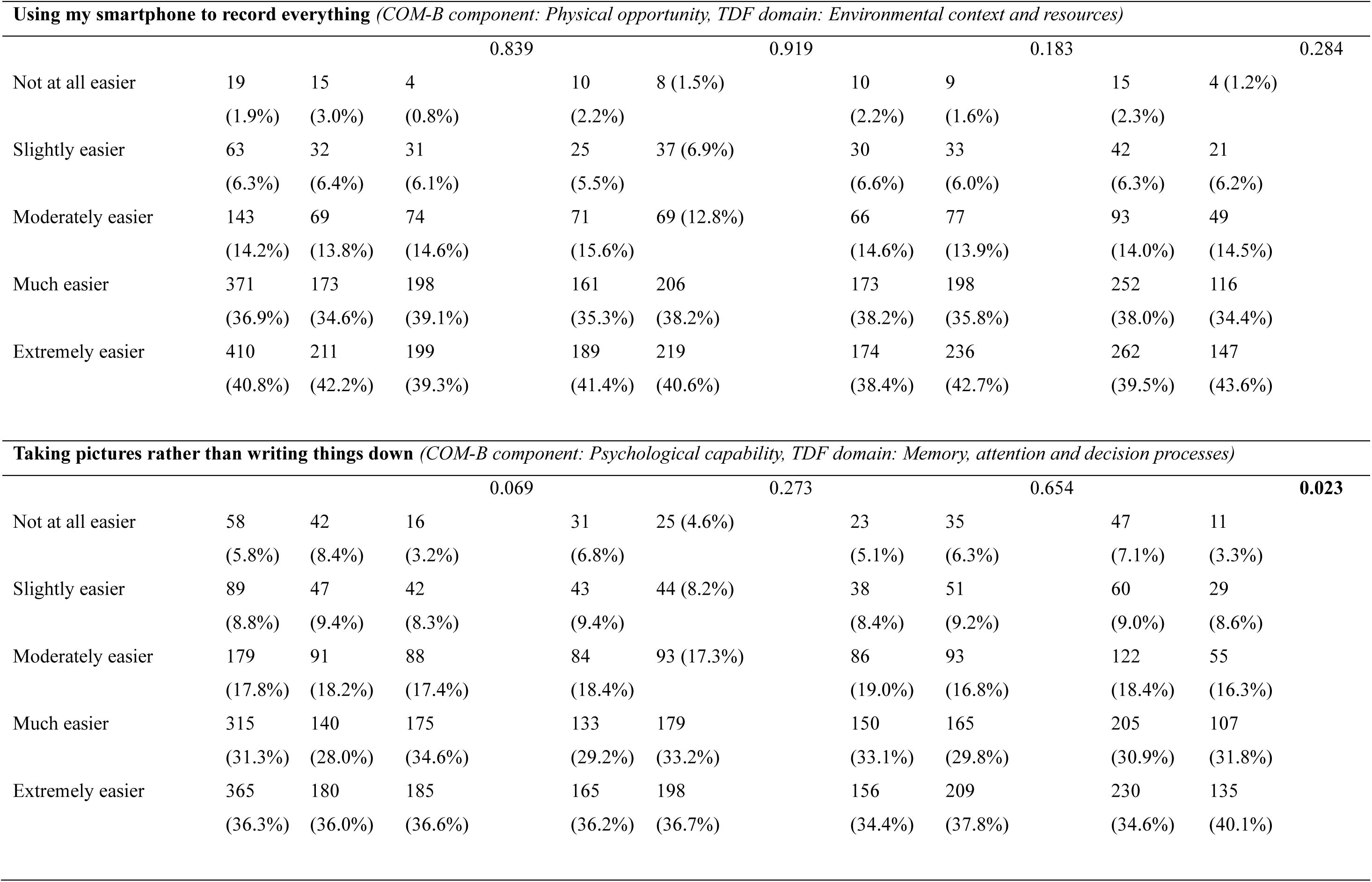
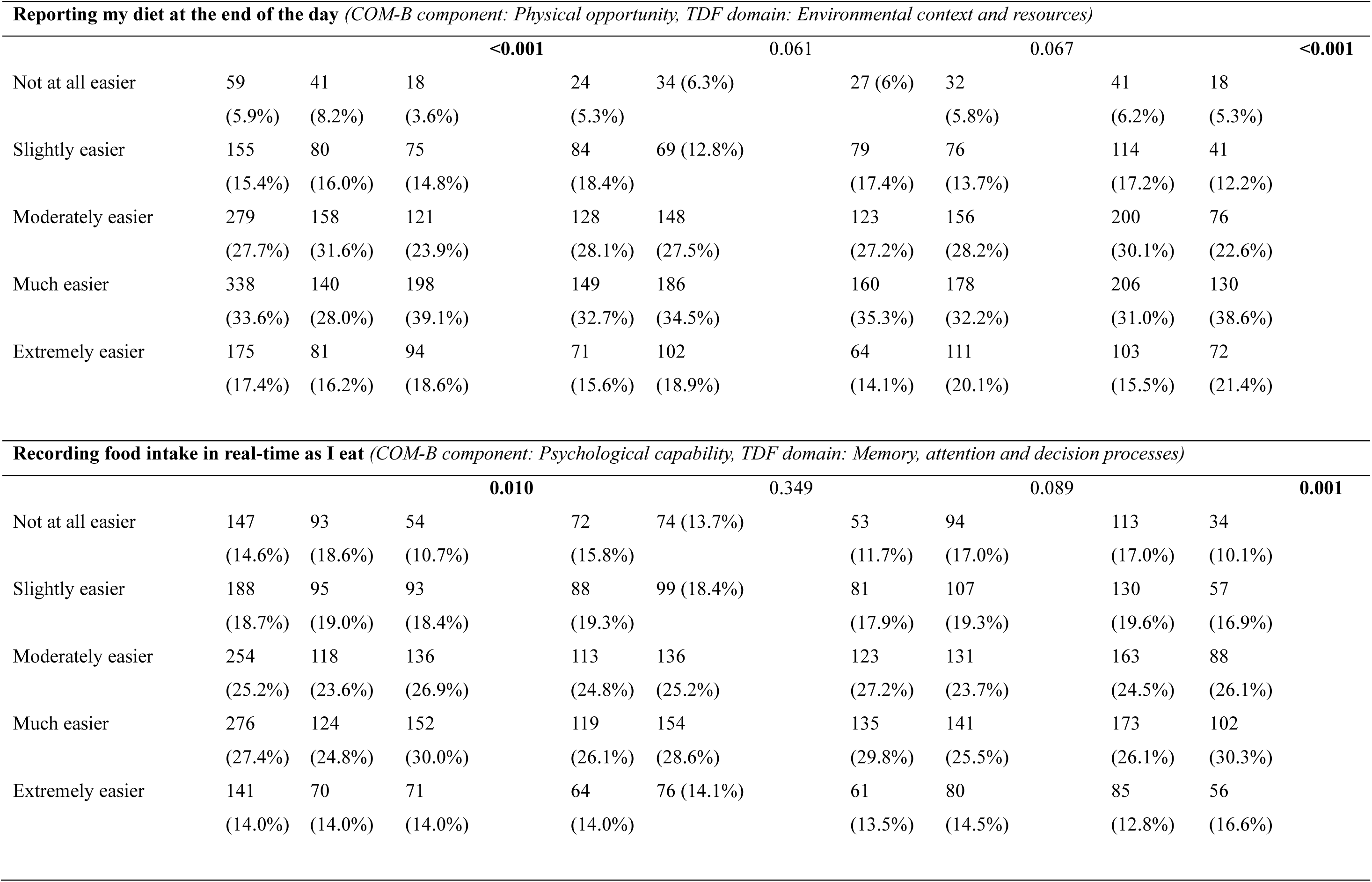
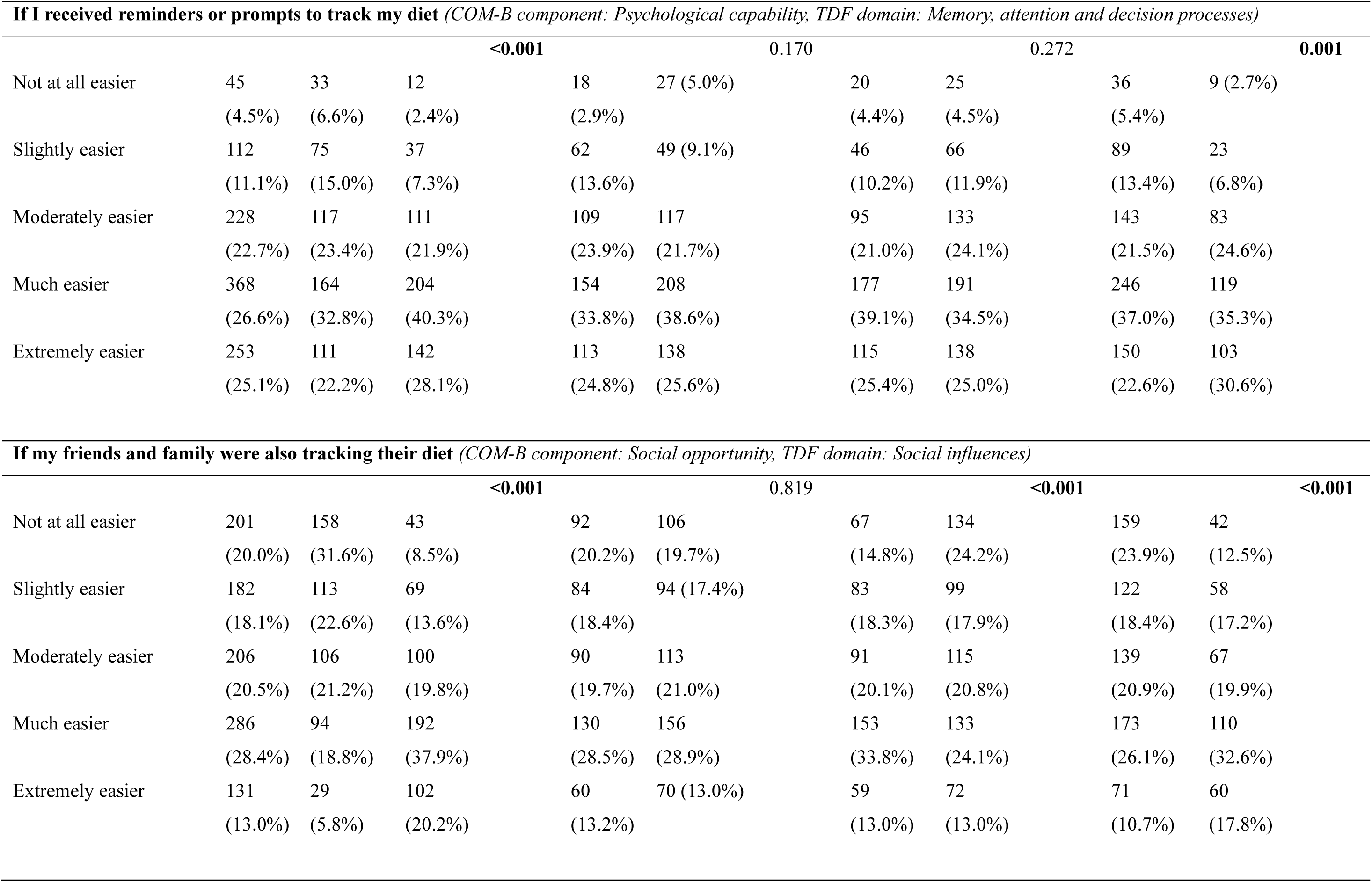
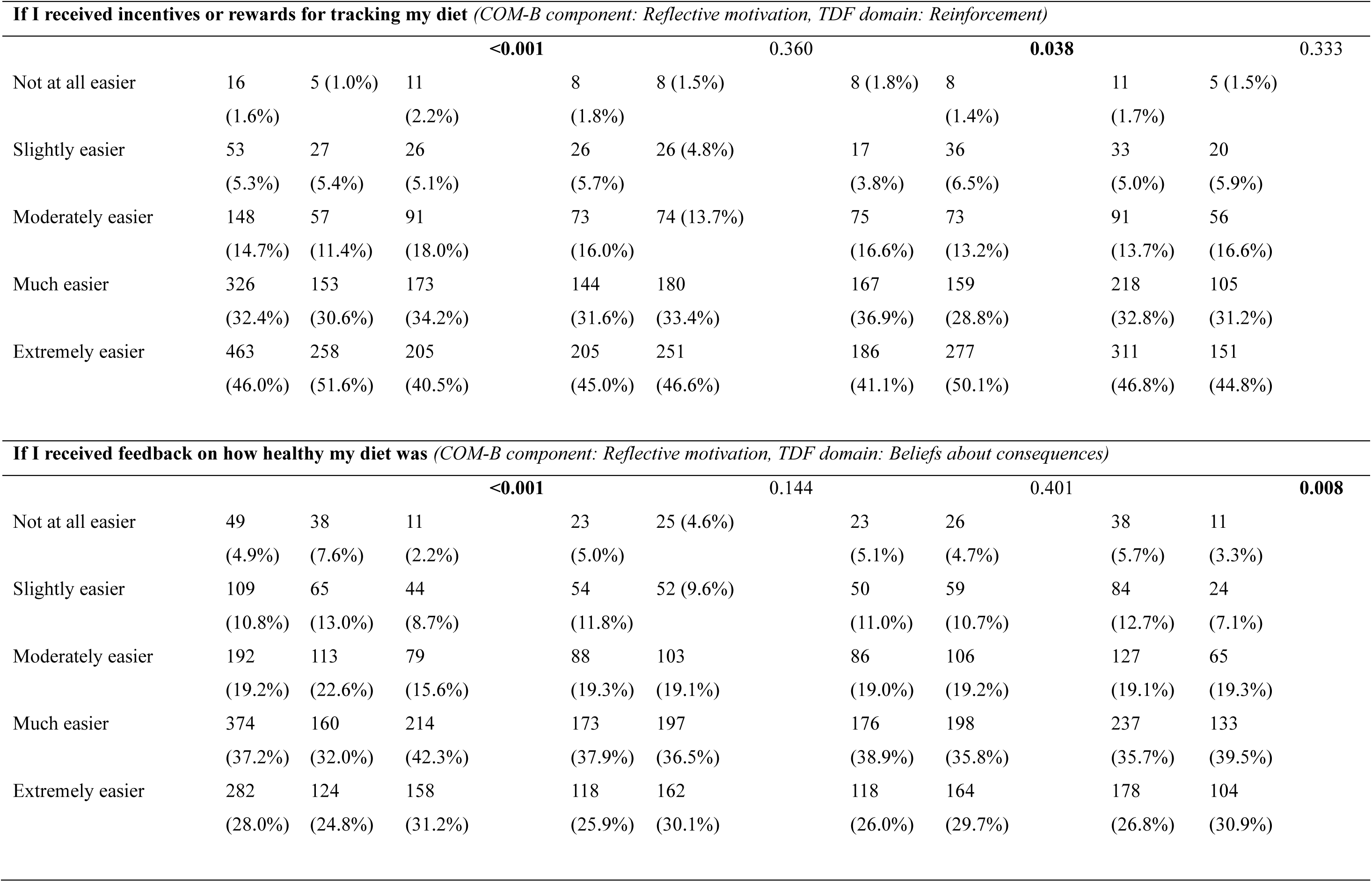
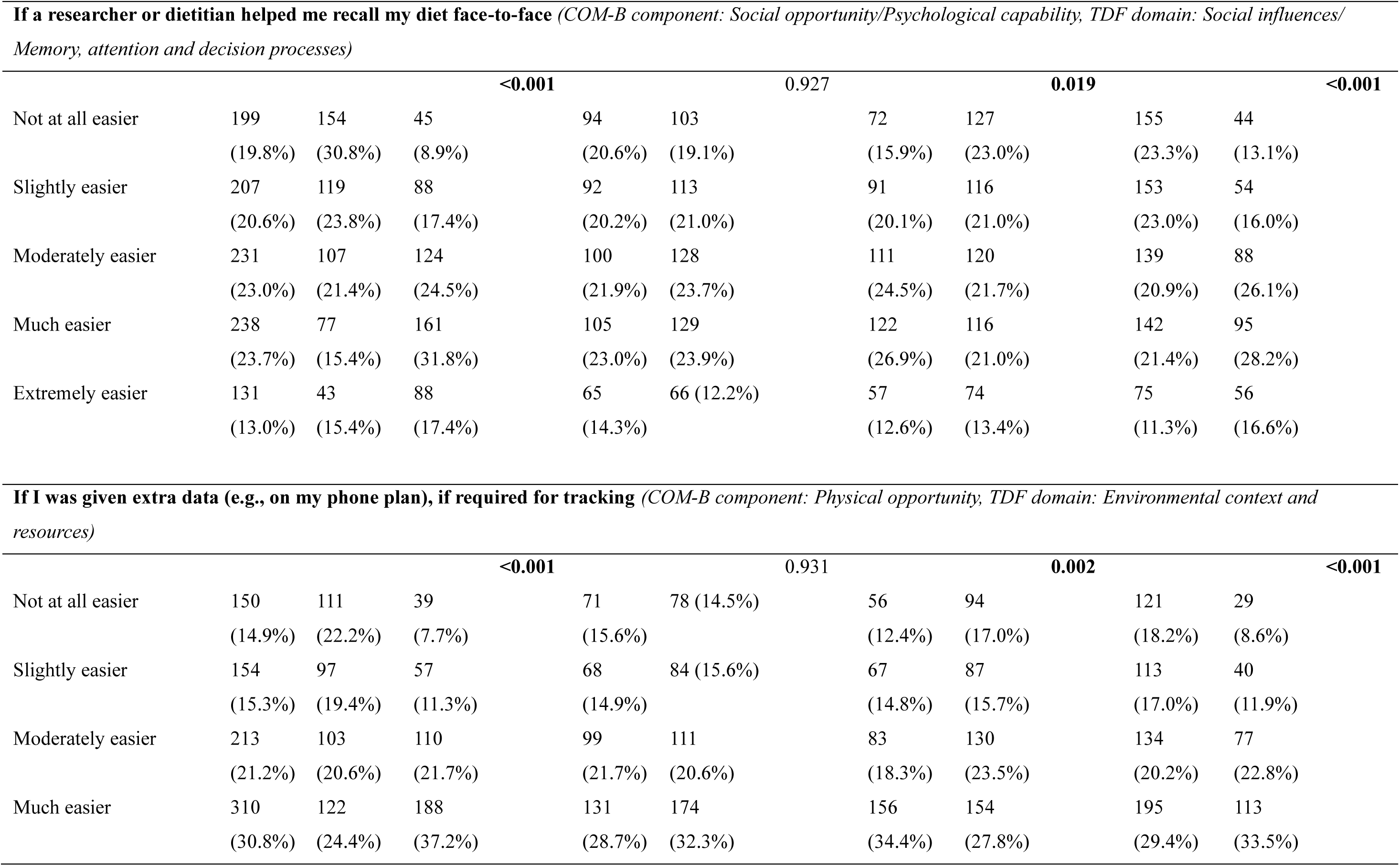
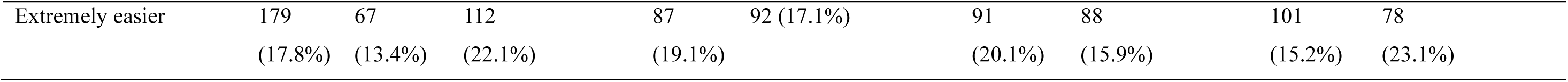
Barriers and enablers for diet tracking, with mapping against the COM-B model and TDF.

### Barriers and enablers for PA tracking

Most barriers to PA tracking were not perceived as major challenges and were typically rated as not at all or slightly challenging (Table 6). The greatest perceived barriers for physical activity tracking were forgetting to record activity levels, day-to-day variability, and privacy concerns. These barriers primarily map onto the TDF domains of ’Memory, attention and decision processes’ (e.g., day-to-day variability, forgetting to record) and ‘Beliefs about consequences’ (e.g., concerns about data sharing). The strongest perceived enablers were automatic tracking without manual logging, receiving incentives or rewards, and having flexibility in when to report activity. These enablers primarily map onto the TDF domains of ’Memory, attention and decision processes’ (e.g., automatic tracking, reminders), ’Reinforcement’ (e.g., incentives/rewards), and ’Environmental context and resources’ (e.g., flexibility, device options).

**Table 6.**
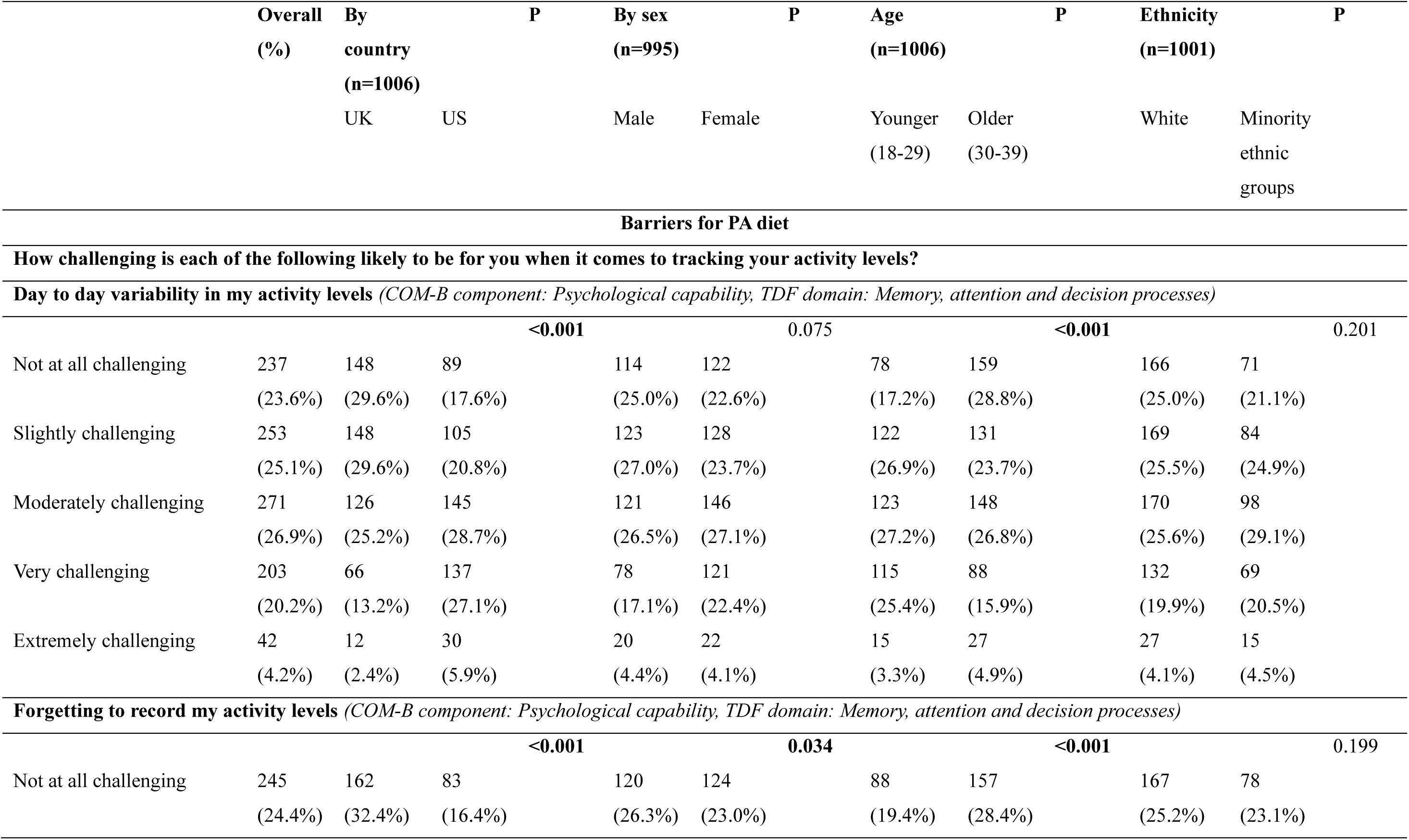
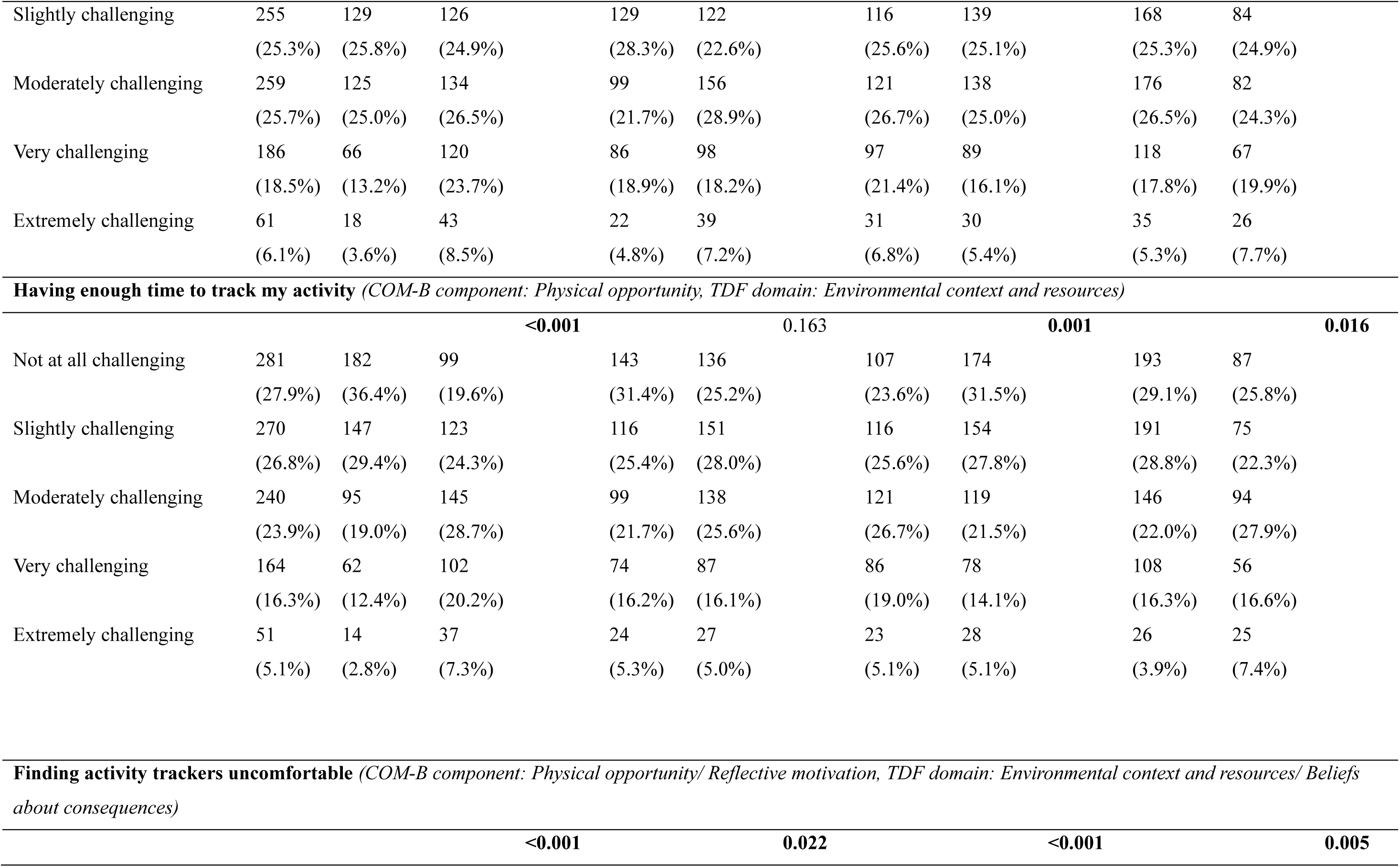
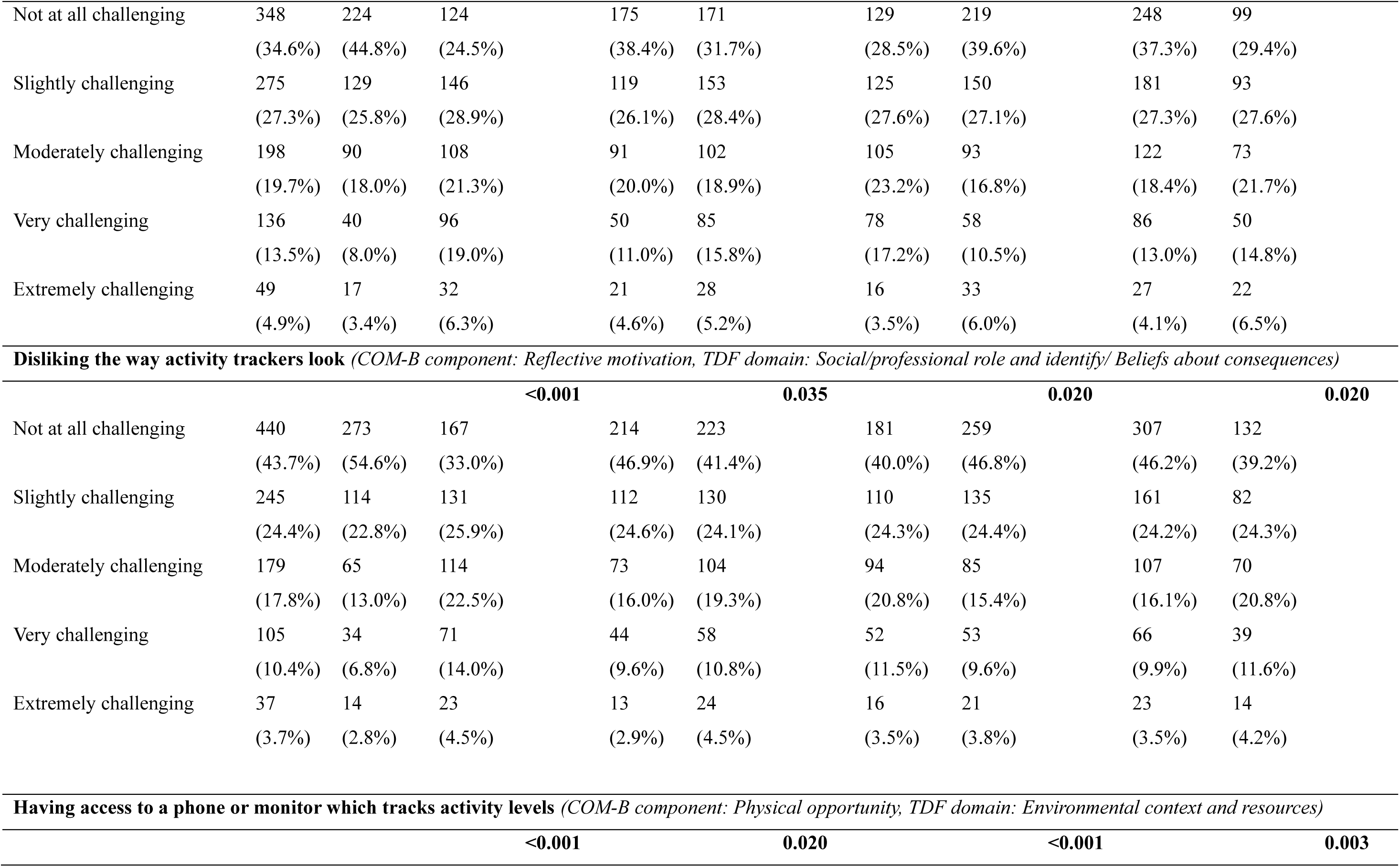
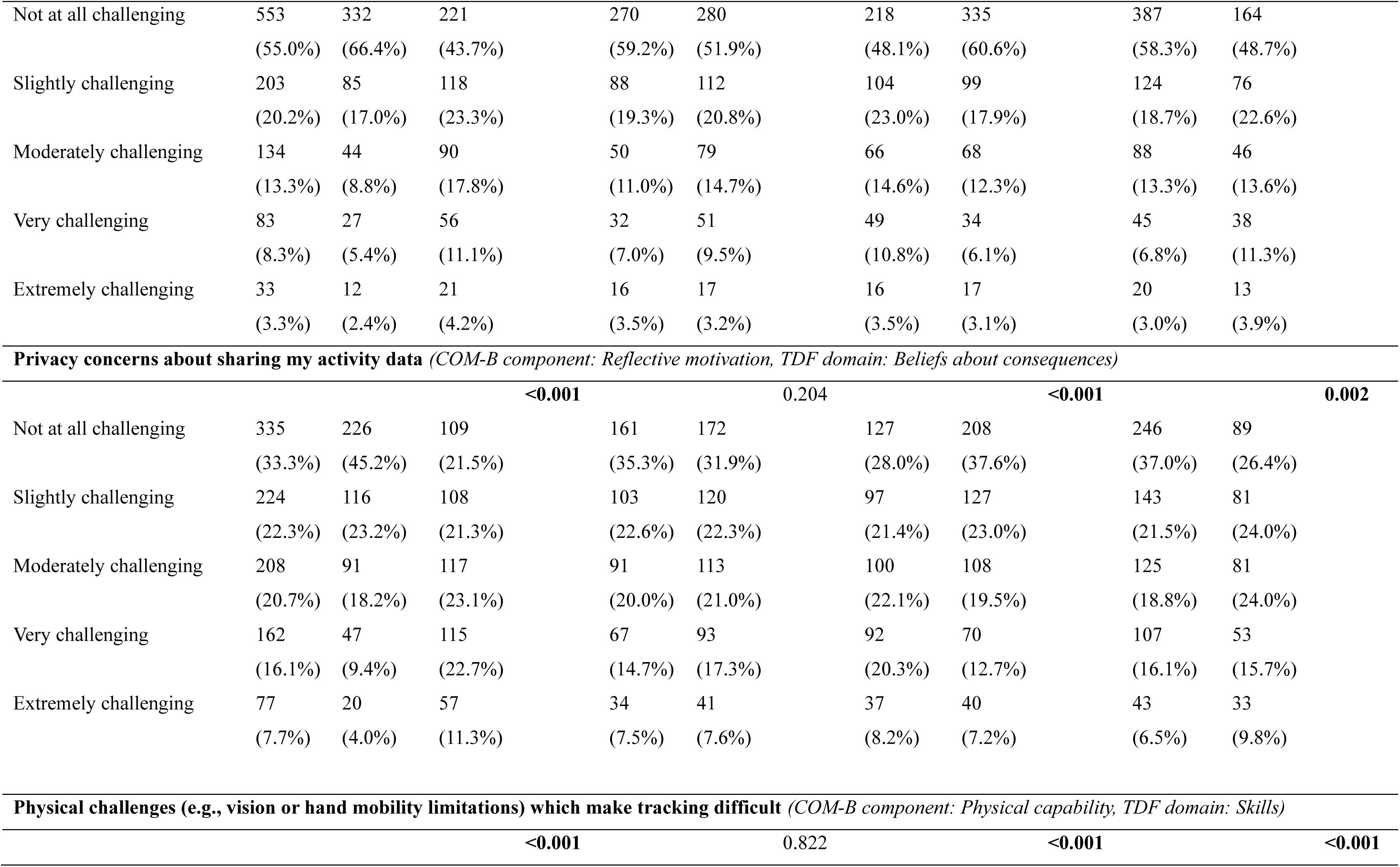
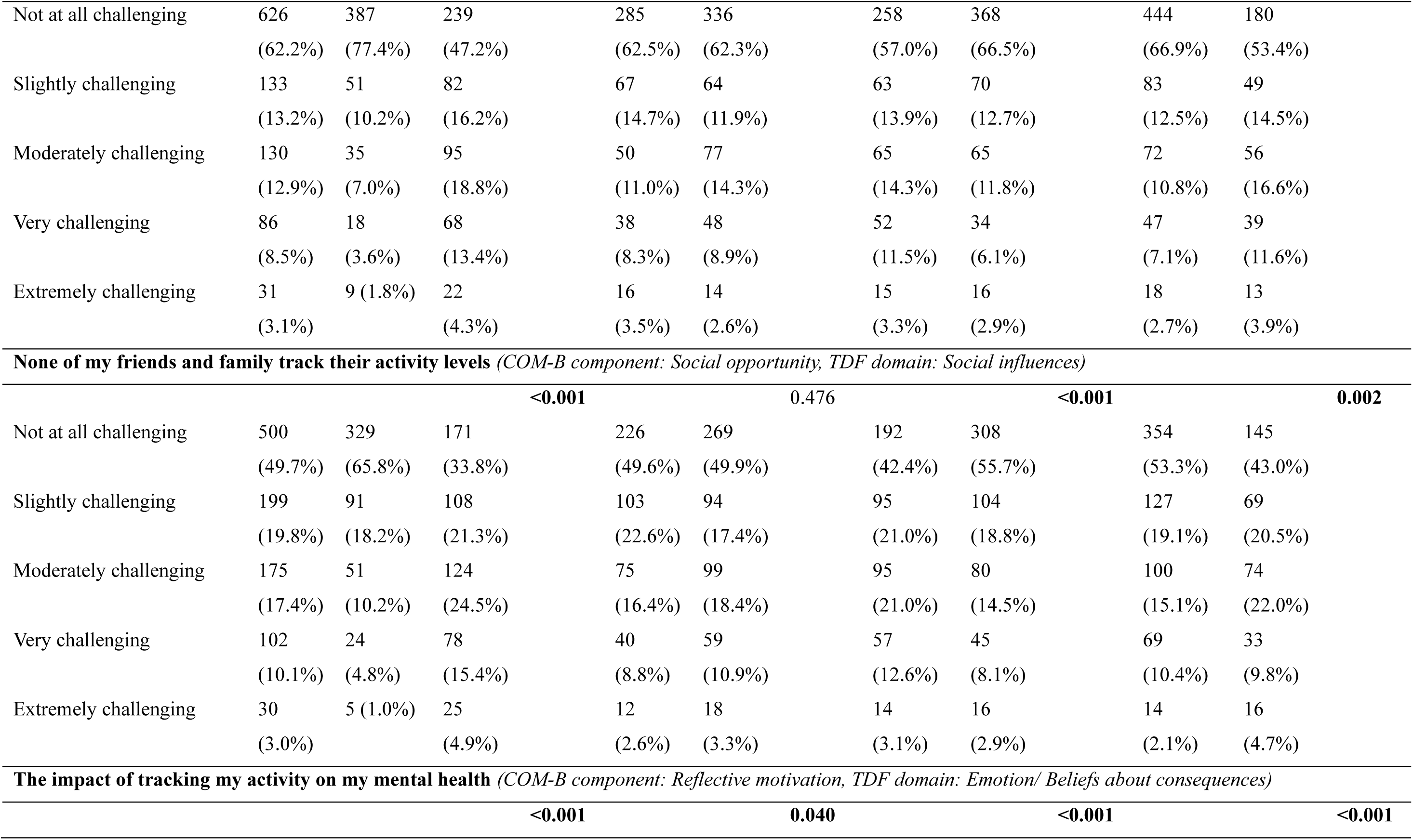
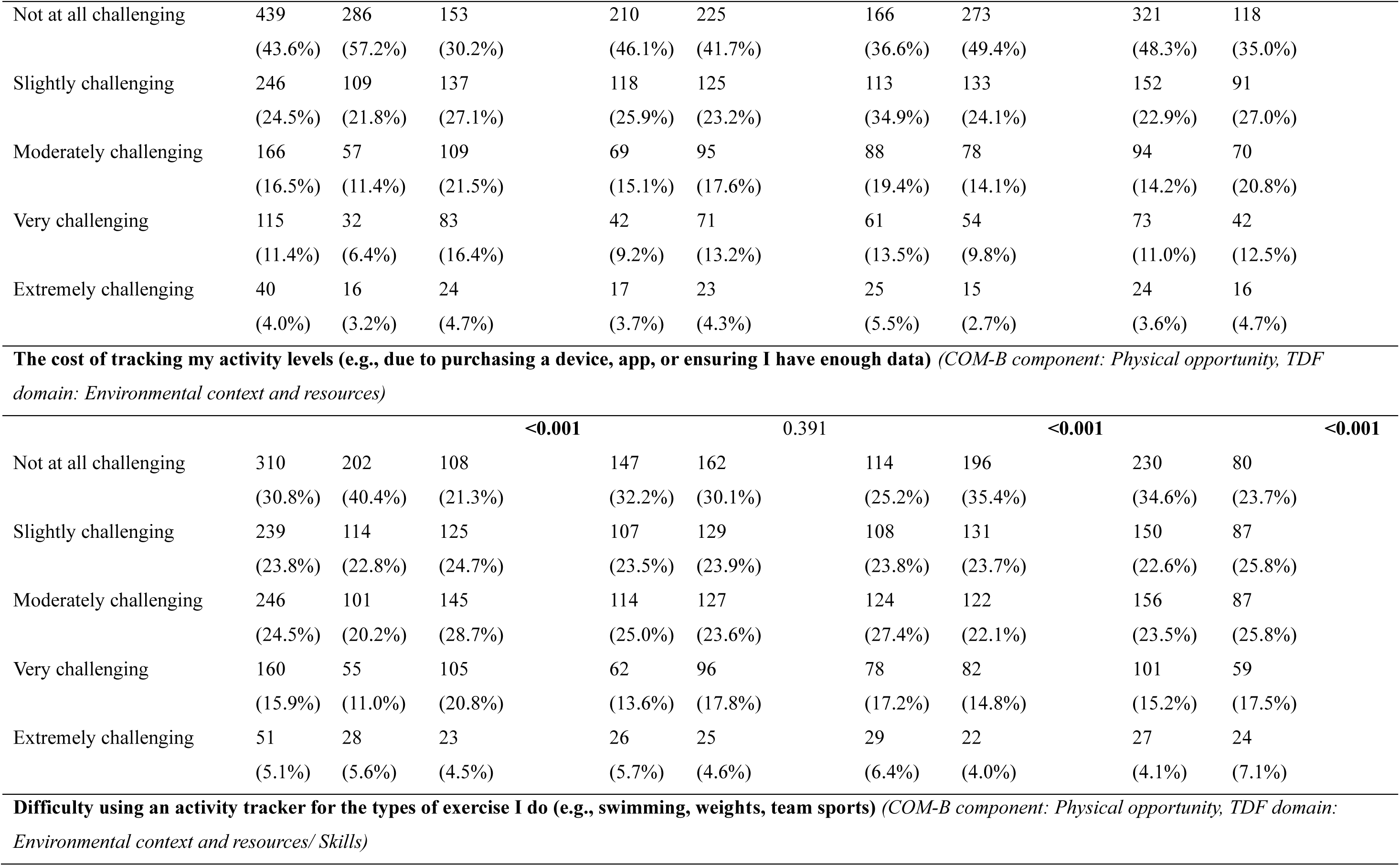
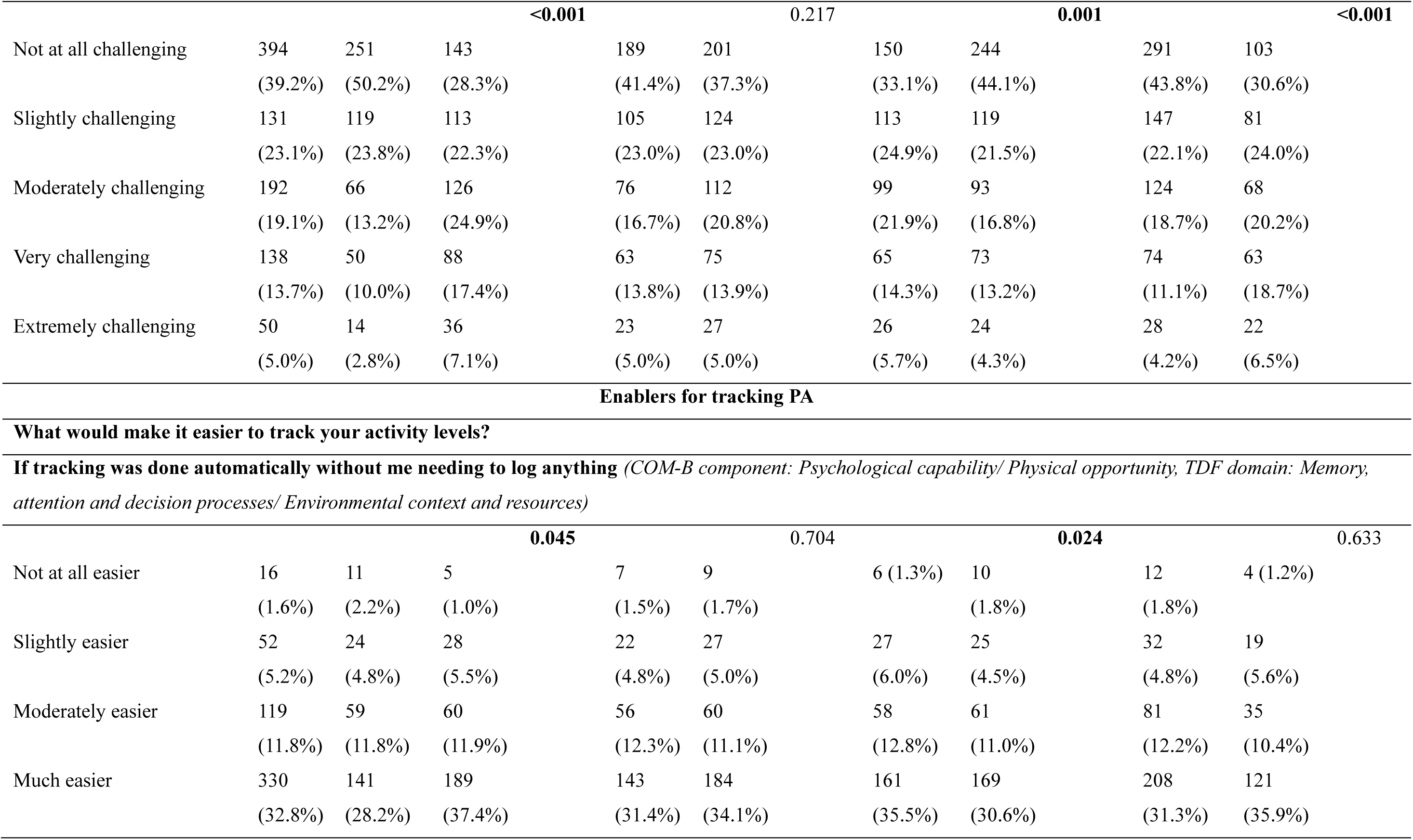
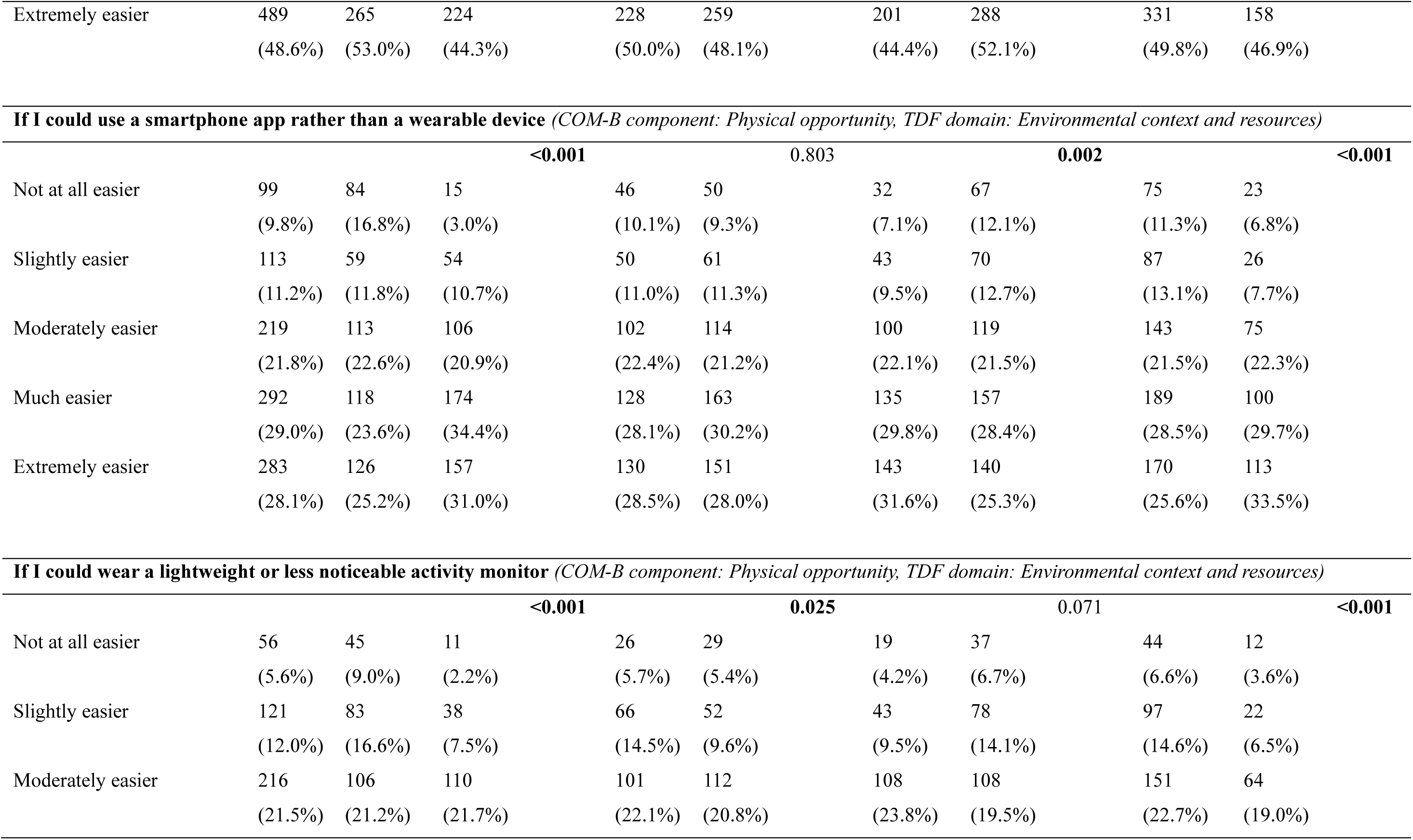
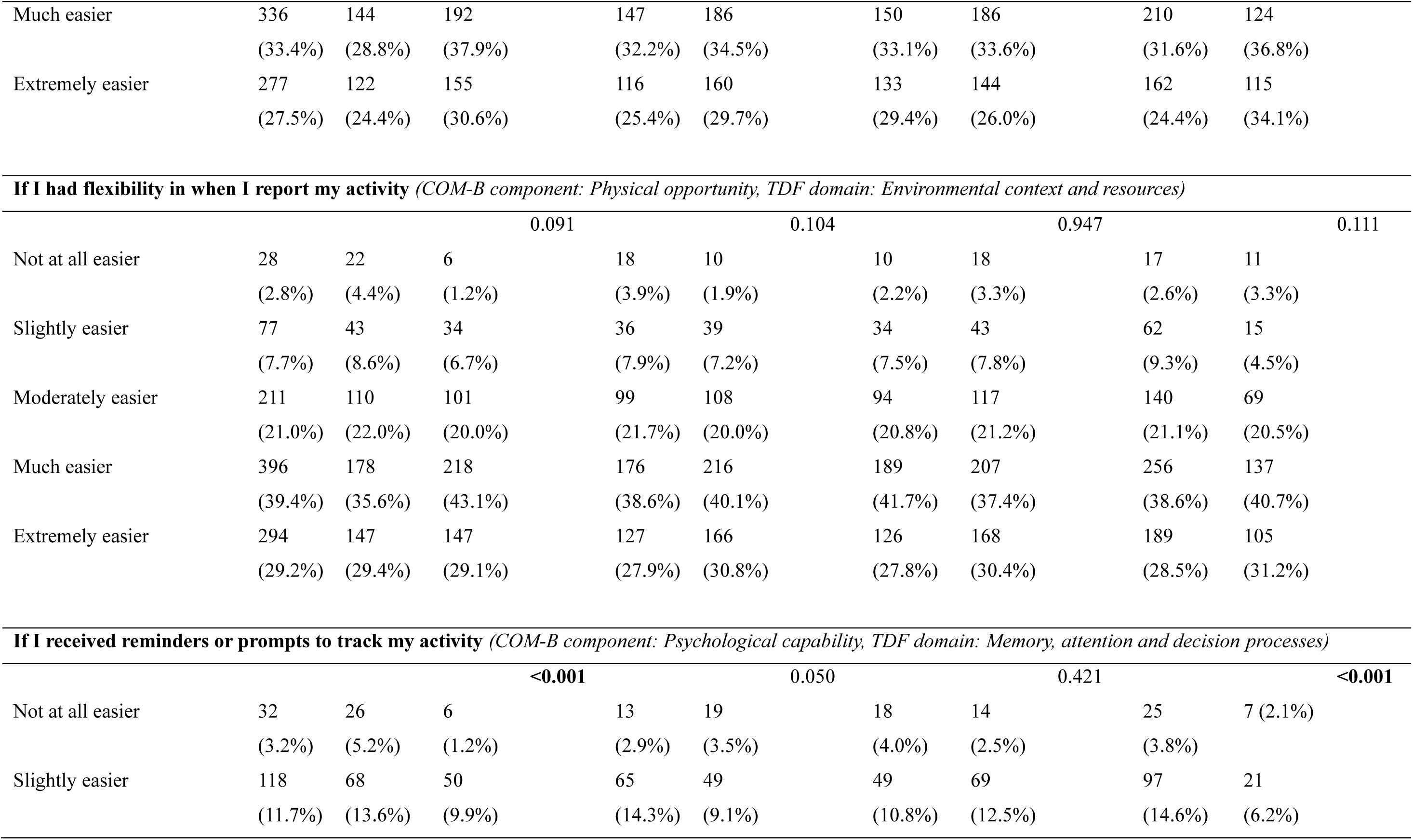
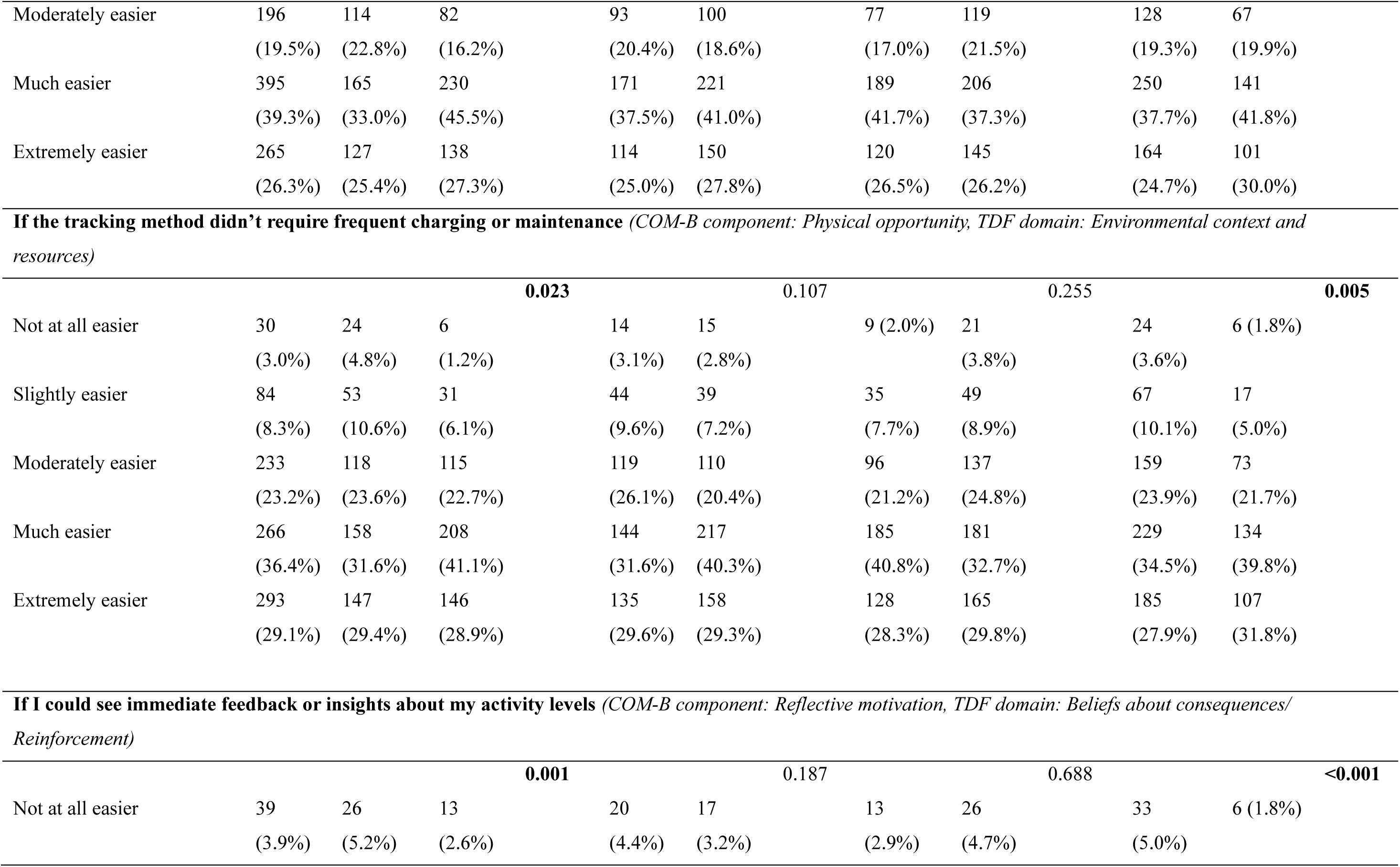
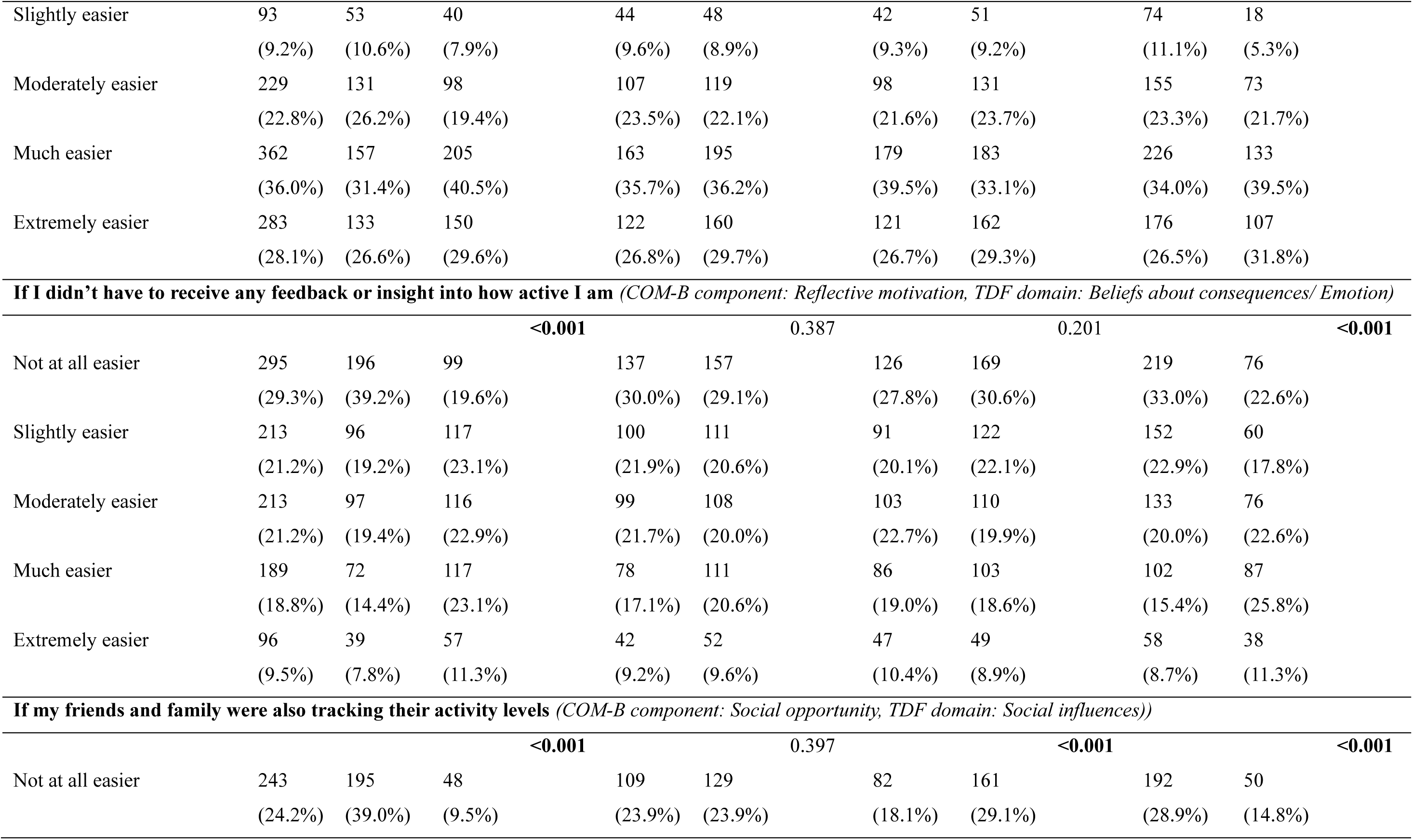
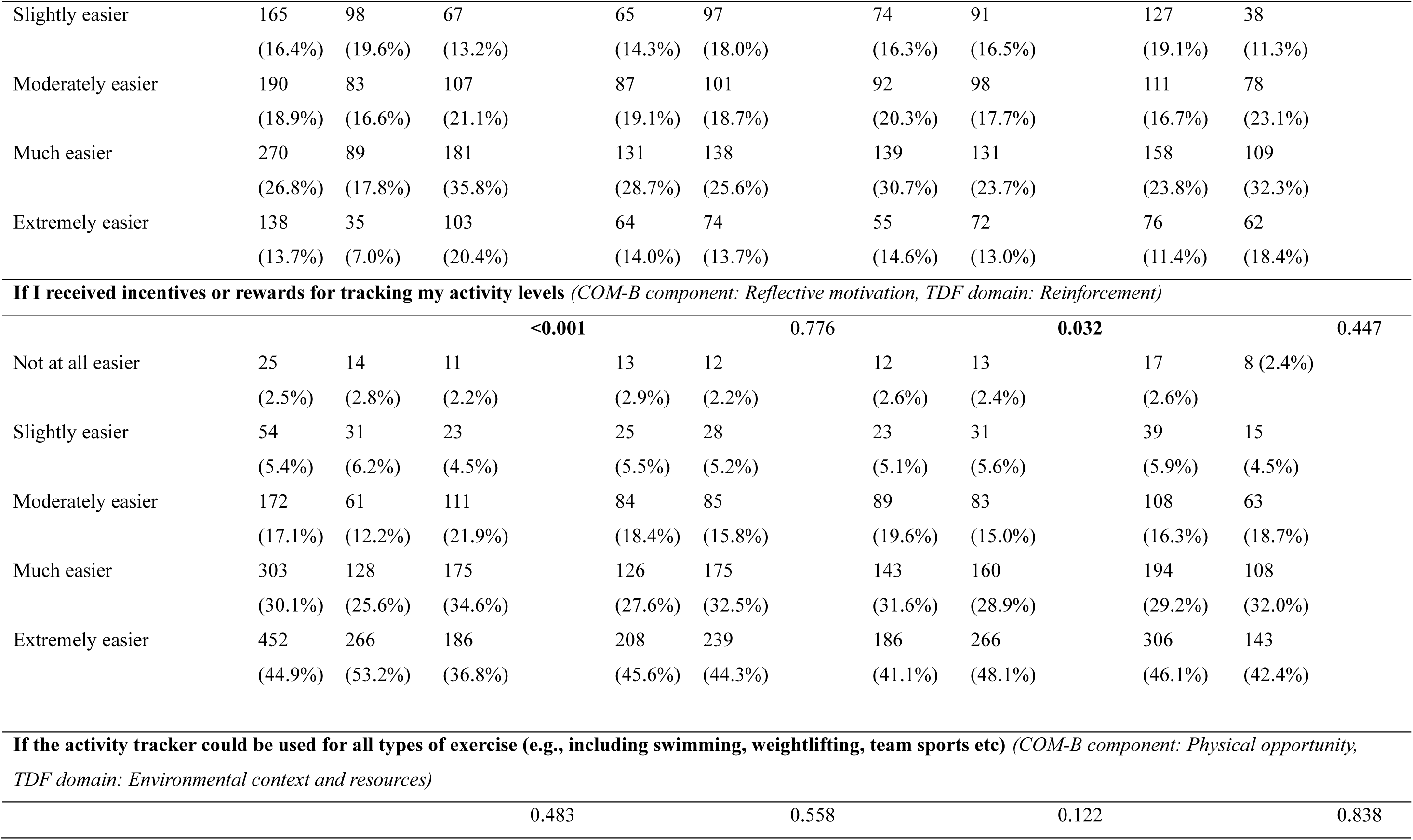
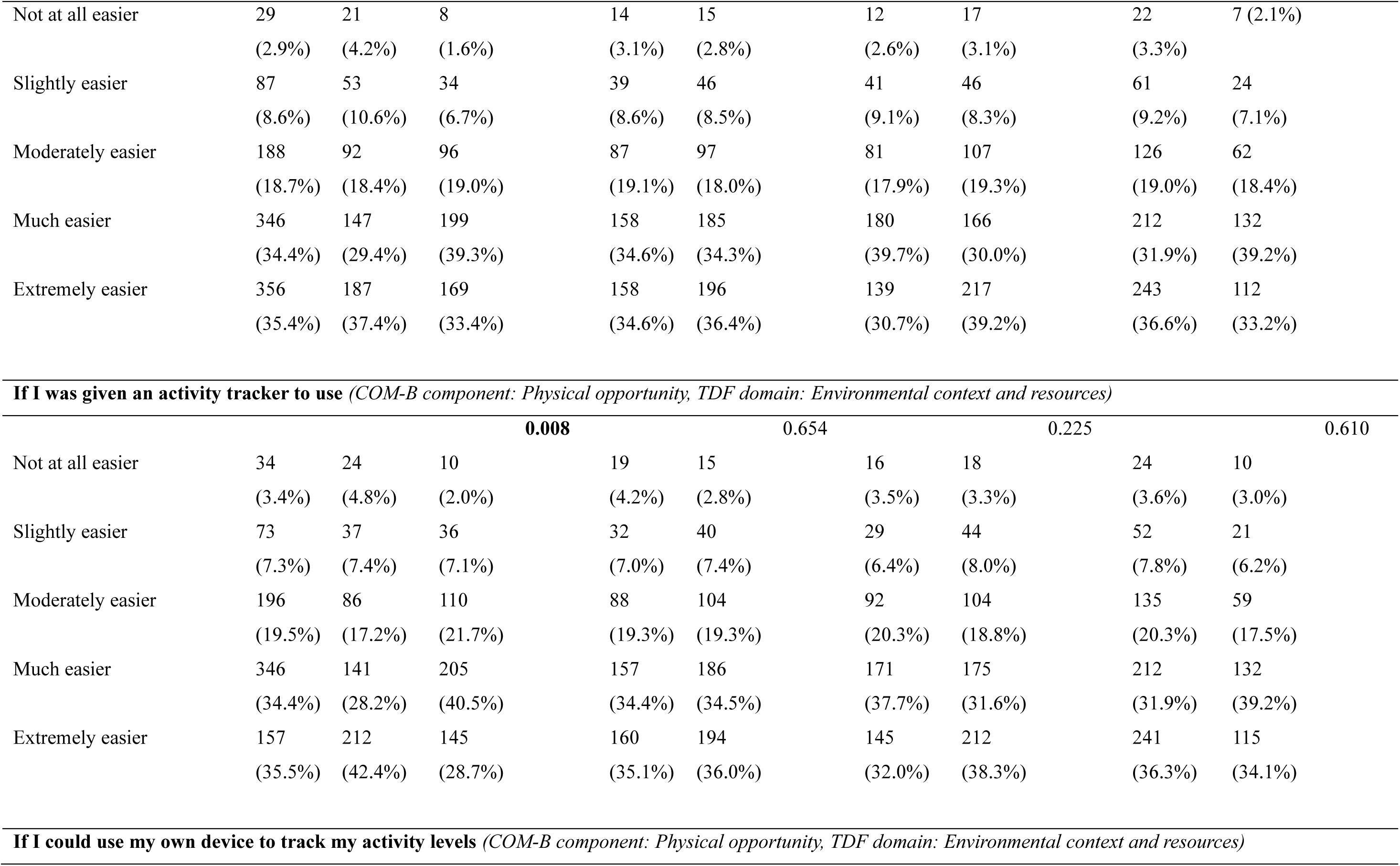
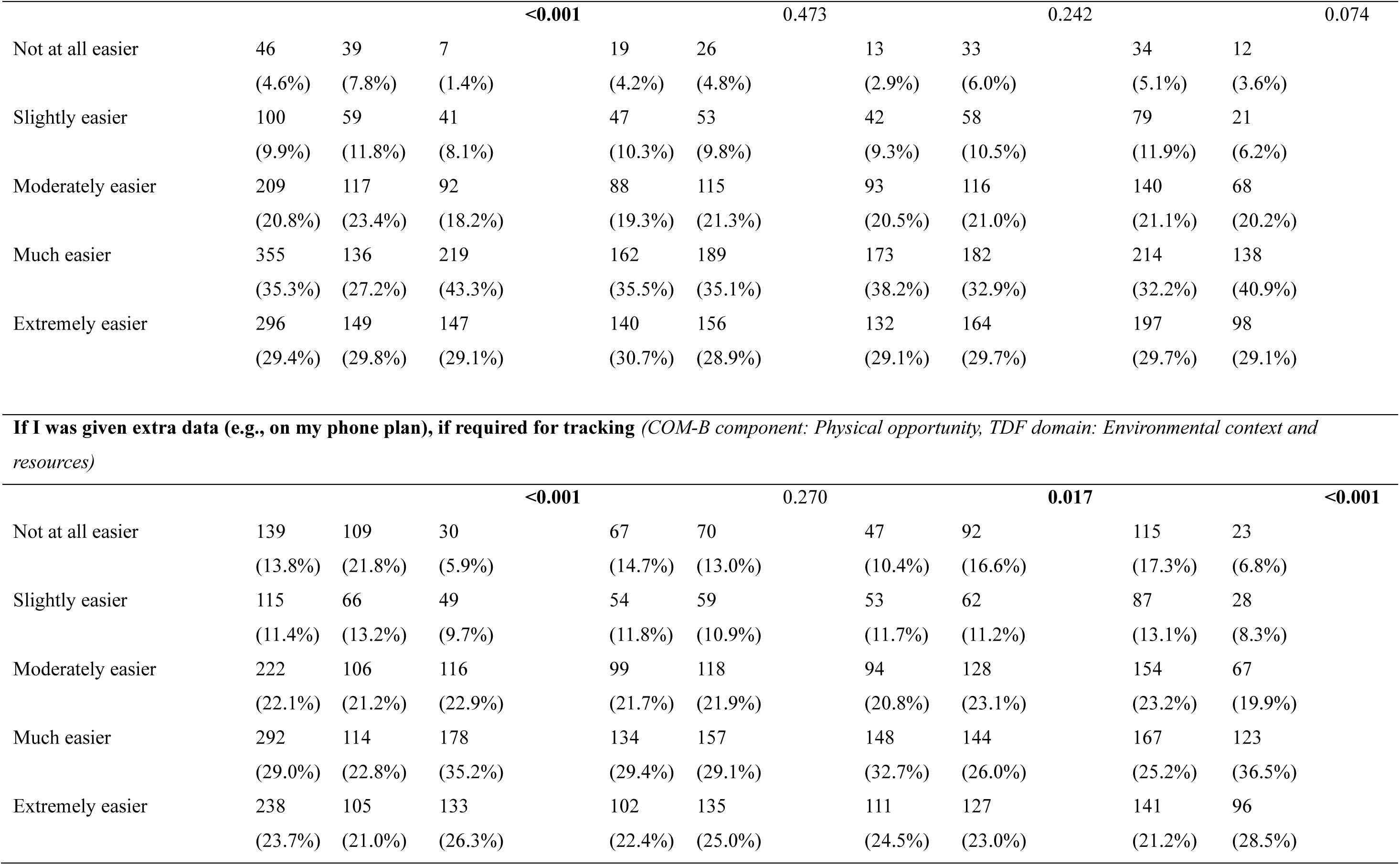
Barriers and enablers for PA tracking

### Differences by country, sex, age and ethnicity

Across all questionnaire domains, the most consistent differences in response emerged between UK and US participants. US respondents typically reported more challenges to both diet and PA tracking. However, they also demonstrated stronger motivation and greater preference for enablers such as reminders, social support, and flexible reporting options. Meanwhile, UK participants showed higher willingness to participate in brain-health research and greater acceptability of remote data-collection methods. UK respondents were also more accepting of most diet and activity assessment tools.

Participants from minority ethnic groups consistently reported greater challenges for diet and PA tracking, particularly around device access, privacy, physical discomfort, and cost. However, they also expressed higher enthusiasm for interactive and supportive enablers, including reminders, social support, and immediate feedback. White participants more frequently rated traditional questionnaire-based assessment methods (e.g., FFQs and PA questionnaires) as highly acceptable and were more willing to provide annual data.

Age-related differences were limited but followed a consistent pattern, with younger adults (18-29 years) reporting more barriers than those aged 30-39 years. This included greater difficulty tracking food and activity outside the home and more frequent concerns about privacy and the mental health impact of tracking behaviours. Sex differences were also limited, with minor differences noted in current behaviours (e.g., men reporting slightly higher activity levels and greater use of tracking for fitness goals) and motivations (e.g., women were more often motivated to track due to weight management). Further breakdown of differences in response between different sub-groups is provided in the Supplementary Material.

## DISCUSSION

This large cross-sectional survey explored attitudes towards, and barriers and enablers to, measuring diet and PA in younger adults, with the explicit aim of informing a future longitudinal study of lifestyle and brain health. Our findings show strong willingness to participate in brain health research and suggest that technological solutions are highly acceptable for capturing diet and activity data in younger adults. Differences in barriers, enablers, and method preferences between demographic groups highlight the need for careful consideration of the most appropriate approaches to optimise recruitment, retention, and data completeness in brain health research to ensure inclusion and equitable representation across diverse population groups.

Technological approaches, particularly smartphone-based approaches, were deemed to be highly acceptable to participants, who already had substantial existing engagement with self-monitoring tools. For example, >70% of participants tracked their diet at least a few times per month, predominantly via apps). Likewise, >70% tracked their PA with similar frequency, commonly using smartwatches or apps. These figures exceed some recent reports of tracking behaviours in younger and middle-aged adults, suggesting our population was particularly tech-engaged. For example, O’Loughlin et al. (2023) found that among 676 young adults in Canada, only 14% and 37% reported past-year diet and PA tracking, respectively. Meanwhile, in a sample of 241 Australian adults (mean age 41 years), Fewings et al. (2022) reported that ∼64% were current or previous users of diet-related apps/websites. Recruiting participants via an online platform in our study is likely to have skewed the group towards those already comfortable with engaging with research in a virtual setting, and may explain some of these discrepancies.

While technology access barriers were less pronounced in our sample, access to technology has been highlighted as a barrier in some other studies, especially in certain population sub-groups. For example, limited access to smartphones or internet connectivity has been reported as a barrier amongst some older adults (Zhou et al. 2025), whilst digital literacy barriers have been noted amongst minority ethnic groups and those with special educational needs (Williams 2006; Islam et al. 2024). Any future brain health focused cohort study which predominantly uses technological/digital tracking methods would therefore need to carefully address potential digital exclusion issues to minimise barriers to participation. Many of the key challenges identified for diet and PA tracking in this study were cognitive and contextual in nature, rather than related to technology. These included day-to-day variability in routines, forgetting to record, and concerns about data privacy. This highlights the potential benefits of a) using passive monitoring tools which track longer-term behaviour (thus accounting for day-to-day variability without requiring participants to deliberately log their diet/PA) and b) having clear and transparent data security procedures, for optimising data integrity, participation and participant confidence in security measures. Specific approaches which may be suitable for diet and PA tracking in this group are explored further below.

Smartphone applications emerged as the most acceptable method for dietary tracking, followed by image-based dietary assessment (e.g., using a smartphone camera), positioning them as promising choices for long-term exposure measurement in future brain health research. Promisingly, both methods could offer solutions to key barriers identified for dietary tracking. For example, for portion size estimation - the greatest perceived barrier - several smartphone compatible dietary recall tools have food images to assist with portion size estimation (Bradley et al. 2016; Wark et al. 2018; Foster et al. 2019). Similarly photograph-based dietary assessment tools reduce burden on participants for quantifying portion size and instead rely upon either a researcher or computer programme to interpret portion size from a photograph. Although both methods still rely upon participants (accurately) recording all that they eat, more advanced passive photography-based methods (e.g., cameras integrated into glasses or clipped to clothing) are beginning to emerge (Hill et al. 2025). Such methods could help further reduce burden and increase participant compliance providing potential challenges around e.g., privacy, battery life and quantifying foods/nutrients within complex foods can be overcome (Wang et al. 2025). Similarly, artificial intelligence-based approaches for categorising complex meals and quantifying portion sizes from photographs are becoming increasingly sophisticated and accurate (Kalpakoglou et al. 2025). As both methods can be used in a range of settings, they may also help address other major barriers including tracking diet outside of the home environment and eating out or getting takeaways.

In contrast to digital methods, which showed superior overall acceptability, biological sampling to assess nutritional biomarkers - particularly via more invasive procedures like venous blood sampling - demonstrated lower acceptance rates. Conversely, less invasive approaches showed greater acceptability, with fasting urine deemed to be particularly acceptable. It is possible that urine sampling could be incorporated periodically or within a subsample of participants to allow biomarker validation without imposing excessive burden. Such validation is particularly valuable for dietary assessment, which remains vulnerable to self-report bias and measurement inaccuracies (Kuhnle 2012). For example, metabolomic fingerprinting approaches show the ability to identify differences in diet quality between participants (Almanza-Aguilera et al. 2017). Similarly, various urine-based biomarkers have been proposed to describe intake of individual foods or food groups, although many of these tend to reflect recent rather than longer-term dietary intake (Jackson et al. 2025).

For PA assessment, smartphone apps were also one of the most attractive options for monitoring, alongside wrist worn devices and completion of an activity questionnaire. Although PA questionnaires can provide low-burden assessment of activity levels, and show moderate agreement with objectively measured PA, they lack granularity and can both under and overestimate specific PA domains (Lee et al. 2011; Helmerhorst et al. 2012). For example, the IPAQ-Short Form - a questionnaire commonly used in cohort studies - overestimates vigorous PA and underestimates sedentary time versus accelerometery (Dyrstad et al. 2014). Smartphone apps and wrist-worn monitors provide objective and accurate assessments of PA, especially when physiological sensors (e.g., heart rate monitors) are incorporated (O’Driscoll et al. 2020; Parmenter et al. 2022). However, accuracy can vary considerably between apps/devices, suggesting the need for careful selection of the most appropriate tool (O’Driscoll et al. 2020; Parmenter et al. 2022). These methods also benefit from aligning with participant preferences for automatic passive tracking, which was viewed as the strongest facilitator for PA monitoring, with most reporting it would make assessment easier. This could be due to reduced cognitive load with passive tracking. Existing cohorts which allow exploration of associations between diet and PA with health (including brain health) outcomes typically measure these exposures at the study baseline or on a limited number of follow up occasions, often several years apart (e.g., (Bingham et al. 2001; Wareham et al. 2002; Bradbury et al. 2018; Le Goallec et al. 2023; Lagström et al. 2025)). Because diet and PA can fluctuate over time, this increases the risk of misclassification error with prolonged follow up periods (Armstrong et al. 2011; Smith et al. 2015). Whilst more frequent assessments would reduce bias, traditional approaches face logistical and financial constraints, compounded by concerns about overwhelming participants with excessive respondent burden (Hasselhorn et al. 2022). Low burden technological assessment methodologies, rated highly by our participants, could help overcome some of these traditional barriers. Interestingly, our findings demonstrate strong participant acceptance for annual assessments of both diet and PA. It is therefore possible that with the right tools and measurement strategy, more frequent assessment could be possible.

Many existing cohort studies are limited by a lack of socioeconomic and ethnic diversity, making it difficult to generalise findings to wider populations (Krishnan et al. 2024). Ensuring better representation of minoritised groups is therefore a key priority for future brain health and lifestyle cohort studies, with careful consideration needed to maximise both recruitment and sustained engagement. In the present study, significant demographic differences were observed in both barriers and enablers to diet and PA tracking, especially between UK and US participants. US respondents generally reporting more challenges but also stronger motivation and responsiveness to enablers such as reminders and social support. From an equity perspective, participants from minority ethnic groups consistently reported greater challenges across most domains, yet showed higher enthusiasm for interactive and supportive enablers such as reminders, feedback, and social support, consistent with previous findings that minority ethnic groups may face structural barriers but engage strongly with socially supportive and interactive forms of digital health delivery (Ellis et al. 2022; Aldosari et al. 2023). Adults aged 18–29 years also reported greater challenges compared with those aged 30–39 years - particularly regarding privacy, cost, and tracking in social contexts - highlighting that life-stage factors such as irregular schedules, financial constraints, and sensitivity to perceived surveillance shape engagement more than technical proficiency (Kanstrup et al. 2018; Jeong and Nam 2024). Collectively, these findings demonstrate that even within a relatively young and digitally literate cohort, barriers and facilitators to diet and PA tracking are not uniform, and highlight the need for inclusive, context-sensitive design strategies to ensure equitable participation and data quality in future large-scale cohort studies.

### Strengths and limitations

Strengths of this study include the large and diverse sample, with participants from both the UK and US, which provides insight into potential challenges and opportunities for tracking diet and PA in different population groups and settings. We surveyed all participants on the same day, minimising temporal influences on responses. Our questionnaire was designed with reference to behaviour change theory (e.g., COM-B model and TDF) to ensure that we captured key barriers and enablers for evaluating diet and PA in younger adults. Collection of detailed demographic data also enabled sub-group analyses to understand geographical, age, sex and ethnicity-related factors likely to affect diet and PA tracking in a future brain health cohort study. Limitations include the convenience sampling used for participant recruitment and exclusively online nature of the survey, which limits generalisability to other population groups including those with limited digital literacy/access. Participants’ responses were based on their perception of different hypothetical study features, and pilot testing would be needed prior to carrying out any large-scale cohort study to ensure feasibility. We were unable to conduct more detailed analyses within specific ethnic groups and instead compared white participants with those from minority ethnic groups. Consequently, important differences between individual ethnic groups may have been overlooked. Finally, we focused here on which methods may be most acceptable to participants and the design of any future cohort study will need to also consider validity, reliability, and cost-effectiveness of different assessment methods.

## Conclusion

This study provides insight into the potential challenges and opportunities for measuring diet and PA in the context of a future prospective cohort study to identify risk and resilience factors for poor brain health in later life. It demonstrates considerable interest in participating in such a study with >90% of our respondents suggesting that they would be likely or very likely to participate in a brain health focused cohort study. Our findings also highlight the high acceptability of technological approaches including those which use smartphones to allow regular tracking of diet and PA. Carefully addressing behavioural and structural barriers, and leveraging passive/low burden monitoring approaches, will be crucial to creating a diverse, scalable cohort capable of characterising lifestyle-brain health pathways from young adulthood onwards. Pilot testing data collection methods is now warranted to assess and optimise feasibility and engagement.

## Data Availability

Data are available from the authors on reasonable request

## ACKNOWLEDGEMENTS

This study was supported by an Alzheimer’s Research UK North Network Engagement/Involvement Grant and a Network Grant from the Royal Society of Edinburgh. KCs salary is supported by a Major Project Grant from Alzheimer’s Research UK (ARUK-MPG2024-008).

Acknowledgements **N/A**

## Declarations

**Funding:** This study was supported by an Alzheimer’s Research UK North Network Engagement/Involvement Grant and a Network Grant from the Royal Society of Edinburgh. KCs salary is supported by a Major Project Grant from Alzheimer’s Research UK (ARUK-MPG2024-008).

**Conflicts of interest:** OMS has received research grants from EPSRC, BBSRC, Rank Prize, MRC, Wellcome Trust, NIHR, ARUK, OHID, the Fruit Juice Science Centre and the Nutrition Society. He has carried out paid consultancy (paid to institution) for Delta Hat Ltd and is a Section Chair for the Nutrition Society. SG is an employee of Scottish Brain Sciences. All other authors declare no conflicts of interest.

**Ethics approval:** This study was conducted according to the guidelines laid down in the Declaration of Helsinki and all procedures involving research study participants were approved by the Newcastle University Ethics Committee (REF: 57735/2023).

**Consent to participate:** Written informed consent was obtained from all subjects/patients prior to data collection.

**Consent for publication:** Not applicable

**Availability of data and material:** Data are available from the authors on reasonable request.

**Code availability:** Analytic code is available from the authors on reasonable request.

**Author contributions:** The study was conceived by OMS and SG. The online questionnaire was developed by the same authors, with further input from AG, JM, KB, LB and FRF. Data were collected by OMS and SG. All authors contributed to data interpretation. OMS analysed the data and drafted the manuscript, which was critically revised by AG, JM, KC, KB, LB, FRF and SG. All authors approved the final version of the manuscript.

## Notes

### Funding Statement

This study was supported by an Alzheimers Research UK North Network Engagement/Involvement Grant and a Network Grant from the Royal Society of Edinburgh. KCs salary is supported by a Major Project Grant from Alzheimers Research UK (ARUK-MPG2024-008).

### Author Declarations

The Faculty of Medical Sciences Research Ethics Committee of Newcastle University gave ethical approval for this work (REF: 57735/2023).

